# Modeling impact and cost-effectiveness of gene drives for malaria elimination in the Democratic Republic of the Congo

**DOI:** 10.1101/2020.06.29.20142760

**Authors:** Nawaphan Metchanun, Christian Borgemeister, Gaston Amzati, Joachim von Braun, Milen Nikolov, Prashanth Selvaraj, Jaline Gerardin

## Abstract

Malaria elimination will be challenging in countries that currently continue to bear high malaria burden. Sex-ratio distorting gene drives, such as driving-Y, could play a role in an integrated elimination strategy if they can effectively suppress vector populations. Using a spatially explicit, agent-based model of malaria transmission in eight representative provinces of the Democratic Republic of the Congo, we predict the impact and cost-effectiveness of integrating driving-Y gene drive mosquitoes in malaria elimination strategies that include existing interventions such as insecticide-treated nets and case management of symptomatic malaria. Gene drive mosquitoes could eliminate malaria and were the most cost-effective intervention overall if the drive component was highly effective with at least 95% X-shredding and associated cost of deployment below 7.17 $int per person per year. Suppression gene drive could be a cost-effective supplemental intervention for malaria elimination, but tight constraints on drive effectiveness and cost ceilings may limit its feasibility.

## Introduction

Female *Anopheles* mosquitoes can transmit *Plasmodium* parasites that cause malaria, a life-threatening infectious disease. The most commonly used vector control methods to prevent mosquito bites are sleeping under insecticide-treated mosquito nets (ITNs) and spraying the inside walls of a house with an insecticide (indoor residual spraying, IRS) (1). Treatment of symptomatic malaria cases with artemisinin-based combination therapy (ACT) can effectively manage malaria burden, although access to prompt and quality care remains a barrier.

Nevertheless, despite being preventable and treatable, with considerable control successes during the last 20 years (2,3), malaria still has devastating impacts on health and livelihoods of people around the world. The World Health Organization (WHO) estimates that about 3.7 billion people are at risk of the disease in 97 predominantly tropical countries (3,4), even though billions of dollars are spent annually on malaria control and elimination. Most malaria cases occur in sub-Saharan Africa (SSA), accounting for 93% of total malaria cases worldwide (3,5). With 12% of all cases in SSA, the Democratic Republic of the Congo (DRC) is the second highest-burden country on the continent (3). Nearly all of the DRC’s population lives in high malaria transmission zones (6). Consequently, the disease remains one of the country’s most serious public health problems and is the number one cause of death (7,8).

Despite sustained malaria control, malaria incidence in the DRC has increased in the last few years (3), and more than 40% of children who fell ill because of malaria did not receive adequate care (3,9). Health system weaknesses and gaps in the coverage of core interventions caused by financial and programmatic limitations are likely responsible for this recent rise in cases (10), and elimination remains elusive. Sustained access to vector control has been a central strategy in the DRC’s complex operating environment, where challenges are compounded by domestic political conflicts (8) and insufficient funding for malaria control (11). These challenges emphasize the urgent necessity of developing new strategies for malaria control and elimination for the DRC and beyond (3,10,12,13).

Transgenic mosquitoes carrying gene drives have recently been successfully developed in the laboratory (14). Gene drive is a novel method that involves the inheritance of specific traits from one generation to the next at rates higher than the 50% chance afforded through Mendelian inheritance in heterozygotes, and gives certain genes a substantially higher or lower probability of inheritance and thereby alters the frequency of such genes in the population. If given a gene that could alter fertility or survival of the target species, this could alter its population size over a few generations (15,16). Given rising resistance to existing insecticides and antimalarial drugs (17–20), gene drive mosquitoes might hold great potential to accelerate and achieve lasting gains in malaria control (14). The future of gene drives itself also depends on the economic aspect of the technology compared with existing or future alternatives (21). This study assesses the cost-effectiveness of gene drives together with conventional interventions by estimating Disability-Adjusted Life Years (DALYs), DALYs averted, and cost-effectiveness of vector control methods in the DRC.

Although gene drive has yet to pass the research and development stage, with driving-Y gene drive has yet to be developed in the laboratory and lead candidates of gene drives are only now in confined cage trials (22), public concern has been voiced over gene-related technologies. Concerns on previously developed genetic controls, such as a genetically modified version of *Aedes aegypti* for control of mosquito-transmitted arboviral diseases, has led to a debate on whether the technology is suitable for a large-scale implementation (23). At the same time, proof of efficacy presents a challenge, and informed decision-making on gene drive releases into the wild will require additional information about potential effectiveness (14,24).

Disease modeling is a powerful tool that can complement laboratory findings and help develop control strategies involving transgenic mosquitoes. The scientific community, including the WHO and other policy groups, has increasingly recognized the importance of disease modeling in guiding the development of gene drives and genetically modified organisms (14,24,25). In this work, we explore the possible outcomes of applying gene drives as an intervention for malaria control in representative SSA settings in combination with established control programs - including ITNs and ACT distributions - while also evaluating the economic cost of the resulting programs.

We model areas in eight provinces of the DRC by calibrating the transmission intensity of the selected areas to malaria prevalence estimates from open data sources, accounting for existing intervention coverage, and using local rainfall and temperature to drive seasonality in vector abundance. In each selected province, we determine effective release strategies of gene drive mosquitoes and define parameter regimes of a sex-ratio distorting suppressive gene drive system, the driving-Y system, that result in elimination of malaria. In the driving-Y system, the process of shredding the male’s X chromosome results in male-biased progeny as the Y- chromosome can still be carried through sperm unaffected to be driven to the next generation (26). The system leads to fecundity reduction, the reduction of the potential to produce offspring, that affects the egg batch size and has implications for the success of the driving system (27,28). We simulate various intervention scenarios, including both conventional and gene drive approaches to vector control, identify combinations of interventions that lead to malaria elimination, and use modeled predictions of malaria burden to estimate DALYs averted and compare the cost-effectiveness of driving-Y gene drives and existing vector control interventions in the DRC.

## Results

This study uses mathematical modeling to explore the potential role of driving-Y gene drives for malaria control and elimination in the DRC. Models were parameterized to capture malaria transmission in eight provinces that span the range of transmission seasonality and intensity across the DRC. Releasing gene drive mosquitoes lowers parasite prevalence in all modeled locations regardless of transmission intensity but is especially effective in higher-transmission areas (Figure 2, Supplementary 1). Tripling the number of gene drive mosquitoes released from 100 to 300 resulted in slightly lower wildtype vector fraction at similar parasite prevalence reduction. In most locations, when compared to three releases of 100 gene drive mosquitoes with one-year interval between each release, a single release of 300 gene drive mosquitoes decreases wildtype vector fraction slightly faster, resulting in faster reduction of parasite prevalence.

**Figure 1.**
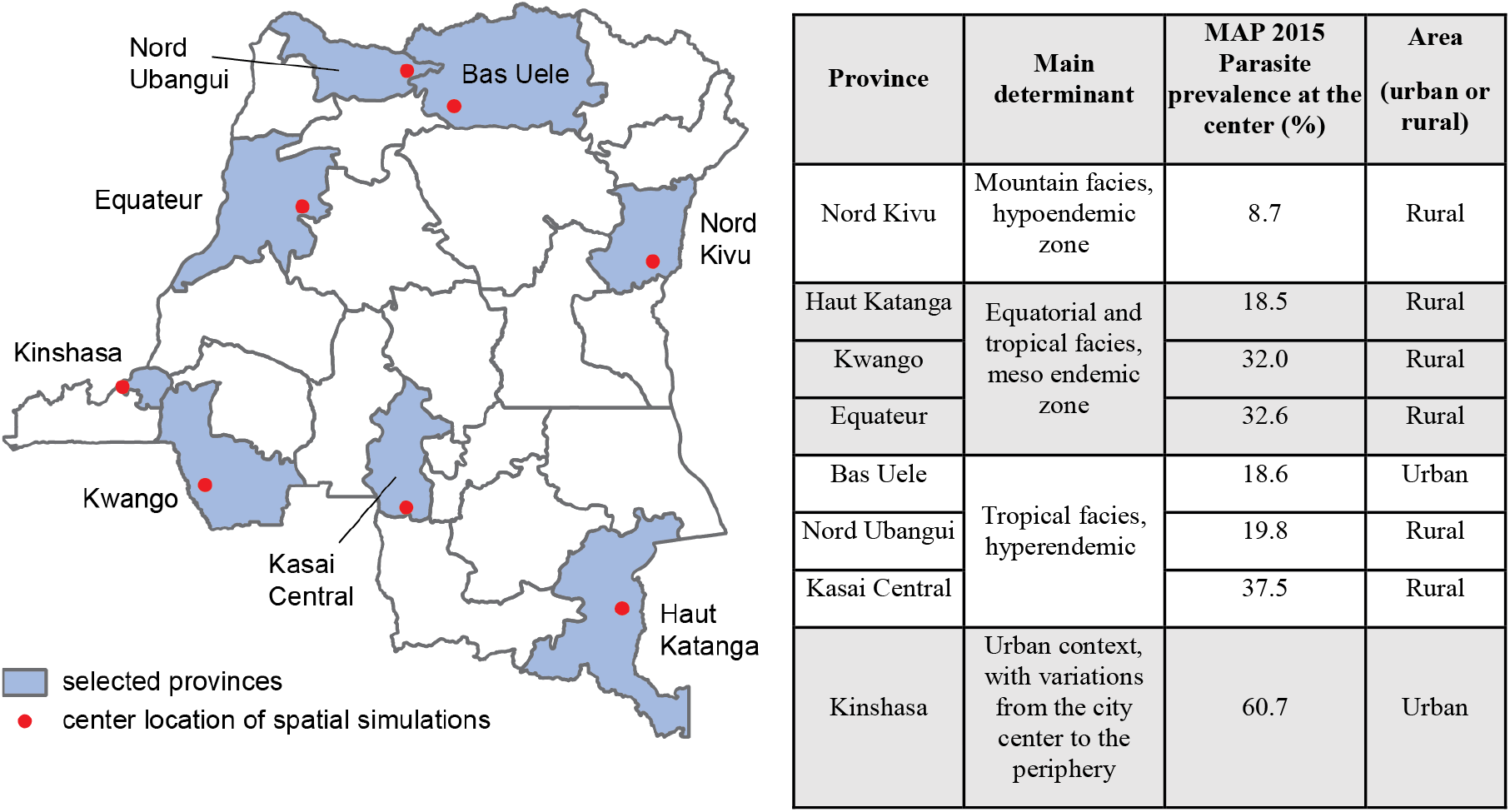
Locations of central nodes of the 25×25km simulation areas. The table describes the main determinants and rural/urban classification of the areas, and Malaria Atlas Project (MAP) estimates of the 2015 parasite prevalence at the central nodes used in calibration in this study.

**Figure 2.**
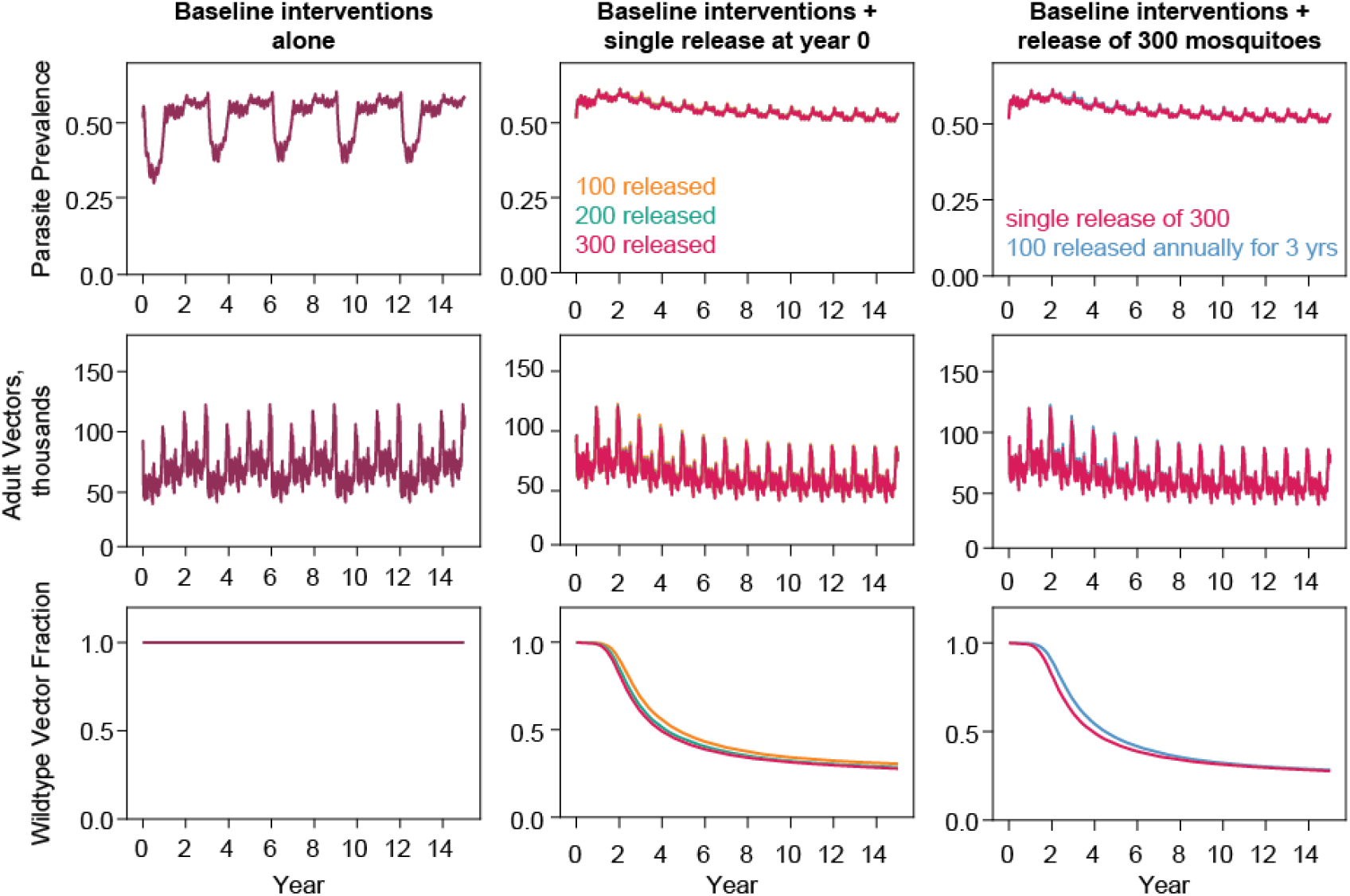
Characteristic non-spatial model output time series from testing a single release of 100, 200, and 300 driving-Y gene drive mosquitoes at year 0 and multiple releases of 100 gene drives at year 0, year 1, and year 2. In the simulations, the X-shredding rate, which is the shredding rate of the X chromosome that favors unaffected Y-bearing sperm ranging from 0.5 to 1.0. The fecundity reduction is the reduction of the potential to reproduce offspring ranging from 0 to 0.5. Results from Equateur site presented here, see Supplementary 1, non-spatial simulation framework: number and frequency of driving-Y gene drive mosquitoes released, for remaining sites.

Successful drives were those with very efficient X-shredding at little to no cost in fertility. Across settings, gene drive was most successful at reducing malaria prevalence when the X-shredding rate ranged from 0.95 to 1.0, and fecundity reduction ranged from 0 to 0.15. The similar sensitivity of mosquito population suppression to X-shredding efficiency could be observed in previous studies using simpler models that studied mosquito population dynamics in a homogeneous and constant environment (29,30) and an extended model that applied regional heterogeneity to model malaria mosquitoes at national scale(15). In our study, the release of drives reduced parasite prevalence, adult vectors, and wildtype vector fraction with a similar trend across all sites (Figure 3, Supplementary 2). Our results also show that in areas where parasite prevalence is high, more efficient drives are required. This hindrance of suppression from possible higher productivity of wildtype population was also reported in previous modeling work (15). Furthermore, our study compared the allowable parameter regimes between fecundity reduction and X-shredding rate that had not been investigated in the previous studies (15,29,30).

**Figure 3.**
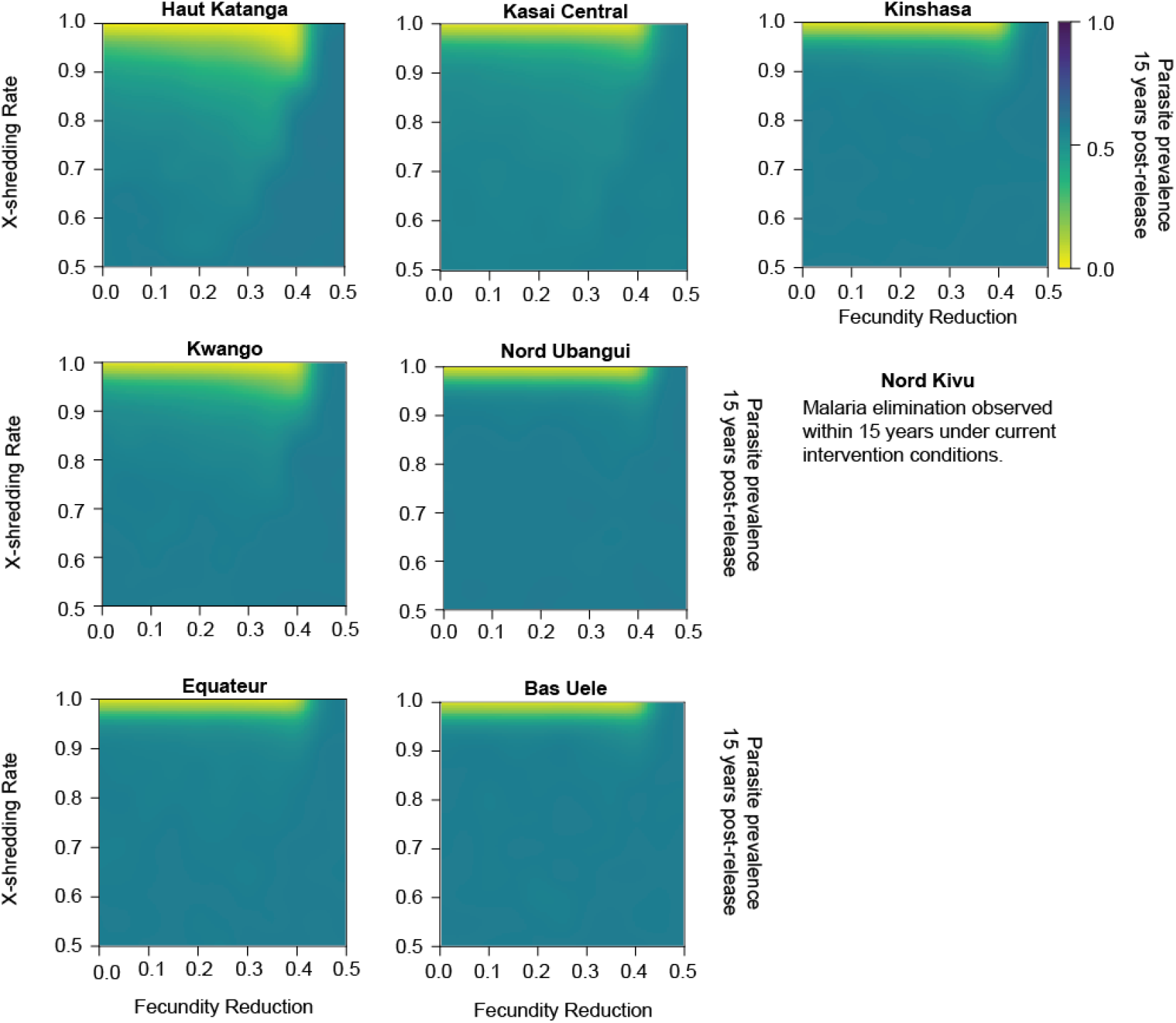
Simulation outputs, observed at the end of 15-year simulation timeframe, post-release of 300 gene drive mosquitoes at year 0 in the non-spatial framework of eight study locations by driving-Y parameter values, see Supplementary 2, non-spatial simulation framework: driving-Y parameters of gene drive mosquitoes released, for post-releases of 100 and 200 gene drive mosquitoes.

The prevalence reduction aligns with the reduction in numbers of adult vectors. In high transmission intensity sites, only drives with very high X-shredding rates (≥ 0.95) eliminated malaria. In lower-transmission settings, drives with slightly lower X-shredding rates (0.9 to 0.95) could also eliminate malaria. These parameter ranges resulted in elimination of the vector population. At X-shredding rates where elimination was not observed, the impact on adult vector population size was larger than the impact on parasite prevalence (Supplementary 3). For spatial simulations, we selected a release strategy of a single release of 300 gene drive mosquitoes with fecundity reduction between 0 and 0.15 and X-shredding rate between 0.9 and 1.0.

Under baseline conditions where interventions remain at constant levels, malaria elimination could not be achieved unless the area has very low transmission intensity (Supplementary 1), which is the case for North Kivu. In the lower transmission intensity areas, malaria elimination could be achieved without gene drives when applying at least 80% coverage of both ITNs and ACT. Regardless of transmission intensity, malaria elimination could be achieved by applying gene drives with an X-shredding rate of 1.0 in all simulated settings. In the spatial simulation framework, in which each site is modeled as 25 interconnected areas, and gene drive mosquitoes are released only in the center of the region, we simulated a range of intervention mixes, using the scenario modeled with current coverage of ITNs and ACT as the baseline comparator. Malaria elimination was achievable without gene drive mosquitoes by combining high coverage of both ITNs and ACT in Haut Katanga (80% coverage of both). In contrast, elimination was not achievable in Kwango, Nord Ubangui, and Kasai Central at these coverage levels, showing the need for new tools and echoing conclusions of the Lancet Commission on Malaria Eradication (31) and WHO’s Strategic Advisory Group on Malaria Eradication (32). For all remaining selected areas with moderate to high parasite prevalence (18.6%, 32.6%, and 60.7% in Bas Uele, Equateur, and Kinshasa provinces accordingly) a single release of single species 300 driving-Y mosquitoes with an X-shredding rate of 1.0 and fecundity reduction between 0.05 and 0.15 eliminated malaria within 15 years (Table 2). In the simulations, we assumed all *P. falciparum* parasite inclusively transmitted by *An. gambiae* as this single species dominates transmission in the DRC. However, results are generalizable to other species or multi-species systems if multiple species-specific drives are released.

**Table 1.** Estimates of costs of interventions per year per one million population applied in the study.

| Interventions | | Coverage (%) | Cost per year (\$int, millions) per one million population [i.e. cost per capita] using 2000 base year | |
| --- | --- | --- | --- | --- |
| Scenarios without gene drives | Insecticide-treated bed nets (ITNs) | 50 | 0.47 |  |
|  | Insecticide-treated bed nets (ITNs) | 80 | 0.63 |  |
|  | Insecticide-treated bed nets (ITNs) | 95 | 0.71 |  |
|  | Case management with artemisinin-based combination therapy (ACT) | 50 | 0.19 |  |
|  | Case management with artemisinin-based combination therapy (ACT) | 80 | 0.20 |  |
|  | Case management with artemisinin-based combination therapy (ACT) | 95 | 0.21 |  |
|  | Combination (ITNs & ACT) | 50 | 0.68 |  |
|  | Combination (ITNs & ACT) | 80 | 0.82 |  |
|  | Combination (ITNs & ACT) | 95 | 0.74 |  |
|  |  |  | Lower bound | Upper bound |
| Scenarios with gene drives | 300 gene drive mosquitoes with X-shredding rates = 1.0 alone | NA | 0.72 | 7.17 |
|  | ITNs plus gene drives with X-shredding rates = 0.95 and 1.0 | 80 | 1.35 | 7.80 |
|  | ITNs plus gene drives with X-shredding rates = 0.95 and 1.0 | 95 | 1.43 | 7.88 |
|  | ACT plus gene drives with X-shredding rates = 0.95 and 1.0 | 95 | 0.93 | 7.38 |
|  | ITNs+ACT plus gene drives with X-shredding rates = 0.95 and 1.0 | 50 | 1.40 | 7.85 |
|  | ITNs+ACT plus gene drives with X-shredding rates = 0.95 and 1.0 | 80 | 1.54 | 7.99 |
|  | ITNs+ACT plus gene drives with X-shredding rates = 0.9, 0.95 and 1.0 | 95 | 1.46 | 7.91 |

**Table 2.**
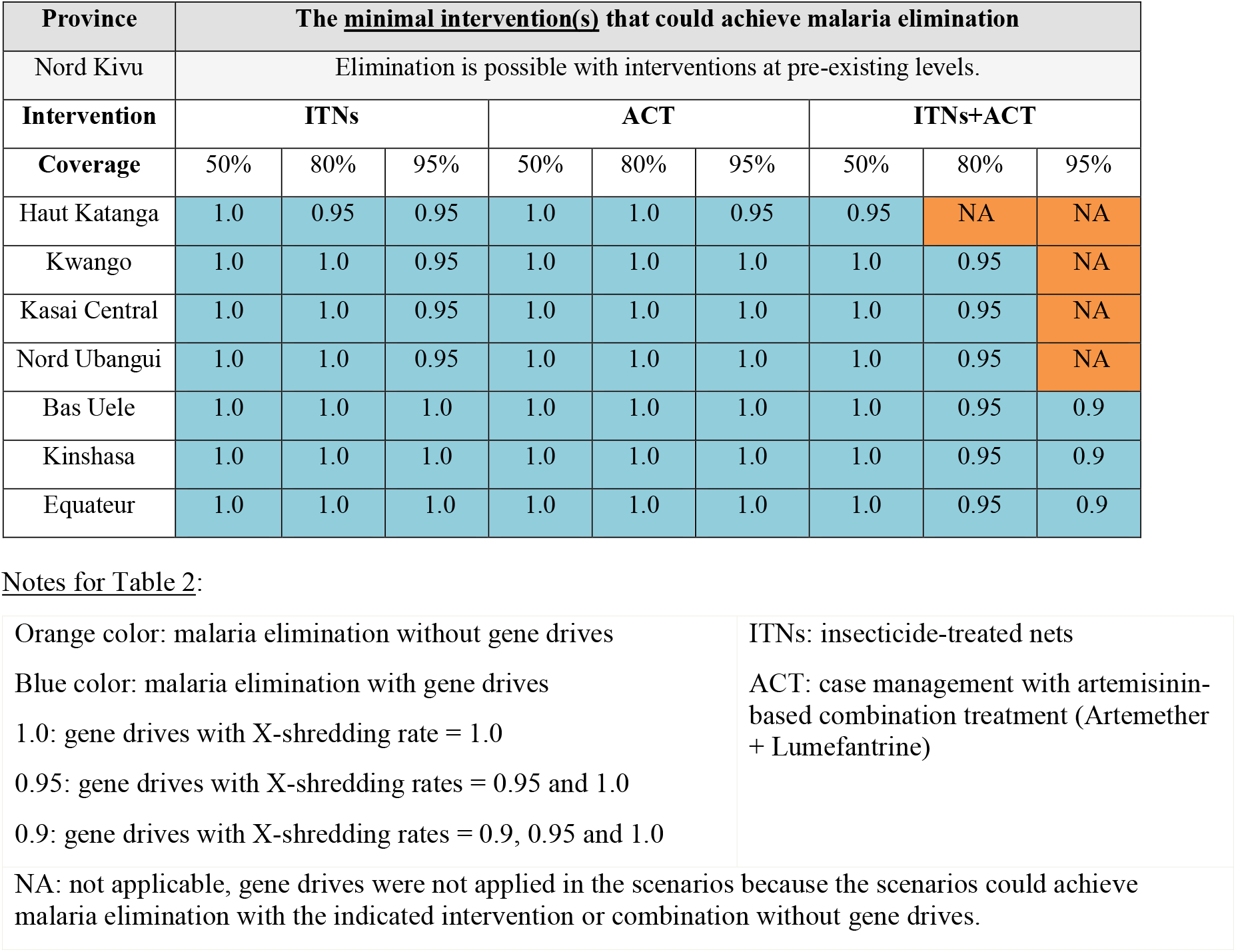
Minimum intervention or combination that can achieve malaria elimination in each target location within 15 years after adding driving-Y mosquitoes into the simulated scenarios. When gene drives were applied, multiple X-shredding rates were simulated. The X-shredding rate in the table is the lowest X-shredding rate that could result in malaria elimination.

| Province | The <u>minimal intervention(s)</u> that could achieve malaria elimination |  |  |  |  |  |  |  |  |
| --- | --- | --- | --- | --- | --- | --- | --- | --- | --- |
| Nord Kivu | Elimination is possible with interventions at pre-existing levels. |  |  |  |  |  |  |  |  |
| Intervention | ITNs |  |  | ACT |  |  | ITNs+ACT |  |  |
| Coverage | 50% | 80% | 95% | 50% | 80% | 95% | 50% | 80% | 95% |
| Haut Katanga | 1.0 | 0.95 | 0.95 | 1.0 | 1.0 | 0.95 | 0.95 | NA | NA |
| Kwango | 1.0 | 1.0 | 0.95 | 1.0 | 1.0 | 1.0 | 1.0 | 0.95 | NA |
| Kasai Central | 1.0 | 1.0 | 0.95 | 1.0 | 1.0 | 1.0 | 1.0 | 0.95 | NA |
| Nord Ubangui | 1.0 | 1.0 | 0.95 | 1.0 | 1.0 | 1.0 | 1.0 | 0.95 | NA |
| Bas Uele | 1.0 | 1.0 | 1.0 | 1.0 | 1.0 | 1.0 | 1.0 | 0.95 | 0.9 |
| Kinshasa | 1.0 | 1.0 | 1.0 | 1.0 | 1.0 | 1.0 | 1.0 | 0.95 | 0.9 |
| Equateur | 1.0 | 1.0 | 1.0 | 1.0 | 1.0 | 1.0 | 1.0 | 0.95 | 0.9 |

Notes for Table 2:

|  |  |
| --- | --- |
| Orange color: malaria elimination without gene drives | ITNs: insecticide-treated nets |
| Blue color: malaria elimination with gene drives | ACT: case management with artemisinin-based combination treatment (Artemether + Lumefantrine) |
| 1.0: gene drives with X-shredding rate = 1.0 |  |
| 0.95: gene drives with X-shredding rates = 0.95 and 1.0 |  |
| 0.9: gene drives with X-shredding rates = 0.9, 0.95 and 1.0 |  |
| NA: not applicable, gene drives were not applied in the scenarios because the scenarios could achieve malaria elimination with the indicated intervention or combination without gene drives. |  |

Reduction in transmission intensity was more sensitive to changes in the X-shredding rate than fecundity reduction in the spatial setting when we compared the interventions with and without gene drives to the baseline (Supplementary 3). With gene drive mosquitoes, the models predicted that malaria elimination could be achieved within 7 years, and in many of these scenarios, it was achieved 4 years post-release.

DALYs averted estimated from the model’s outputs show similar trends as those of WHO (Table S1 in Supplementary 6). Population-level cost-effectiveness estimates for individual and combined interventions as costs per DALY averted in comparison with the baseline scenario indicate that DALYs averted, rather than cost, is the main factor determining cost-effectiveness across interventions (Table S2 in Supplementary 6). In scenarios with gene drives that resulted in malaria elimination, the costs per DALYs averted are lower in areas where the transmission intensity is initially higher as there were more DALYs to avert in the high transmission area with comparable costs between scenarios and the costs decrease over time as DALYs continue to be averted after elimination is achieved (Table 3).

**Table 3.**
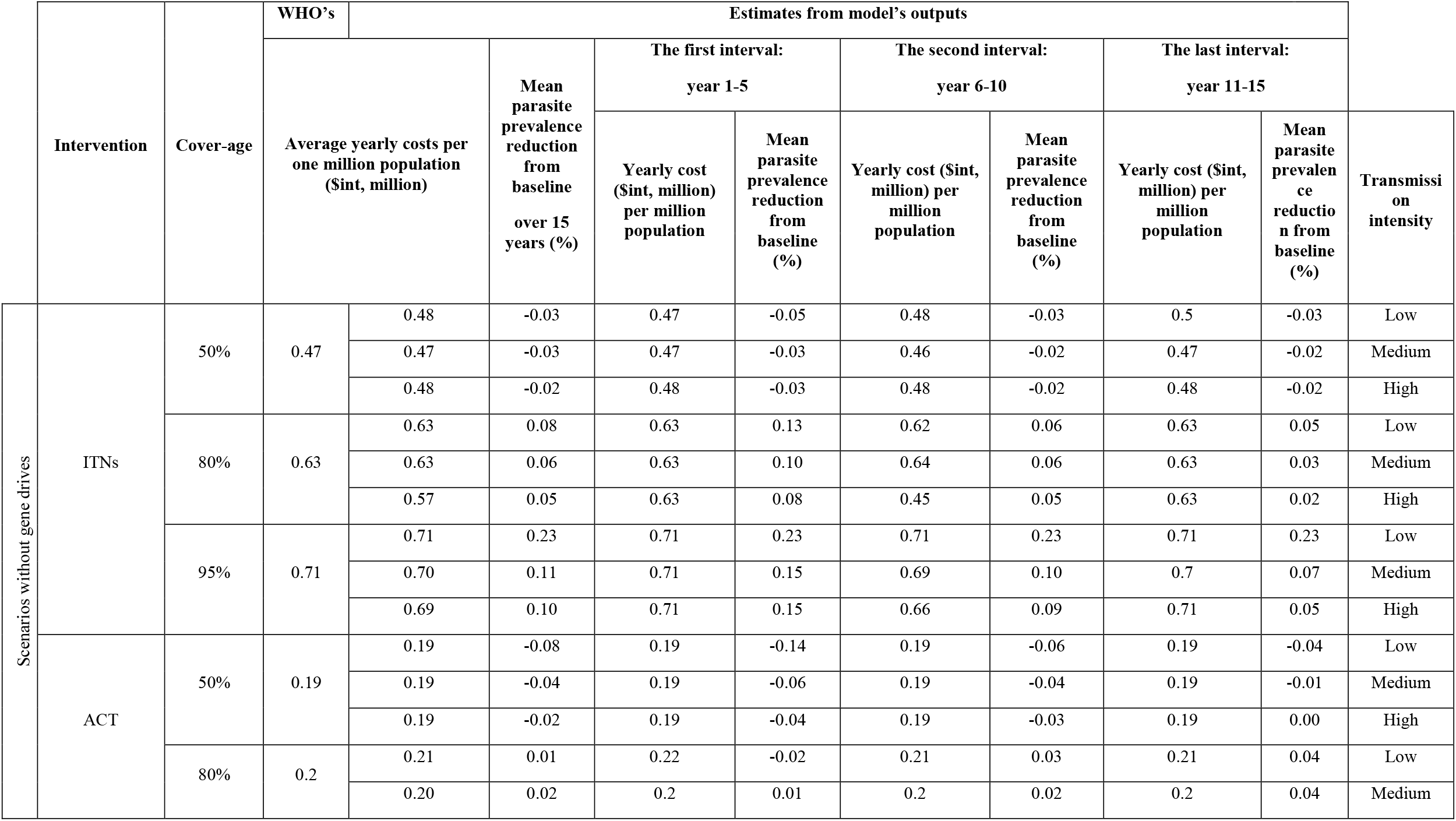
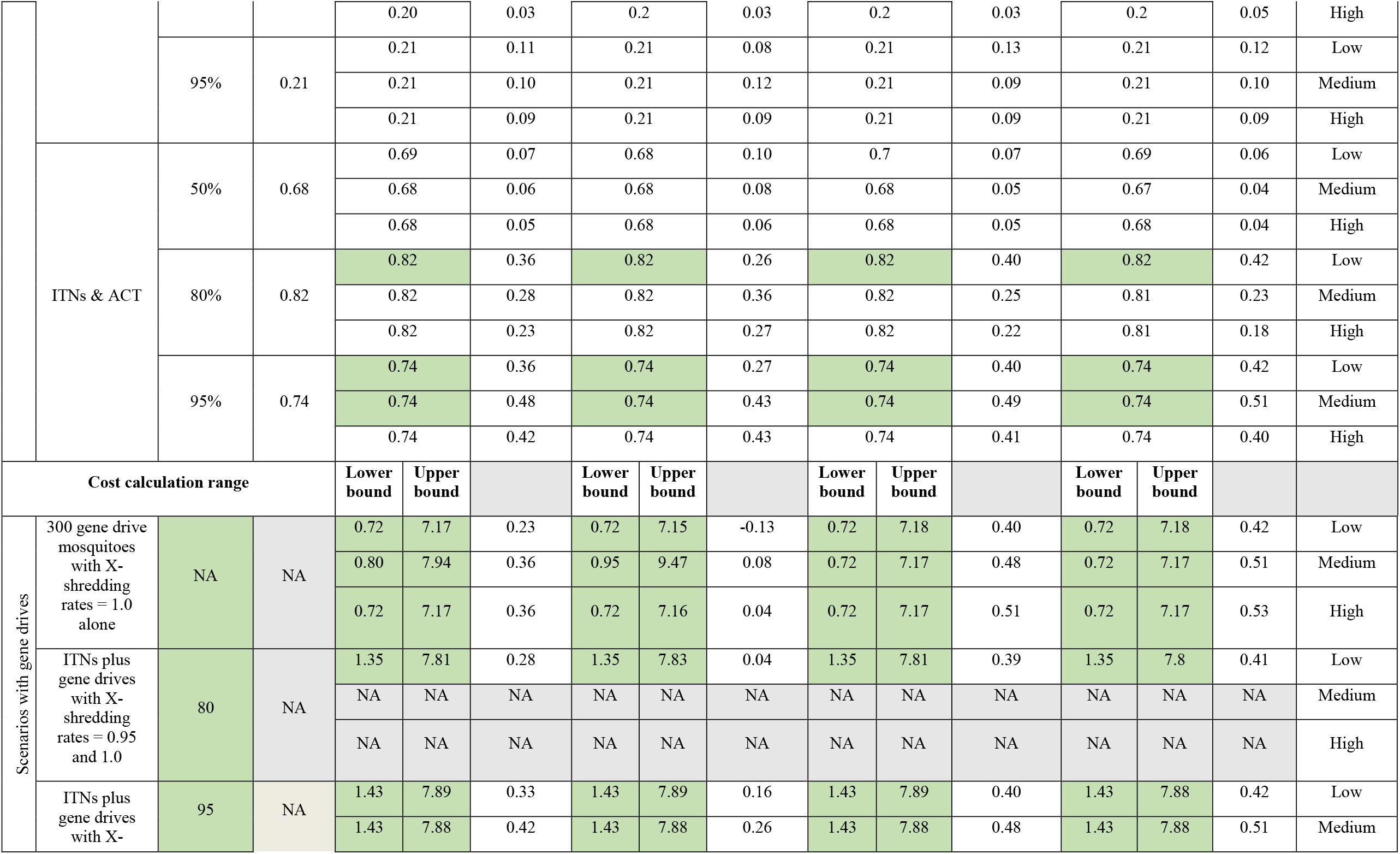
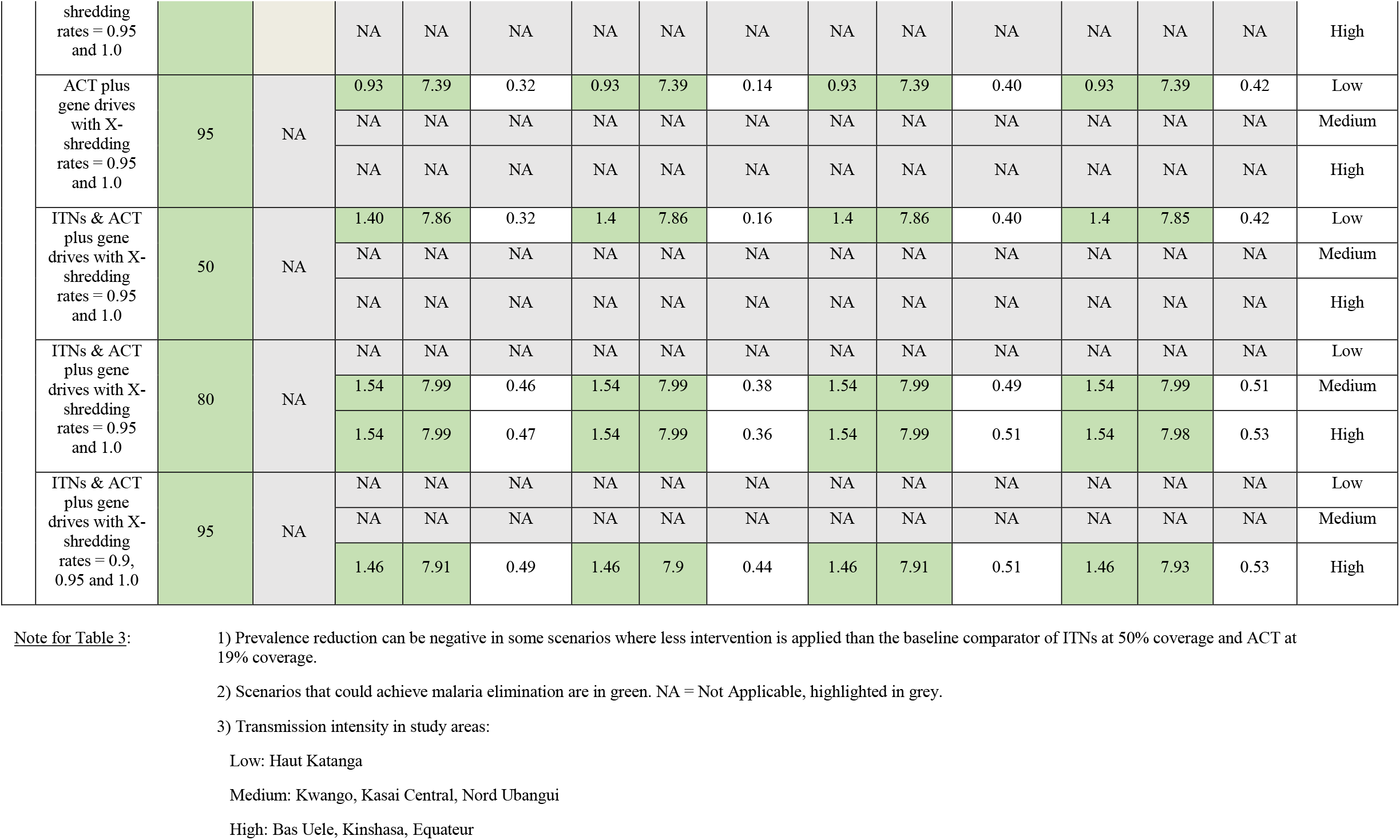
Average yearly cost per one million population and mean parasite prevalence reduction for interventions and combinations of interventions applied in the study.

The expansion paths of all sites show the order in which interventions would be selected at different levels of resources available based on the incremental cost-effectiveness ratio (ICER). ICER indicates additional costs required to avert each additional DALY by moving from the lower-cost to the higher-cost intervention (33). It is calculated using average yearly costs and yearly effectiveness (Table 3). Notable differences exist between the first and the following two 5-year intervals. The combination of ITNs and ACT at 95% coverage is the most cost-effective intervention plan. However, it is not clear that 95% coverage of either ACT or ITNs as single interventions or in combination would be achievable under the estimated costs, as even with high expenditures, such levels of coverage have not yet been achieved (31,34). For the first interval, the high-coverage combinations of ITNs and ACT are more cost-effective (Figure 4 and Table S3 in Supplementary 6). In the following years (second and third intervals; Table S3 in Supplementary 6), the unit cost of gene drive mosquitoes affects the priority of the strategies on the expansion path as gene drives become more cost-effective compared to other interventions (Figure 4).

**Figure 4.**
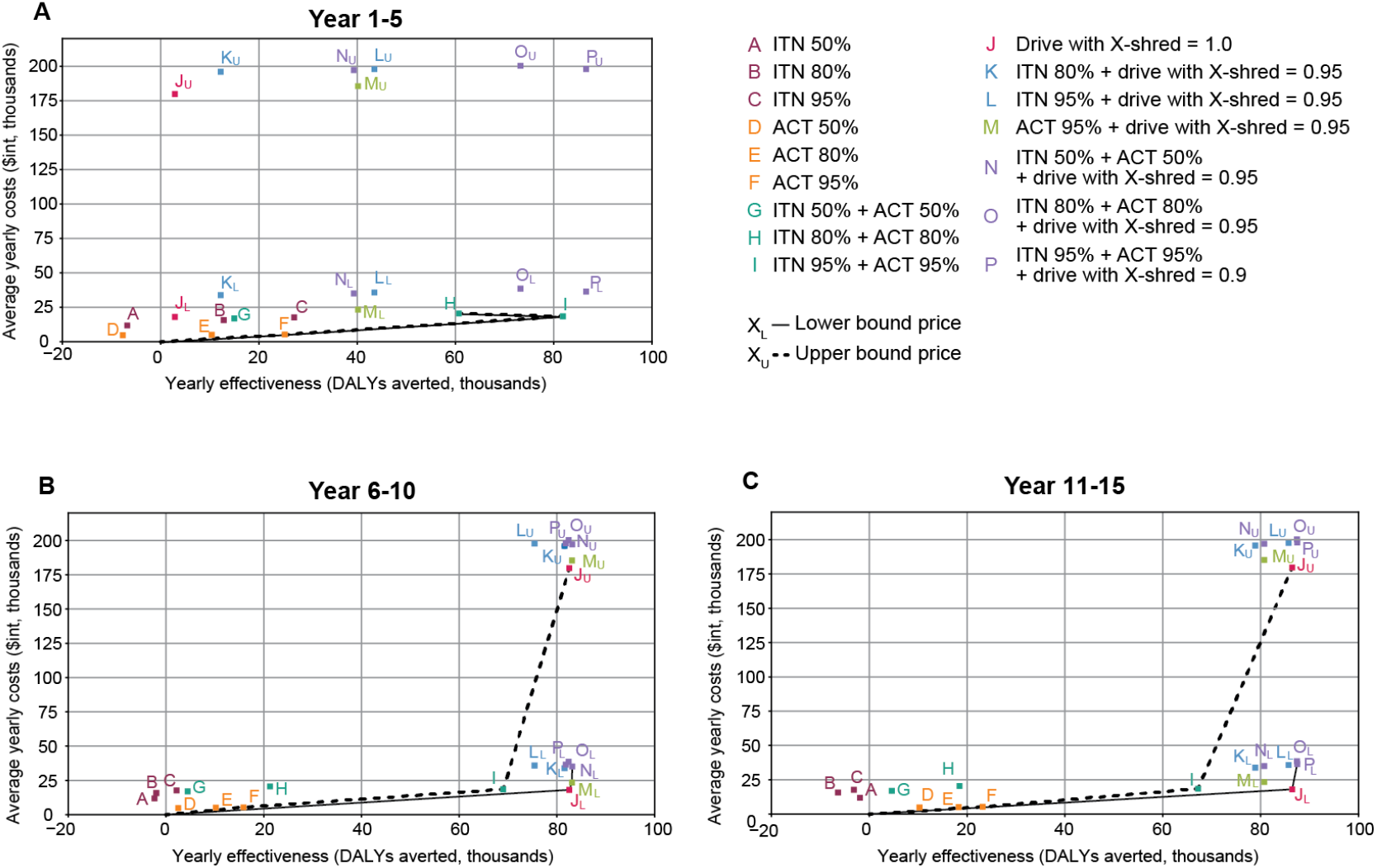
Cost-effectiveness plane showing 16 intervention packages (10 individual and combination interventions at three assumed coverage levels) and expansion paths for year 1-5, year 6-10, and year 11-15. The black lines (X_L_) are expansion paths based on the lower bound and the dashed lines (X_U_) are expansion paths based on the upper bound costs of gene drives.

The cost of gene drives as a single intervention in this study calculated in International Dollar ($int), a hypothetical unit of currency that has the same purchasing power that the U.S. Dollar (US$) has in the United States at a given point in time (33), ranged from 0.72 $int to 7.17 $int per person per year (Table 1). The calculation is based on the costs of gene drives per person in previous studies (35,36). In case of combinations, the cost of gene drives was added to the cost of other interventions. Using the lower bound price for the cost of gene-edited mosquitoes, gene drive as a single intervention is the most cost-effective intervention overall as gene drive mosquitoes with an X-shredding rate of 1.0 could eliminate malaria in all contexts and would be the first choice where resources are limited. The effect, however, could be seen after elimination was achieved from the second 5-year interval onwards. If malaria elimination cannot be achieved, 95% ACT coverage is the most cost-effective intervention. The result suggests that, if gene drives with 100% effective construct, an X-shredding rate of 1.0 and fecundity reduction of 0.05-0.15 are available, and their cost is comparable to other existing interventions, gene drives would be the most cost-effective single intervention for malaria elimination. It is possible to maintain existing interventions, especially ACT and ITNs, early on, while gene drives are propagating. Once the mosquito population collapse, gene drives become more cost-effective over a medium timeframe.

## Discussion

This study uses mathematical modeling to describe the potential role of sex-ratio distorting gene drive mosquitoes in malaria control across the transmission spectrum in the DRC, an area where achieving effective control has historically been challenging. Our results suggest that population suppression through gene drives could be an effective strategy for malaria elimination in the DRC, either as a single intervention or in combination with other interventions. To the best of our knowledge, this is the first study that models the epidemiological impact and cost-effectiveness of gene drive mosquitoes for malaria elimination. Previous studies involving gene drives for malaria control are limited in scope to laboratory experiments (37–40), and the development and parameterization of mathematical models (15,41–43). By extending previous modeling work (44) to approximately estimate the cost-effectiveness of gene drive in representative settings, our work helps fill a gap in evidence about the programmatic implementation of gene drives in the context of limited resources. The study not only estimates the feasibility of gene drives in realistic malaria elimination scenarios, but also evaluates cost of gene drives in comparison to other currently available interventions. This work helps gauge the probabilities of success and possible outcomes of gene drives that are strictly laboratory-contained or in the transition from the laboratory-based research to future field-based research. In addition to the technical perspective provided in this study, further work is necessary, including on the ethical perspective, i.e., standard research ethics, procedural ethics, and participatory management of the technology (45), as a key component to implement this technology in wild mosquito populations (46).

We found that the success of driving-Y gene drives in all areas regardless of vector density highly depends on the ability of gene drives to shred the X chromosome. Though a naturally occurring driving-Y chromosome that transmits >90% of male progeny can be found in *Aedes* and *Culex* mosquitoes (47) and a CRISPR-based X-shredder can generate up to 100% male bias in the laboratory (48,49), driving-Y gene drives have yet to be developed in the laboratory. The adoption of a driving-Y strategy could be very challenging because it may be difficult to achieve a perfect X-shredding rate at every development stage and during implementation while overcoming the challenge of meiotic sex chromosome inactivation (45). Moreover, possible resistant mutants that could convert wildtype genes and spread resistance, especially in *An. gambiae* that cleavage resistant alleles, have already been observed (48).

Some sex-ratio distorting drives may not be comparable to Y drive or X shredders: for example, if the female carries a drive that inactivates the reproduction of her male progeny. However, the difference in mechanism by which sex ratio distortion is achieved may only lead to differences in how quickly a drive establishes itself rather than downstream outcomes regarding elimination, which is the focus of this study. Our finding that the drive must be highly effective while mild fecundity costs are well-tolerated is likely generalizable to other suppression drives as a whole. Our economic findings on the cost-effectiveness of drives are likely to be order-of-magnitude similar for any highly effective suppression drive.

The success of suppressive gene drives such as driving-Y relies on mosquito population size and allowing enough time for the drives to propagate in the mosquito population. Hence, understanding interactions between existing vector control methods such as ITNs and IRS that temporarily reduce the mosquito population (36,50) and gene drives will be necessary given that vector control typically reduces mosquito populations. While this work focuses on the impact of vector abundance, seasonality, and conventional vector control on gene drive outcomes, spatial connectivity of mosquito populations and terrain heterogeneity will also have important implications for gene drives success. Accurate capture of local variation in mosquito population connectivity requires data on mosquito swarms and habitats at high resolution, mosquito movement patterns, and mosquito species introgression. Some of these quantities are measurable and known but most are unavailable for the DRC. A sensitivity analysis with vector migration rate reduced by up to 3 orders of magnitude did not observe any change in elimination outcomes (Supplementary 5).

While our models predicted that high coverage with ITNs and high access to treatment with ACTs could eliminate malaria in lower-transmission settings, achieving such high coverages of existing measures is not only extremely difficult but also comes with high implementation and logistical costs (51,52). It may take much more investment in logistics and systems to achieve 95% coverage of both ITNs and ACT than WHO’s estimates applied in the study (Table 1) (34). Even if theoretically achievable, it is highly improbable to sustain necessary coverage levels in the complex operational environment of high disease burden countries like the DRC (53,54). The costs estimated in the previous WHO study and applied in this work considered program level, patient level and opportunity costs of currently available interventions in the respective WHO subregion. These estimated costs might still underestimate the costs of existing interventions given the difficulty for widespread distribution of interventions in the DRC (55,56). Considering the potential percolation properties of gene drives, especially in areas where mosquito populations are more connected, successful gene drives could plausibly be more cost-effective if the cost is comparable to currently available interventions. However, this may not be the case for areas with more sparsely connected mosquito populations and higher rates of gene pool variability. Other cost components, e.g., surveillance, will also contribute to the cost of gene drive once implemented.

Our model results show that tailoring the frequency of releases and the number of gene drive mosquitoes to be released can make malaria elimination achievable within 5 years after a single release of gene drive mosquitoes under certain conditions, including but not limited to no importation of vectors or infections. The importation can sustain transmission and cause resurgences (57). Therefore, future work is necessary to include importation of vectors and infections to address the feasibility of release, as well as specify release schedules that are operationally practical and technically necessary for intended deployment areas. Because of their self-propagating and self-sustaining properties (26), gene drives would likely result in better cost-effectiveness once implemented compared to other genetically engineered mosquitoes previously developed (e.g., SIT). Nonetheless, the payoffs can be observed once malaria elimination is reached - in most cases, after 5-year post-release. This waiting period can be critical, given many life losses in the interim in the DRC’s context. We based our cost-effectiveness analysis on the unit costs of OX513A (58) and *Wolbachia* infected mosquitoes per person (59) based on available information considering the limited cost data of genetic control methods. We performed a systematic scoping review (Supplementary 7) to search for evidence in costs of vector control approaches that involve the release of mosquitoes to modify vector population and selected moderate lower and upper bound costs to give approximate values. The rationale to apply the cost of gene drives per person protected instead of using the cost per gene drive mosquitoes is to be conservative in approaching the cost estimation given the low number of gene drive mosquitoes released in the models in this study.

The lower bound cost applied in this study is from a study on *Wolbachia* infected mosquitoes, where costs came from expenses for the entire deployment (60). These estimates are somewhat uncertain because future deployments would likely utilize less monitoring in an operational public health program than in research and occur in settings of higher population density. The self-maintaining of *Wolbachia* post-deployment would also reduce ongoing costs. The upper bound cost is from studies on the Oxitec mosquito (59,61). One study source did not explicitly state the cost components (59) and the other modeled estimated costs based on preliminary estimates by Oxitec (61). Thus, different situations and contexts may naturally lead to deviation in costs from these estimates. The use of Oxitec mosquitoes requires surveillance efforts and recurrent re-licensing from the patent-owning company, which could further increase costs. The lower and upper bound range in costs applied in the study reflects the reality in the field, as the genetic control methods vary in cost components even though the methods were developed to tackle the same disease (53). Future research should explore the cost components of gene drives, especially development, and environmental costs, that may contribute to changes in overall cost of this type of intervention. The changes in unit cost could affect the cost-effectiveness of the method if the cost is too high. As we demonstrated in the incremental cost-effectiveness ratio analysis, the cost-effectiveness is cost-sensitive. The gene drive approach becomes less cost-effective compared to other strategies once its cost increases.

Gene drive technology is at an early stage of development and concerns over ethics, safety and governance, as well as questions on affordability and cost-effectiveness, must be addressed prior to implementation (62). For high malaria burden countries such as the DRC, collaborating on testing, implementing and regulating new technologies like gene drives poses challenges not only from within the country but also with other countries where different systems of governance can further complicate the collaborations (63). Presently, many countries including the DRC have insufficient resources to individually follow recommendations such as extensive risk assessment and safety testing, and close monitoring after mosquito releases (62), making it a challenge to enforce legislation required under the Cartagena Protocol (64). Further research on risk assessments and step-wise implementations may help reduce uncertainties and characterize potential risks and benefits that involve crucial ethical and social challenges (14,65).

This study demonstrated a modeling approach applied to *An. gambiae*, the predominant malaria vector in Africa (66). The methodology developed here can be applied to other malaria-transmitting mosquito species, and in settings with multiple major malaria vectors, releasing an equally effective gene drive for each major vector would approximate the impact of targeting *An. gambiae* in this study. The study identified key aspects of both gene drive technology and its implementation that are fundamental for the technology to be a cost-effective component of a malaria control program. Amid uncertainty about vector abundance and its behavior (67) and no importation of infections and wildtype mosquitoes in the models, the study offers an evaluation framework. The framework can be generalized to look at other gene drive approaches to effectively plan gene drive strategies in malaria control, especially in other high burden countries where parasite transmission intensity varies.

## Materials and methods

The simulations in this study use Epidemiological MODeling software (EMOD) v2.18 (68), an agent-based, discrete-time, Monte Carlo simulator of malaria transmission with a vector life cycle (69) and within-host parasite and immune dynamics (70,71). The modeling framework combines an epidemiological model of *Plasmodium falciparum* transmission between individual human agents and cohorts of mosquito agents distinguished by life stage, feeding and oviposition stage, age, and genotype. The vector lifecycle consists of four stages: egg, larvae, immature adults, and host-seeking adults, with temperature-dependent larval development, immature maturation, and sporogony. Mosquito abundance is driven by availability of larval habitat, and mosquito mortality is also affected by temperature and humidity. In humans, the model includes asexual parasite and gametocyte densities, human immunity, effects of antimalarial drugs, and symptomatic aspects of malaria, all of which have been previously calibrated to field data (72–74). The model dynamically simulates vector-human and human-vector transmissions during blood meals. Driving-Y is one of several gene drive strategies that can be simulated within EMOD (75).

We selected eight representative provinces in the DRC for simulations (Figure 1) in both non-spatial and spatial simulation frameworks. These approaches differ in whether or not vector migration is included. The selection was based on malaria parasite prevalence data from the DRC-Demographic and Health Survey (DRC-DHS) II 2013-14 (76,77), Malaria Atlas Project (MAP) parasite prevalence estimates (78,79), and provincial stratification by climate zones, endemicity, and urban/rural (13) to ensure that selected locations spanned the range of transmission intensity across the entire DRC. For each site, simulations were run on a square 25×25 kilometer grid containing 25 nodes, 5 kilometers apart in both the non-spatial framework where no vector migration was present across all nodes and the spatial framework where vector migration was included. The model’s outputs of malaria incidence and mortality were then used to assess the cost-effectiveness of interventions.

We selected a central node for each simulated province by identifying a survey point from the MAP parasite prevalence survey database (79) such that the 25-node simulation area was entirely within the selected province (80) and used WorldPop population estimates (81) to verify that the central node is populated. We applied various environmental covariates specified to each study location based on geolocation. The covariates include climate (rainfall, temperature, humidity) and seasonality averaged from monthly vectorial capacity. Since *An. gambiae* mosquitoes, the only modeled mosquito species, breed primarily in temporary puddles replenished by rainfall and drained through evaporation and infiltration (82), the simulations use climate data to model the availability of larval habitat, which drives the number of vectors throughout the year and thus biting intensity and transmission (69). Weather station and readings by National Oceanic and Atmospheric Administration (NOAA) Global Surface Summary of the Day (GSOD layer) were used for generating temperature and dewpoint anomalies. Baseline monthly averages were generated using WorldClim 1.4 raster files in a grid format, 2.5 arc minutes, and 30 arcseconds from WorldClim 1.4 (83). Rainfall files were generated by downscaling RFE 2.0 Rainfall Estimates from NOAA’s Climate Prediction Center (84). In both non-spatial and spatial simulation frameworks, seasonality was enforced in the models by setting the seasonality of larval habitat abundance such that monthly vectorial capacity matched the average monthly vectorial capacity between 2000 to 2015 in two public datasets (v200906 and Sheffield) (85). Daily temperature series was generated for each node as in (86).

We calibrated the overall larval habitat abundance by scaling the previously fitted seasonality profile such that the model’s parasite prevalence was consistent with the mean 2015 annual parasite prevalence of the location from MAP estimates (87). The annual means of estimated parasite rate in children between the ages of two and ten (PfPR_2-10_) from the year 2000 to 2015 were retrieved from MAP rasters (87) for all simulation nodes. For Haut Katanga, the larval habitat multiplier was calibrated so that the average modeled parasite prevalence for years 2013-2015 was close to the MAP estimates for the same period. This adjustment was made due to the site’s very low parasite prevalence. We set each node’s population to 1,000 individuals and set birth and mortality rates to 36.3 per 1,000 people per year. The human population size used is large enough to sustain low-transmission malaria but not unrealistically large for rural areas. The simulation was run for 50 years to initialize population immunity.

In the final 10 years of the 50-year initialization period, the following interventions were imposed (13):

ITNs: modeled ITN usage was based on % of children under the age of five (<5) who slept under an ITN the previous night: 6%, 38%, and 56% in 2007, 2010, and 2013, respectively.

Case management of symptomatic cases with Artemisinin-based Combination Therapy (ACTs): 19% of uncomplicated malaria cases in all ages received treatment with artemether-lumefantrine, based on the 2013 DHS survey reporting 19% of febrile children under 5 receiving ACT.

Indoor Residual Spraying (IRS) was not included as less than 1% of the DRC population was protected by IRS between 2007 and 2018 (3,88).

After the 50-year initialization, simulations were run for the next 15 years in both non-spatial and spatial simulation frameworks. For ITNs used in the model, the initial strength of the blocking effect on indoor mosquito fed on an individual with an ITN was 0.9, and the blocking decayed at an exponential rate with mean 730 days. The blocking effect captures the physical barrier of a bednet that prevents a mosquito from making contact with a human. The initial strength of the killing effect was 0.6, conditionally on a successfully blocked feeding event. The killing effect decayed at an exponential rate with mean 1,460 days. Killing and blocking parameters were obtained from calibration to clinical trial data (71). The model assumed an individual who received an ITN had 0.65 probability of using it on any given night, and ITNs were redistributed every 3 years. For ACT, the parameters and values used in the model followed (72).

Under the gene drive intervention, females that mate with a male carrying driving-Y will have as offspring wildtype females and males carrying the driving-Y. The fraction of offspring that are driving-Y males is then 0.5+0.5*(X-shredding rate), and the fraction of offspring that are females is 0.5-0.5*(X-shredding rate). Only females that mate with a driving-Y male have their fertility reduced, and the total egg batch size is reduced by the fecundity reduction for each female that mates with a modified male (44). We varied the X-shredding rate from 0.5 to 1.0 and fecundity reduction from 0 to 0.5 for single releases of 300 gene drive mosquitoes. We selected parameter sets of X-shredding rates (0.9, 0.95, 1.0) and fecundity reductions (0.05, 0.1, 0.15) to generate 9 combinations of driving-Y parameters that could eliminate malaria in the non-spatial framework (Supplementary 2).

In the non-spatial simulation framework, we sampled the number of gene drive mosquitoes released, frequency of release, and driving-Y parameters and evaluated whether malaria was eliminated. The range of 100 – 300 mosquitoes to release provided a sufficient number of gene drives to seed successfully in the majority of the simulations, avoiding stochastic die outs that occur at lower release numbers. The release size is yet small enough compared to most typical mosquito population sizes in km^2^ areas, which provides an optimistic costing assumption for successful gene drives. The driving-Y gene drive release strategy and parameters were then applied to the spatial simulation framework, where we identified which interventions or combinations of interventions could eliminate malaria in the selected locations. In all simulated scenarios, the interventions, including gene drive mosquitoes, were applied on the first day of year 0 unless indicated otherwise. A scenario was defined as reaching malaria elimination when all-age parasite prevalence in the model is not detectable, i.e., drops to zero.

In the spatial simulation framework, vectors and humans could each move between adjacent nodes less than 10 km apart with a rate inversely proportional to their distance. Adjacent nodes include diagonally adjacent nodes. The vector migration rate ranged from 0.09 to 0.12 (9-12% of female mosquitoes migrate out of a grid cell in any day). Each human individual went on a multi-step journey along nodes and retrace back along their outbound path once they reached 5 nodes from their home node. On the outbound journey, the duration at each node was not set explicitly but was determined by each node’s migration rate that ranged from 0.09 to 0.12; on the return journey, one timestep is spent at each node.

ITNs at 50% coverage and ACT at 19% coverage were applied uniformly across all nodes in baseline scenarios, which were used as the main comparator against other scenarios. Each of the individual ITNs and ACT (case management rate with ACT) and the combination of ITNs and ACT was analyzed at three levels of coverage: 50%, 80%, and 95%, following the selected coverage levels in (89). Scenarios that failed to eliminate malaria were re-simulated with the addition of a single release of 300 driving-Y gene drive mosquitoes at the central node on the first day of year 0.

We calculated Disability-Adjusted Life Years (DALY) from model outputs – population by age group, uncomplicated clinical cases, severe cases, and deaths – at year 5, year 10, and year 15 by giving equal weights to years of healthy life lost at young ages and older ages and with 0% discount rate for future lost years of a healthy life. The standard life expectancy at the age of death in years, and the DRC’s country lifetable (90), and disability weights (35) of moderate (0.051) and severe (0.133) were applied in DALYs calculation. DALYs averted (91,92) were then calculated by comparing outcomes to those of baseline scenarios.

Costs of all interventions, including gene drive mosquitoes, were calculated standardized to year 2000 without discount rates in order for the results to be comparable to those of previous WHO milestone studies (89,93). Estimated costs are expressed in international dollars ($int) (33). Coverage-dependent costs per person per year of applying ITNs, ACT, and the combination of ITNs and ACT were obtained from WHO-CHOICE database (94) and a previous WHO study (93). For scenarios that included gene drive mosquito releases, we assumed the financial cost of gene drives as a single intervention ranged from 0.72 $int to 7.17 $int per person per year (Table 1) based on costs of gene drives per person in previous studies (59–61) found in a systematic scoping review (Supplementary 7). We applied the US government consumer price index (CPI) to adjust for inflation and the cumulative inflation rate to year 2000 values. Costs of gene drive were added to the costs of any underlying intervention(s) also distributed (Table 1). Cost-effectiveness was calculated for each 5-year interval beginning in 2015 by dividing average yearly costs in $int by average yearly effectiveness in DALYs averted (33). More cost-effective interventions were identified by drawing a graph of an expansion path through ICER, which uses the monetary value to compare the interventions (33), and selecting interventions with more favorable cost-effectiveness. The expansion path is drawn to connect the choices of interventions and present the order that the interventions would be chosen once more resources become available, considering only cost-effectiveness. The additional cost required to avert each additional DALY, ICER, is the slope of each expansion path (33).

## Data Availability

Codes on GitHub:

gene drive: https://github.com/NaniMet/gene-drive

dtk-tools: https://github.com/InstituteforDiseaseModeling/dtk-tools

EMOD: https://github.com/InstituteforDiseaseModeling/EMOD

https://github.com/NaniMet/gene-drive

https://github.com/InstituteforDiseaseModeling/dtk-tools

https://github.com/InstituteforDiseaseModeling/EMOD

## General

The authors thank the Institute for Disease Modeling and, in particular, Benoit Raybaud and Edward Wenger for their generous technical support and resource sharing. We are immensely grateful to Philip Welkhoff and David O’Brochta for helpful discussions and comments on earlier versions of the manuscript.

## Author contributions

Conceptualization and experimental design: NM, JG, CB, JvB, and MN. Experimental work: NM. Data interpretation and analysis: NM, JG, and PS. Supervision: JG, CB, and JvB. Writing original draft: NM and JG. Writing – review and editing: NM, JG, PS, CB, JvB, MN and GA.

## Data and materials availability

Codes on GitHub:

gene drive: https://github.com/NaniMet/gene-drive

dtk-tools: https://github.com/InstituteforDiseaseModeling/dtk-tools

EMOD: https://github.com/InstituteforDiseaseModeling/EMOD

