## Supplementary for "Modeling impact and cost-effectiveness of gene drives for malaria elimination in the Democratic Republic of the Congo"

**<u>Supplementary 1</u>, Non-spatial simulation framework: number and frequency of driving-Y gene-drive mosquitoes released.**

***Baseline observed throughout 15-year simulation timeframe in the spatial framework of eight study locations:***

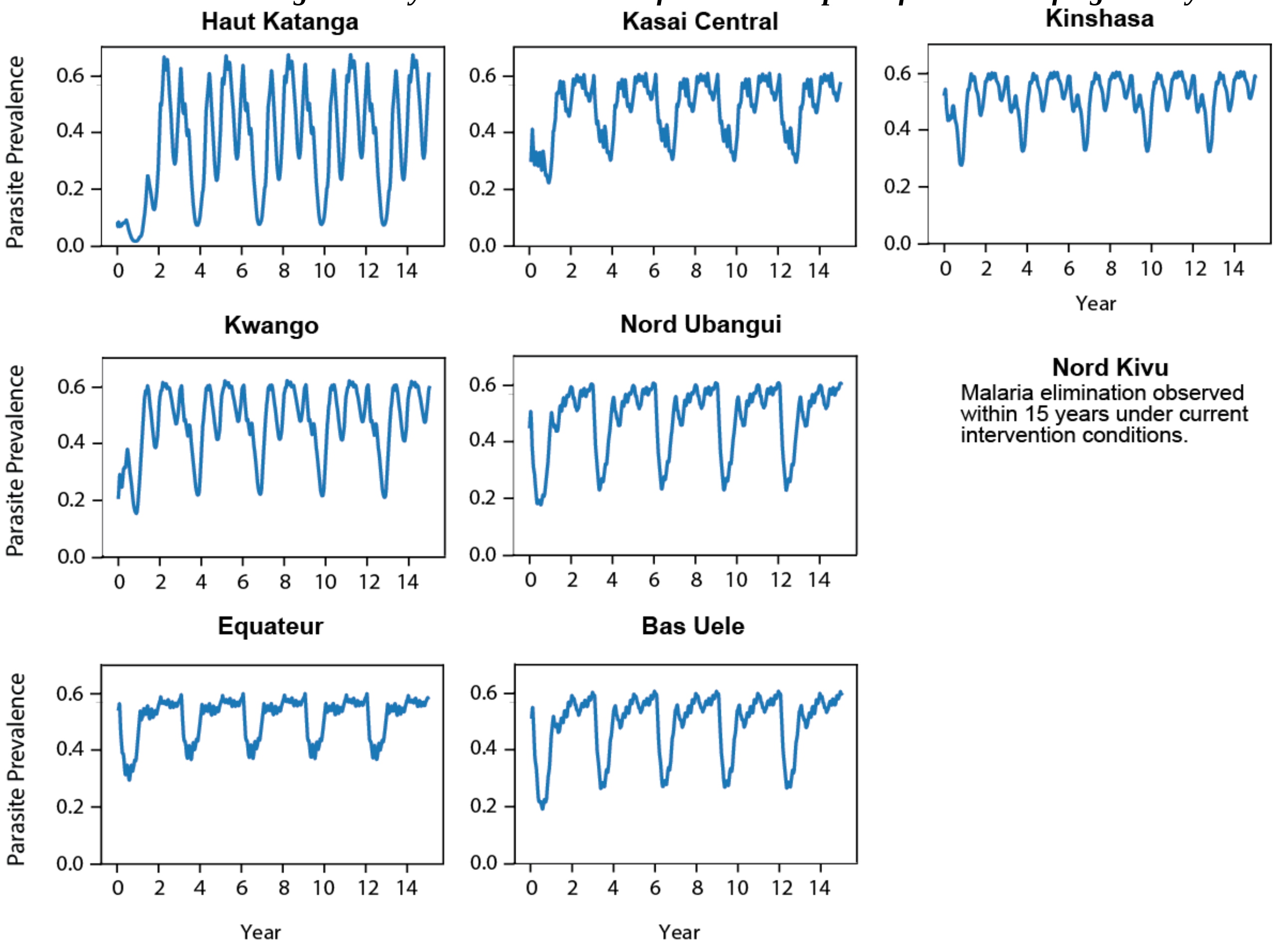

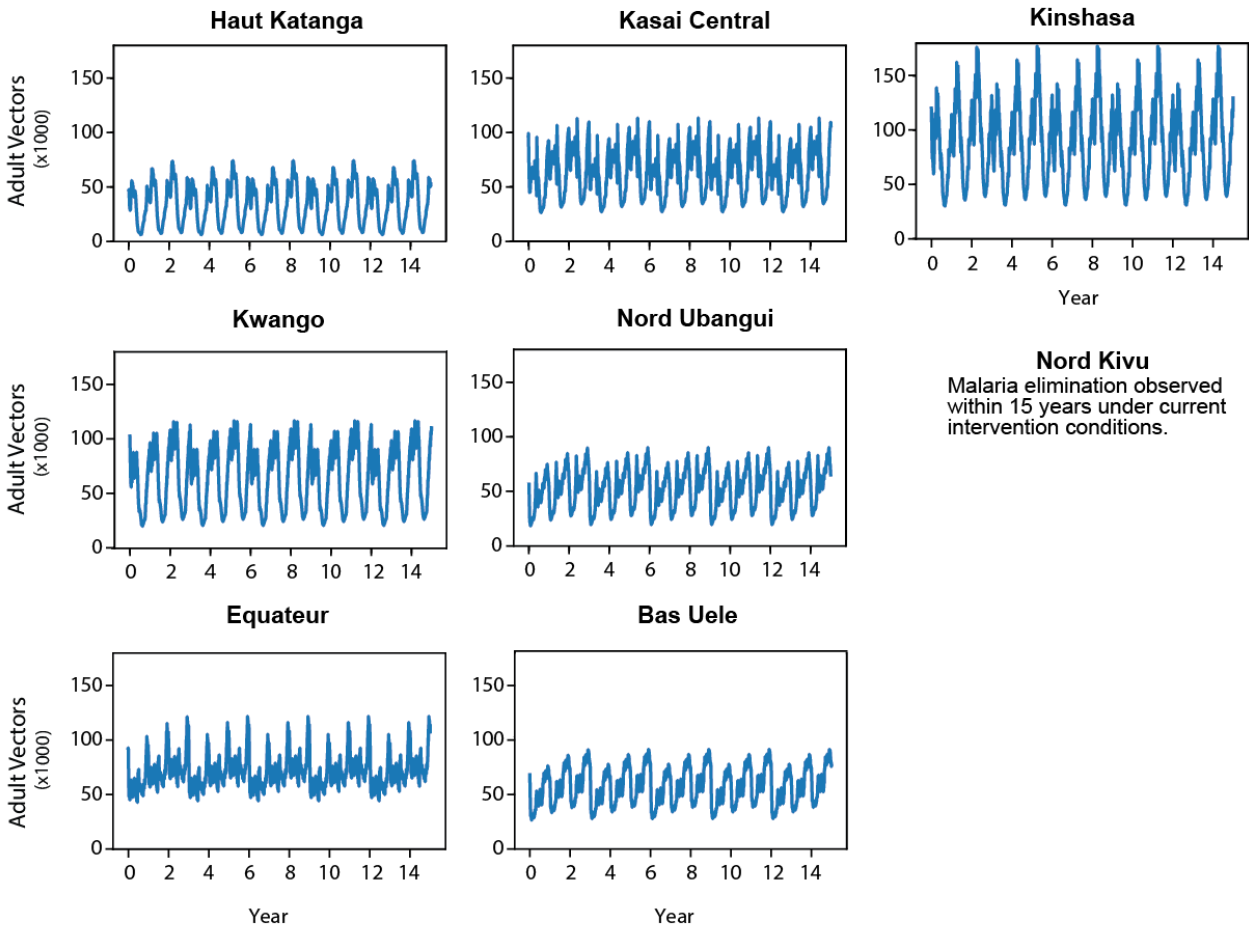
Haut Katanga
Kasai Central
Kinshasa
Kwango
Nord Ubangui
Nord Kivu
Malaria elimination observed within 15 years under current intervention conditions.
Equateur
Bas Uele
Adult Vectors (x1000)
Year
0
50
100
150
0 2 4 6 8 10 12 14

6

7 *Simulation outputs by site:*

**Bas Uele**

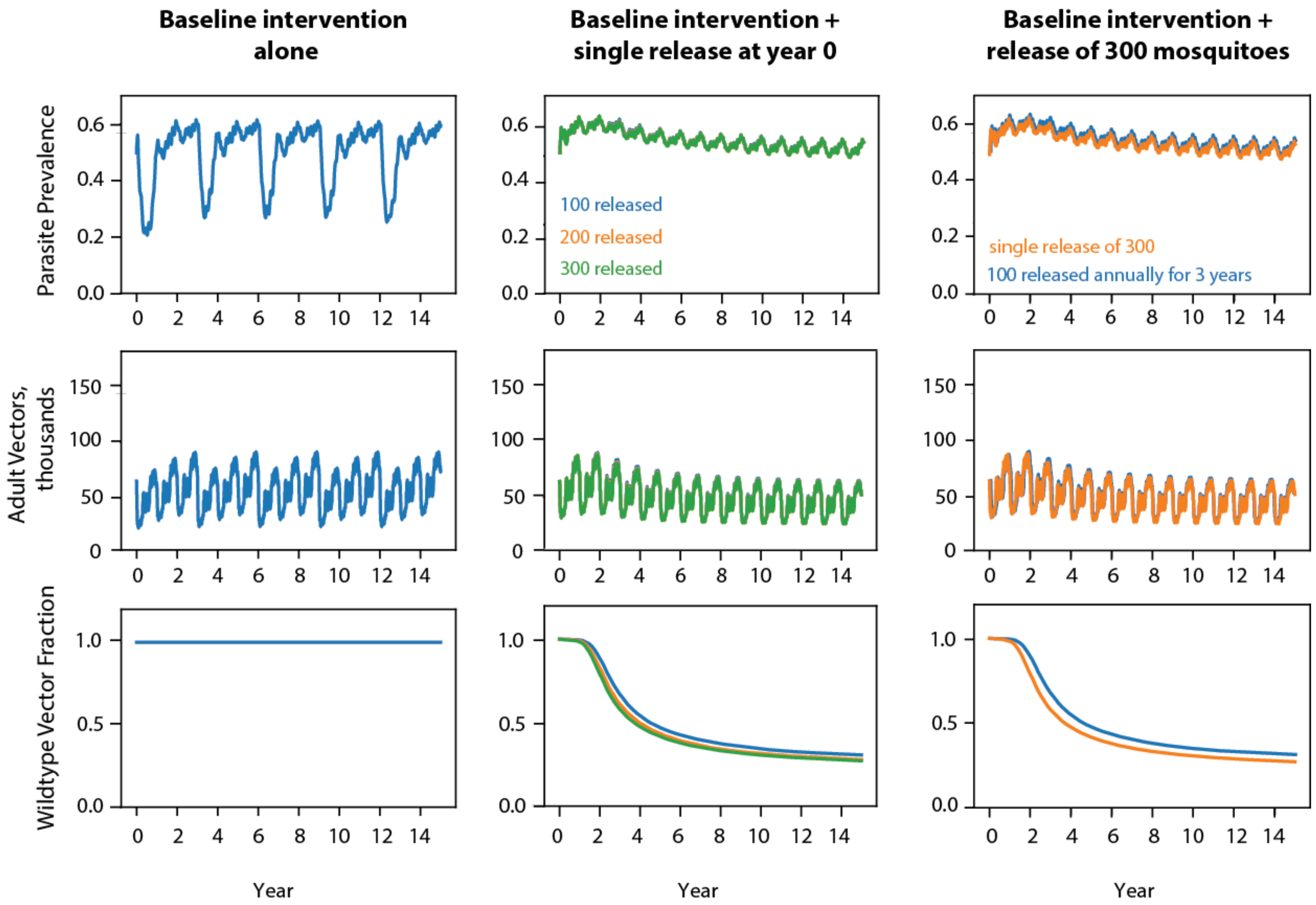

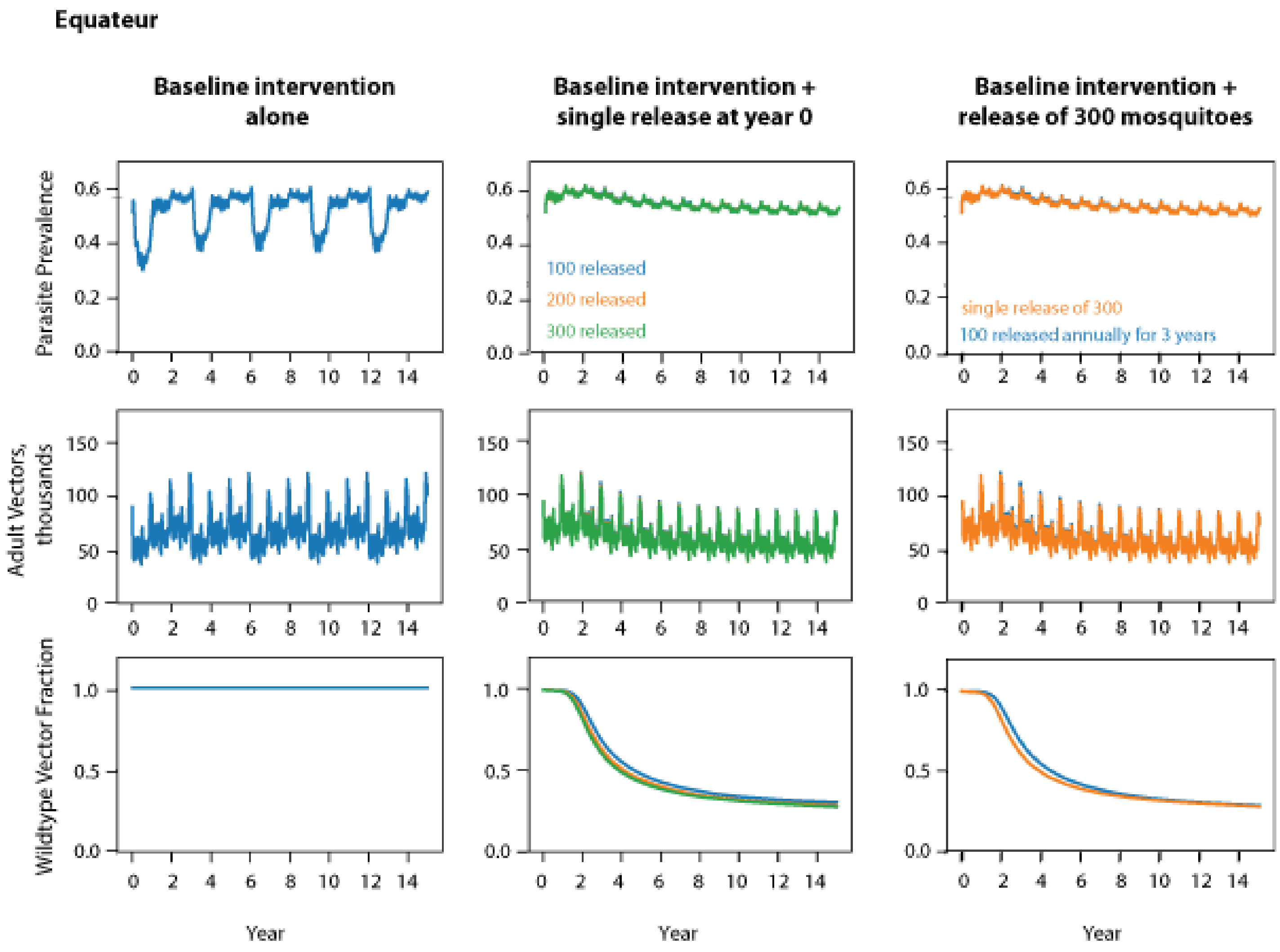
Equateur
Baseline intervention alone
Baseline intervention + single release at year 0
Baseline intervention + release of 300 mosquitoes
Parasite Prevalence
Adult Vectors, thousands
Wildtype Vector Fraction
100 released
200 released
300 released
single release of 300
100 released annually for 3 years
Year
Year
Year

8

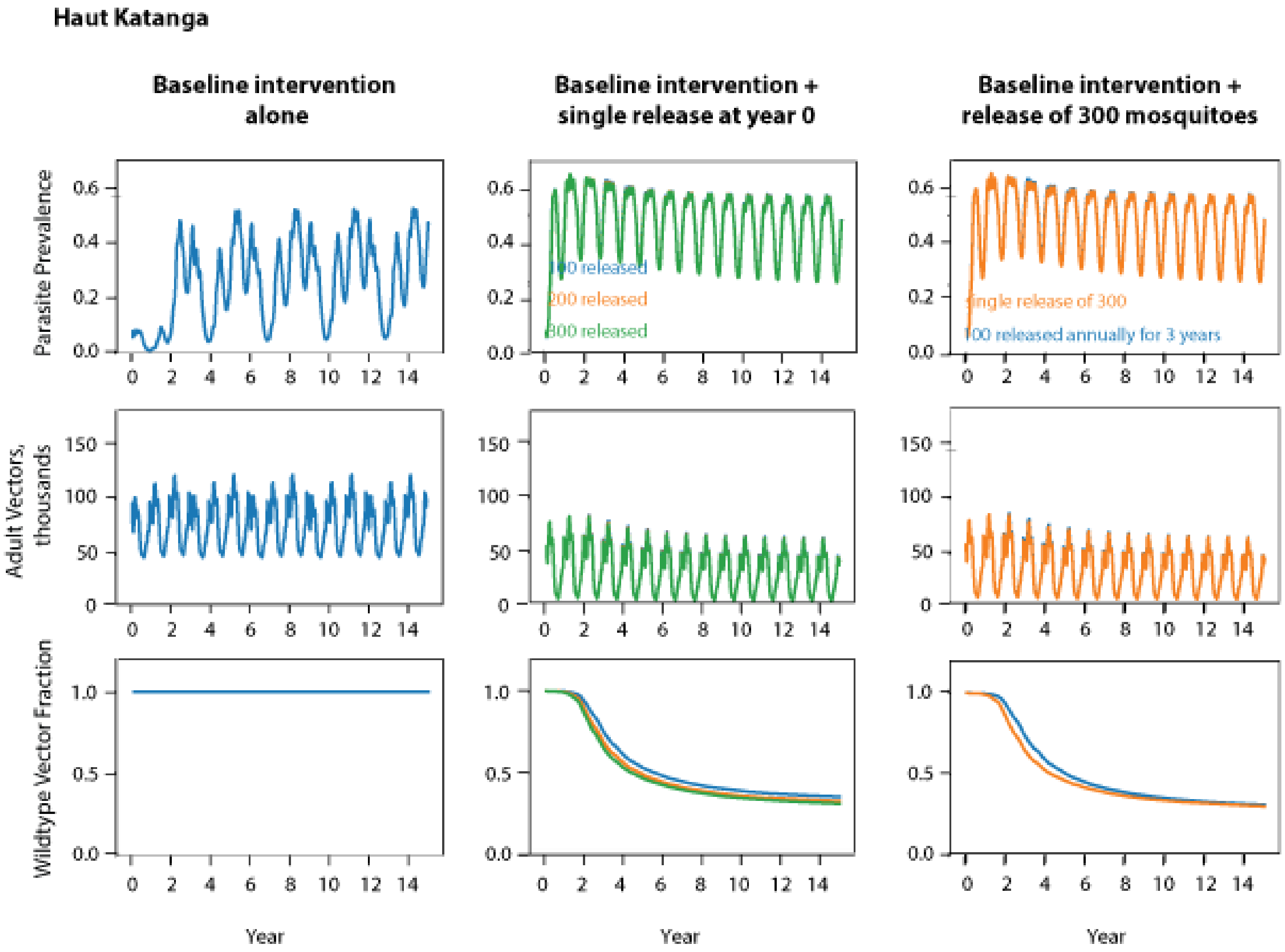
Haut Katanga
Baseline intervention alone
Baseline intervention + single release at year 0
Baseline intervention + release of 300 mosquitoes
Parasite Prevalence
Adult Vectors, thousands
Wildtype Vector Fraction
100 released
200 released
300 released
single release of 300
100 released annually for 3 years
Year
Year
Year

9

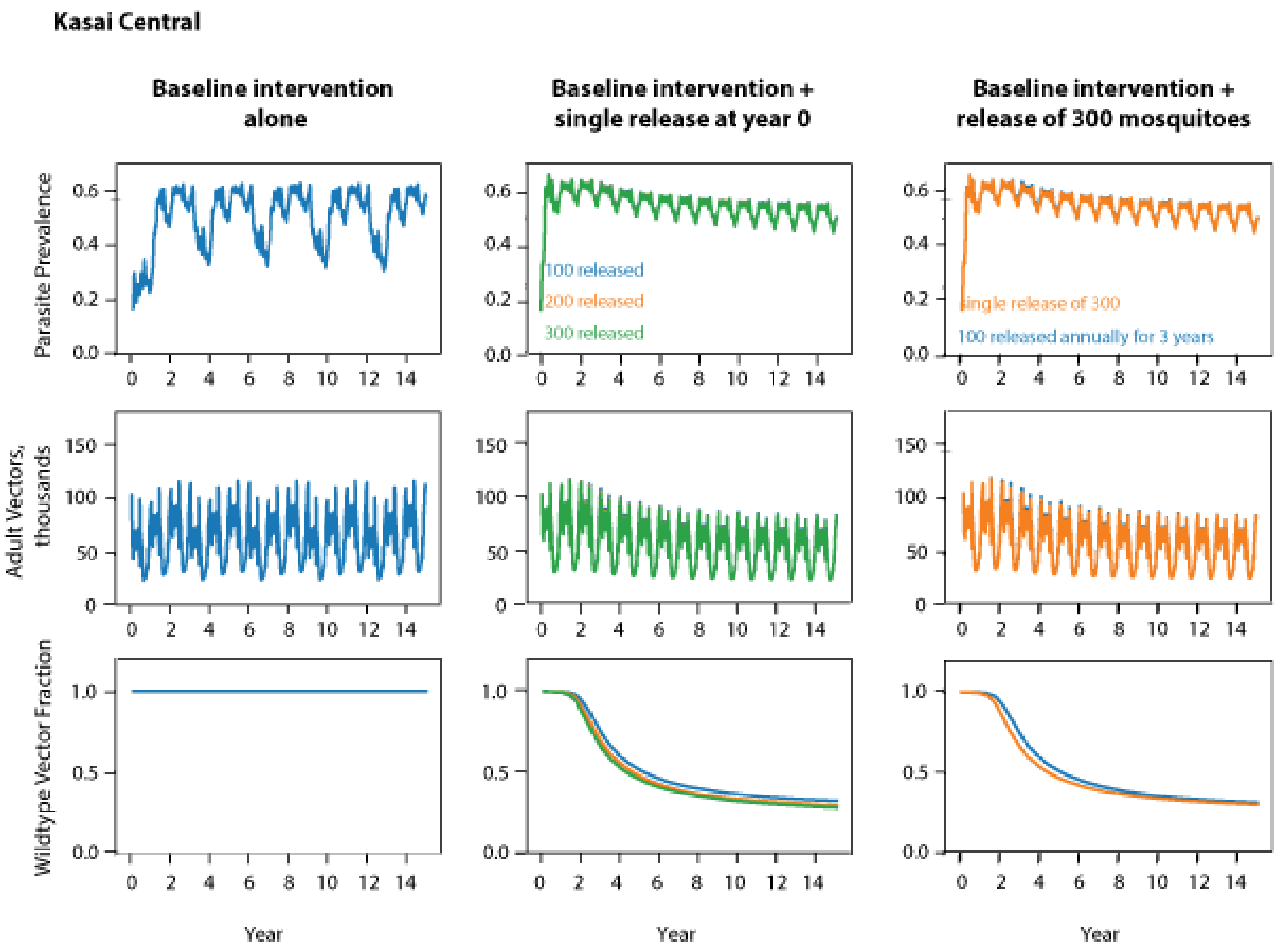
Kasai Central
Baseline intervention alone
Baseline intervention + single release at year 0
Baseline intervention + release of 300 mosquitoes
Parasite Prevalence
Adult Vectors, thousands
Wildtype Vector Fraction
100 released
200 released
300 released
single release of 300
100 released annually for 3 years
Year
Year
Year

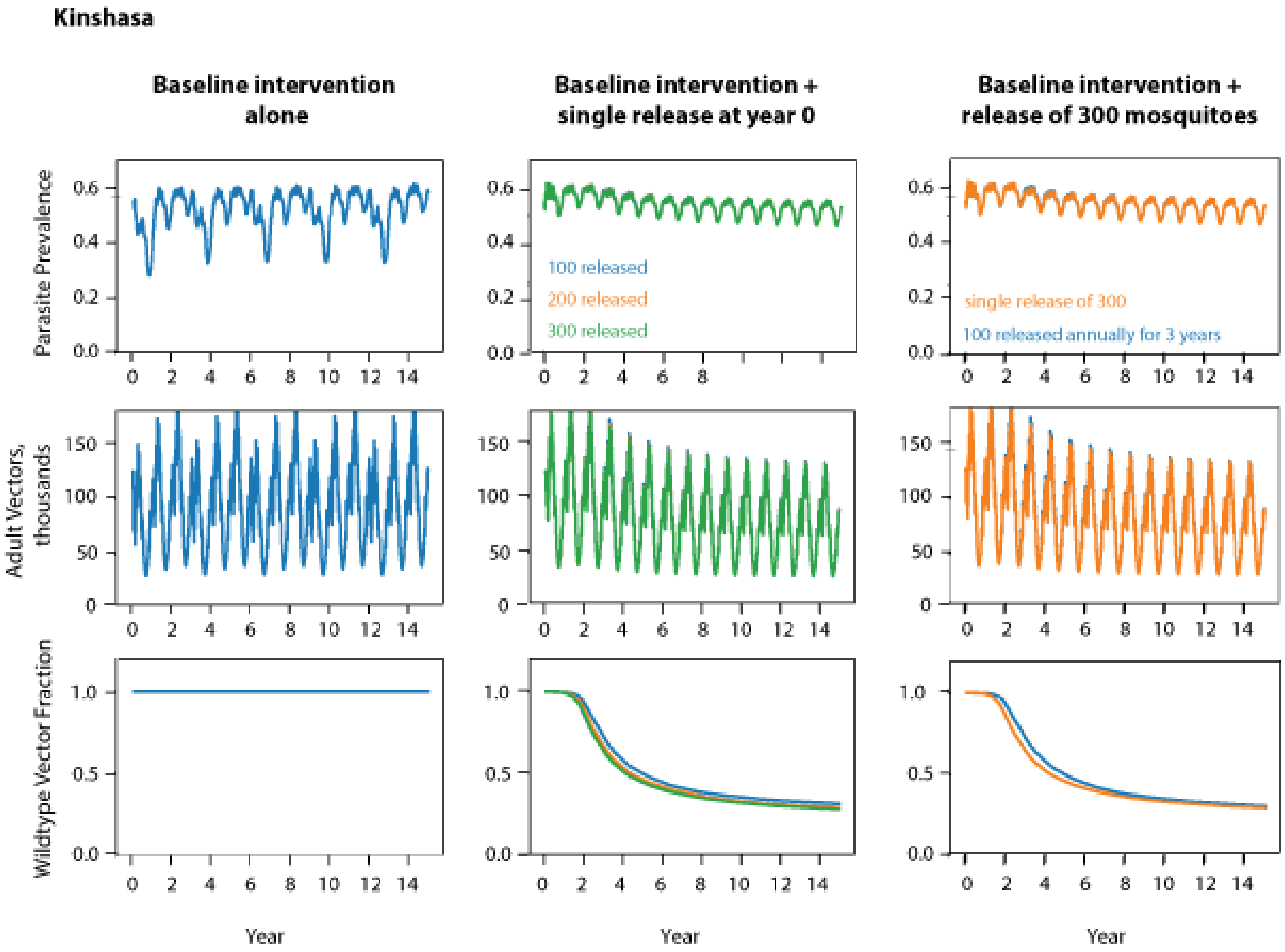
Kinshasa
Baseline intervention alone
Baseline intervention + single release at year 0
Baseline intervention + release of 300 mosquitoes
Parasite Prevalence
Adult Vectors, thousands
Wildtype Vector Fraction
100 released
200 released
300 released
single release of 300
100 released annually for 3 years
Year
Year
Year

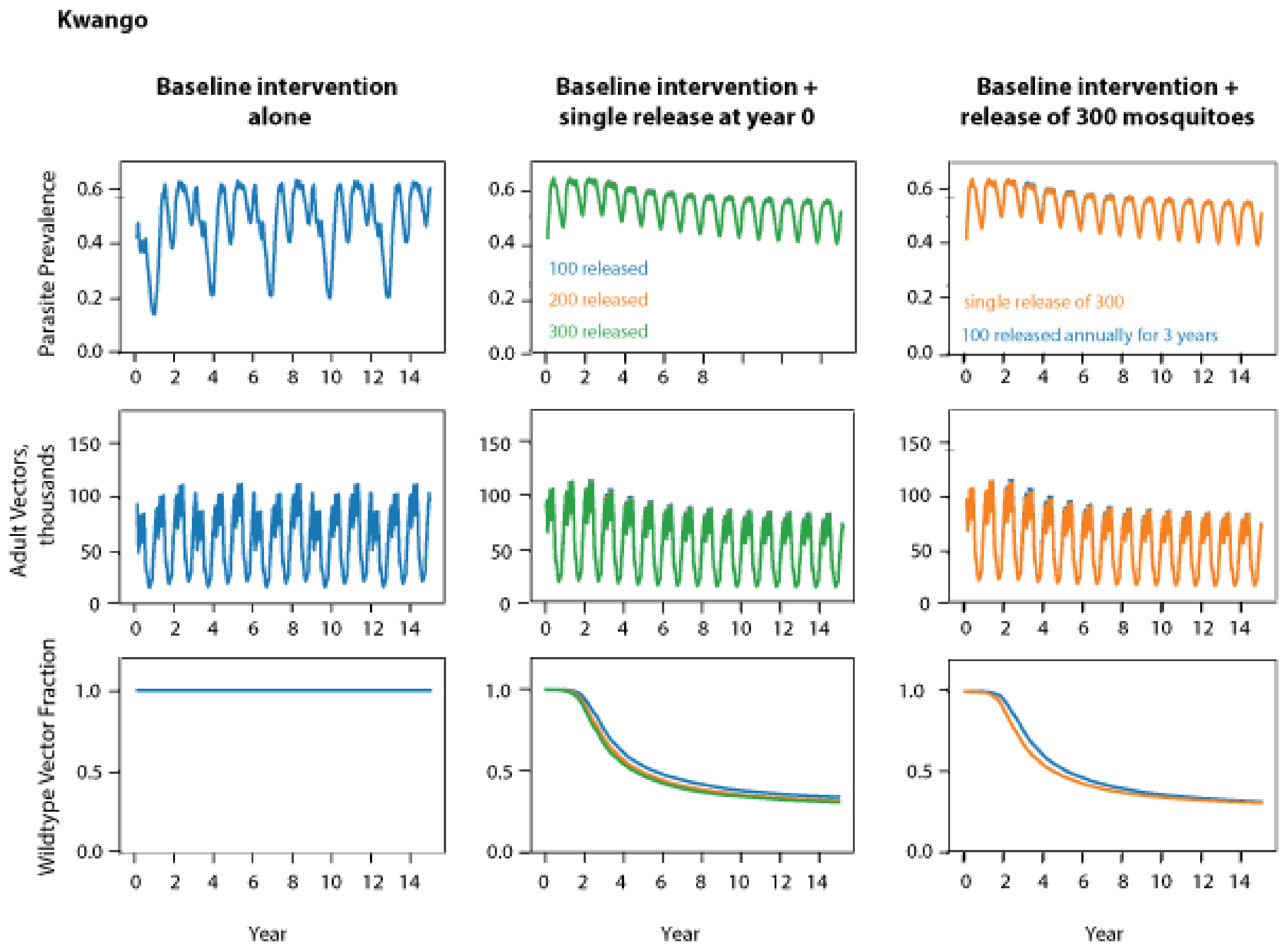
Kwango
Baseline intervention alone
Baseline intervention + single release at year 0
Baseline intervention + release of 300 mosquitoes
Parasite Prevalence
Adult Vectors, thousands
Wildtype Vector Fraction
100 released
200 released
300 released
single release of 300
100 released annually for 3 years
Year
Year
Year

12

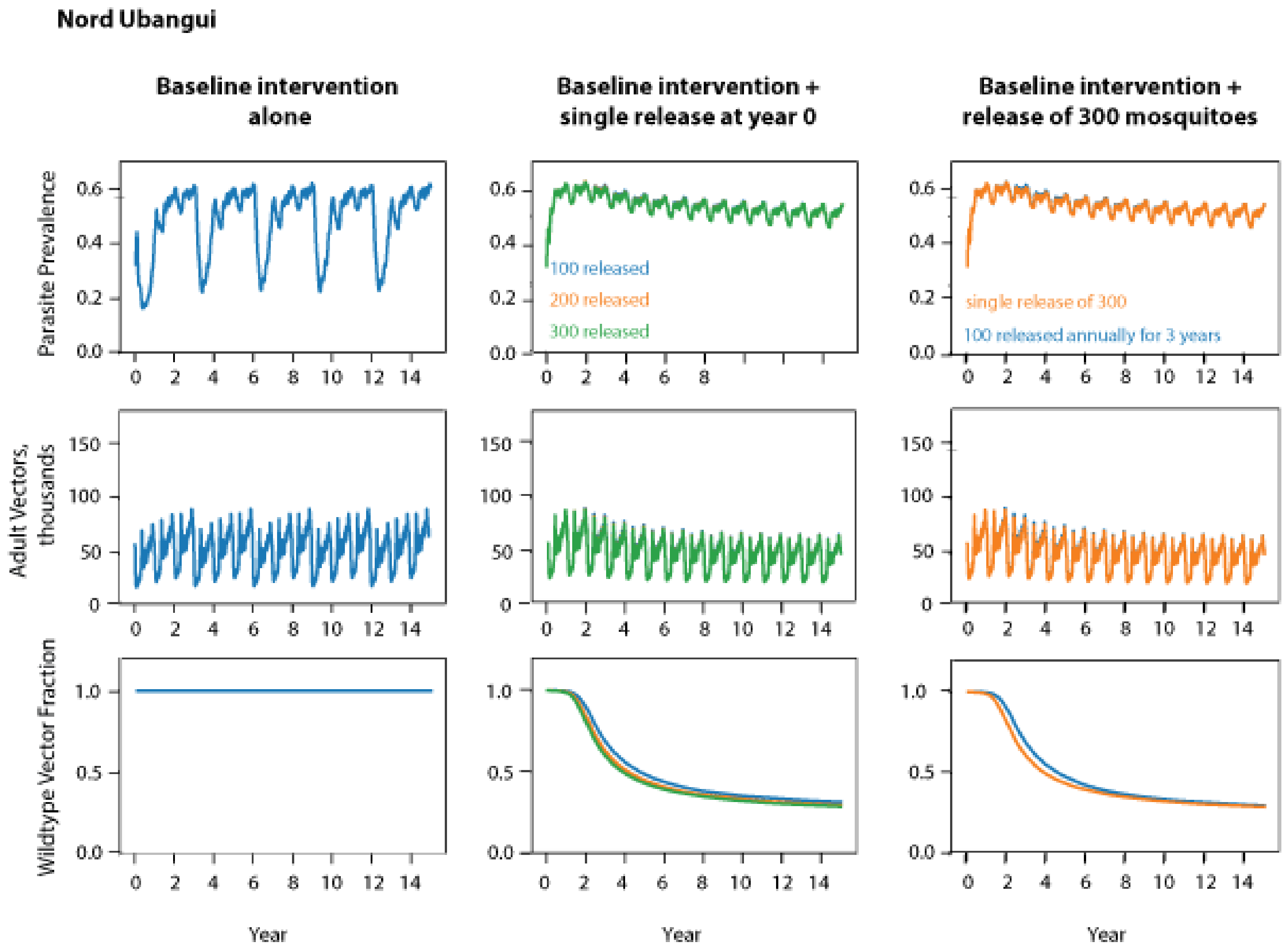
Nord Ubangui
Baseline intervention alone
Baseline intervention + single release at year 0
Baseline intervention + release of 300 mosquitoes
Parasite Prevalence
Adult Vectors, thousands
Wildtype Vector Fraction
100 released
200 released
300 released
single release of 300
100 released annually for 3 years
Year
Year
Year

13

**Supplementary 2, Non-spatial simulation framework: driving-Y parameters of gene-drive mosquitoes single release at year 0.**

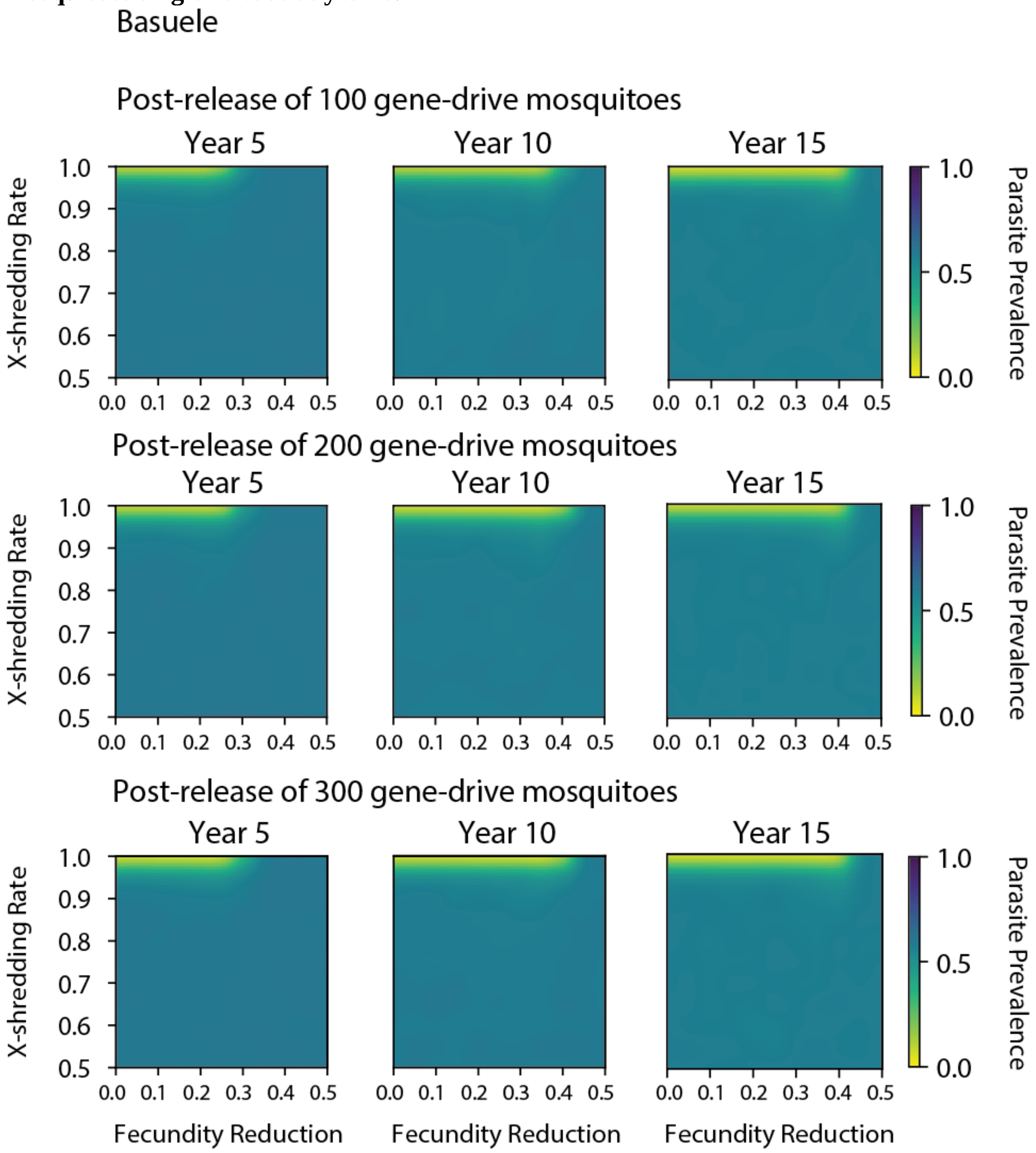

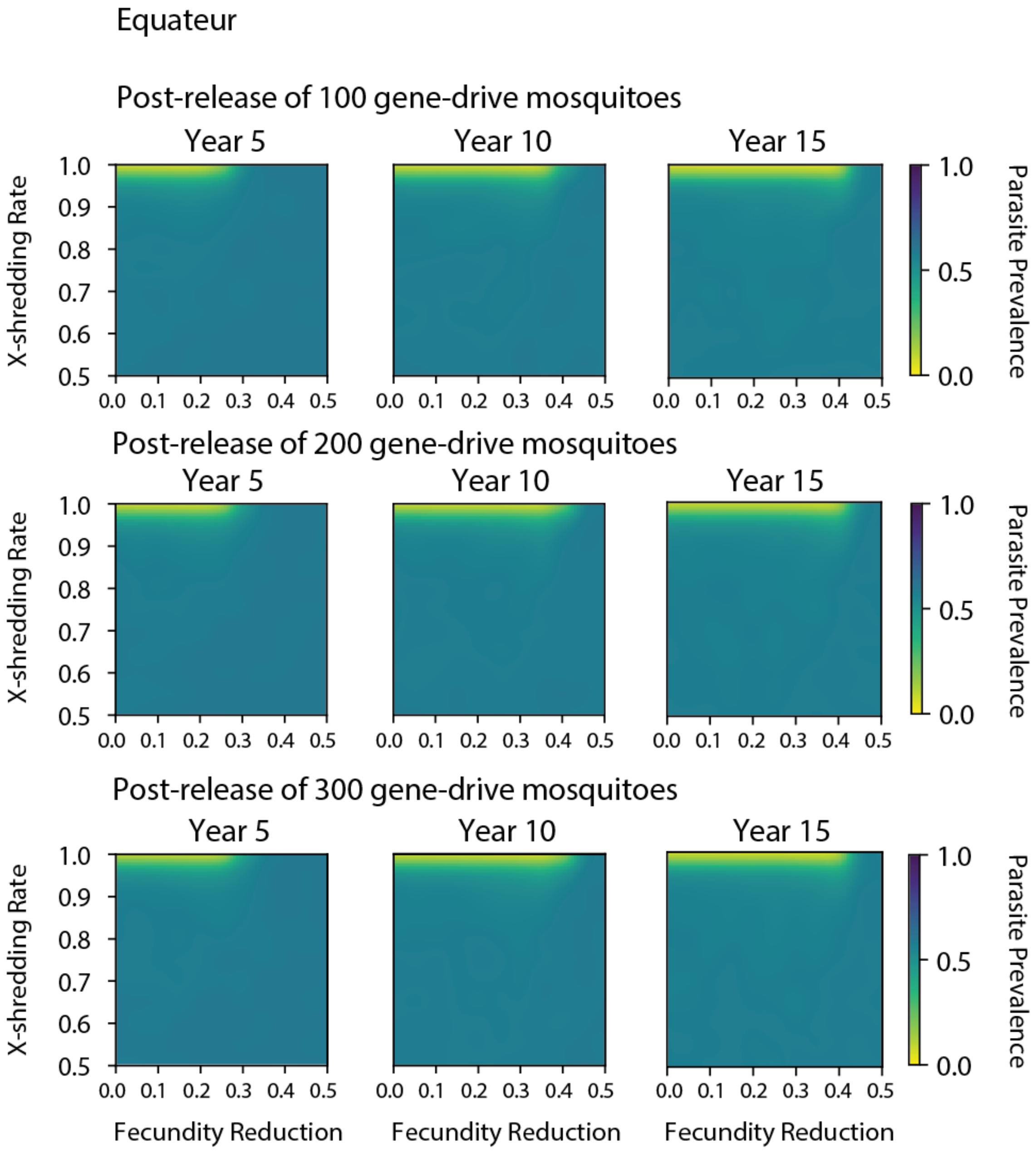
Equateur
Post-release of 100 gene-drive mosquitoes
Post-release of 200 gene-drive mosquitoes
Post-release of 300 gene-drive mosquitoes
Year 5
Year 10
Year 15
X-shredding Rate
Fecundity Reduction
Parasite Prevalence
1.0
0.9
0.8
0.7
0.6
0.5
0.0
0.1
0.2
0.3
0.4

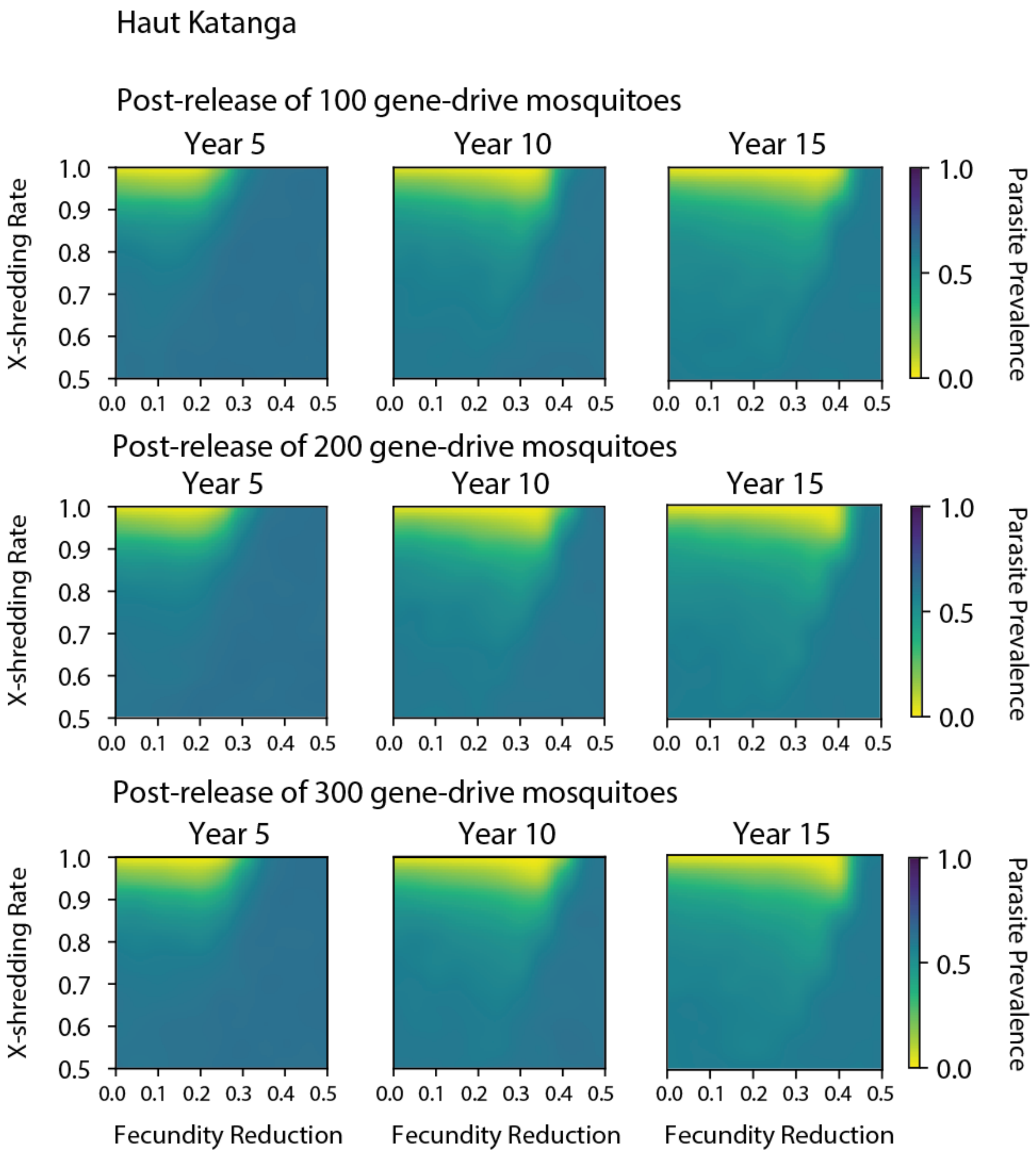
Haut Katanga
Post-release of 100 gene-drive mosquitoes
Year 5
Year 10
Year 15
X-shredding Rate
Parasite Prevalence
Post-release of 200 gene-drive mosquitoes
Post-release of 300 gene-drive mosquitoes
Fecundity Reduction
1.0
0.9
0.8
0.7
0.6
0.5
0.0
0.1
0.2
0.3
0.4

18

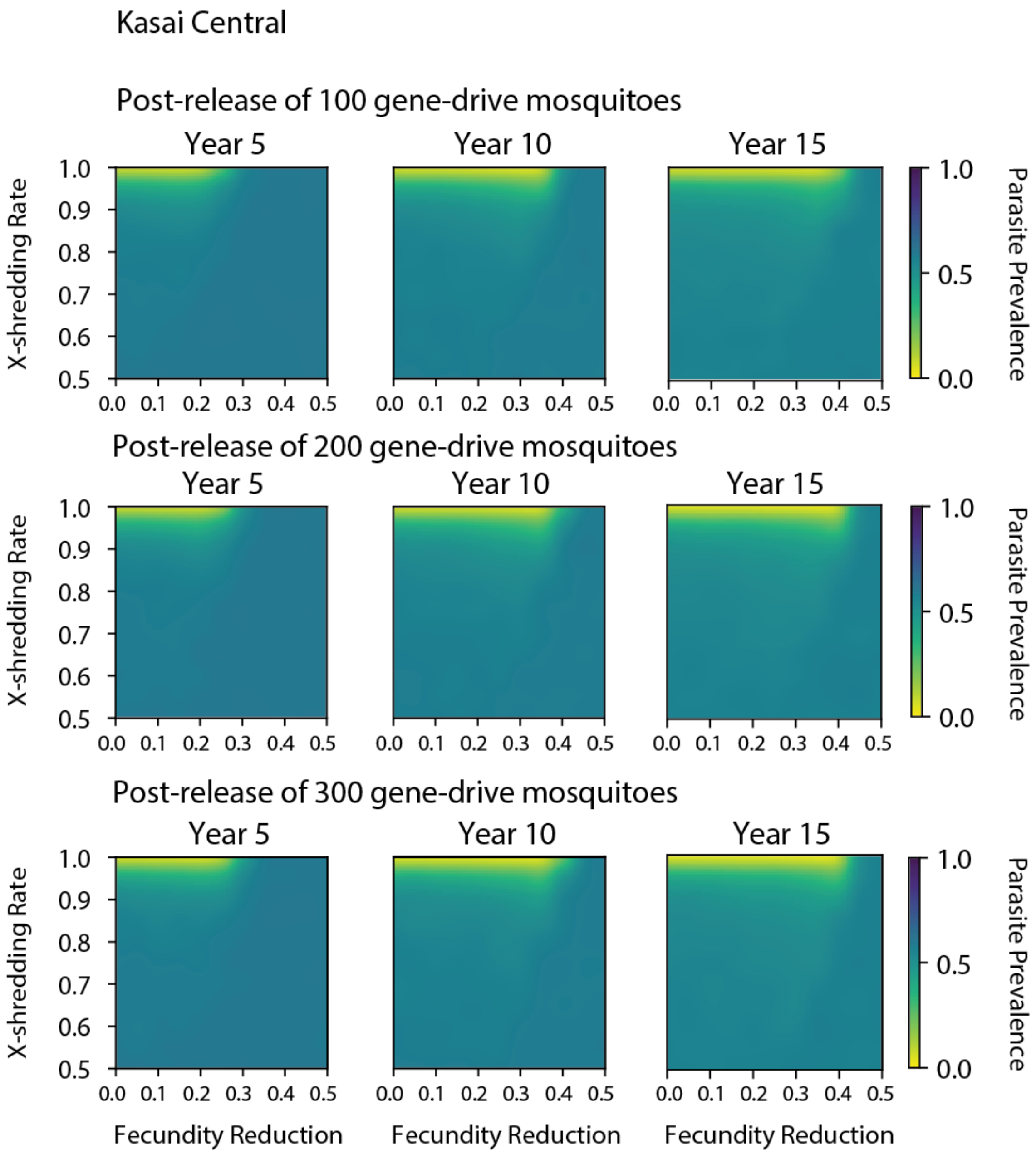
Kasai Central
Post-release of 100 gene-drive mosquitoes
Year 5
Year 10
Year 15
X-shredding Rate
1.0
0.9
0.8
0.7
0.6
0.5
0.0 0.1 0.2 0.3 0.4 0.5
Parasite Prevalence
1.0
0.5
0.0
Post-release of 200 gene-drive mosquitoes
Post-release of 300 gene-drive mosquitoes
Fecundity Reduction

19

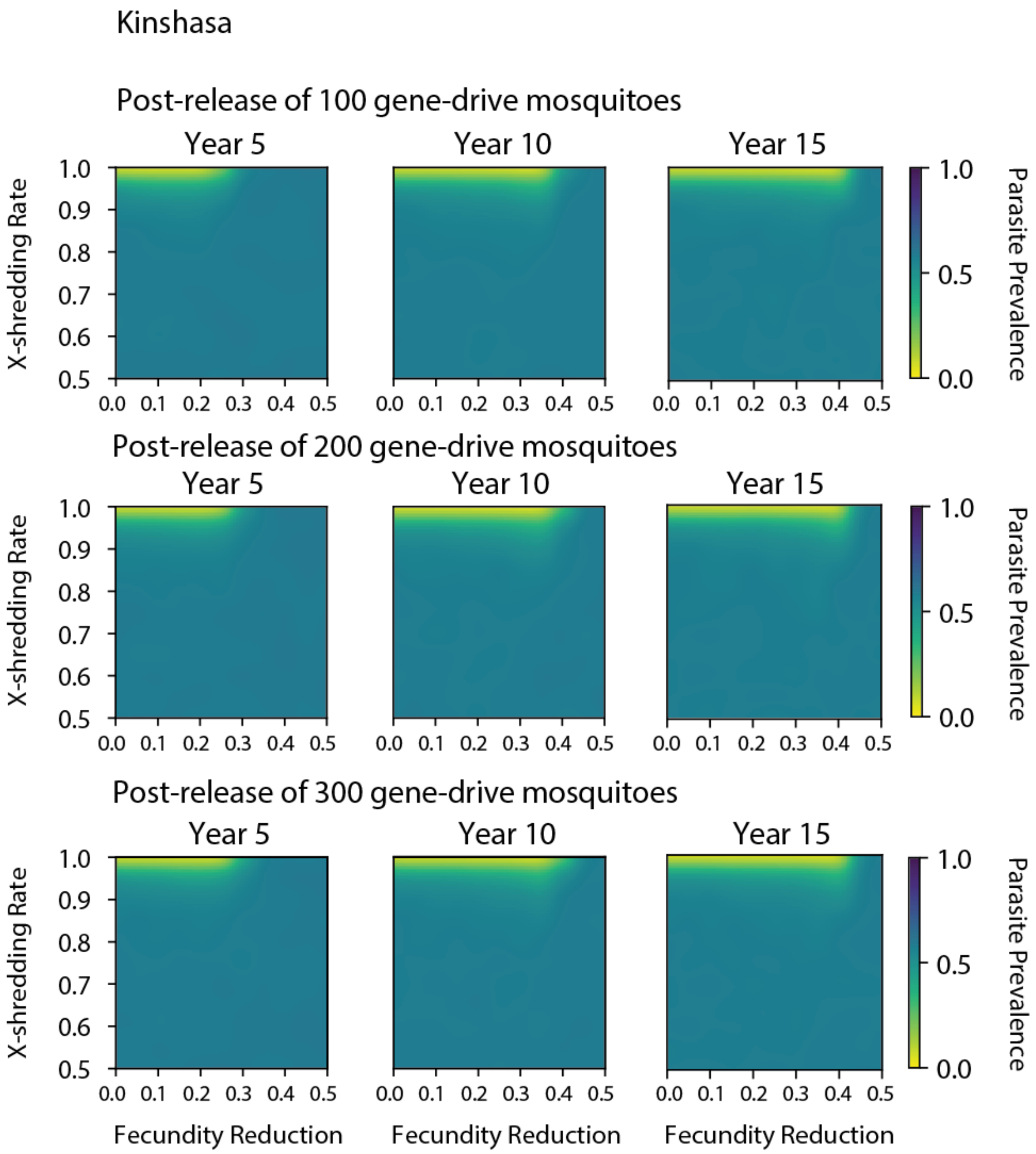
Kinshasa
Post-release of 100 gene-drive mosquitoes
Year 5
Year 10
Year 15
X-shredding Rate
1.0
0.9
0.8
0.7
0.6
0.5
0.0 0.1 0.2 0.3 0.4 0.5
Parasite Prevalence
1.0
0.5
0.0
Post-release of 200 gene-drive mosquitoes
Year 5
Year 10
Year 15
X-shredding Rate
Parasite Prevalence
Post-release of 300 gene-drive mosquitoes
Year 5
Year 10
Year 15
X-shredding Rate
Parasite Prevalence
Fecundity Reduction
Fecundity Reduction
Fecundity Reduction

21 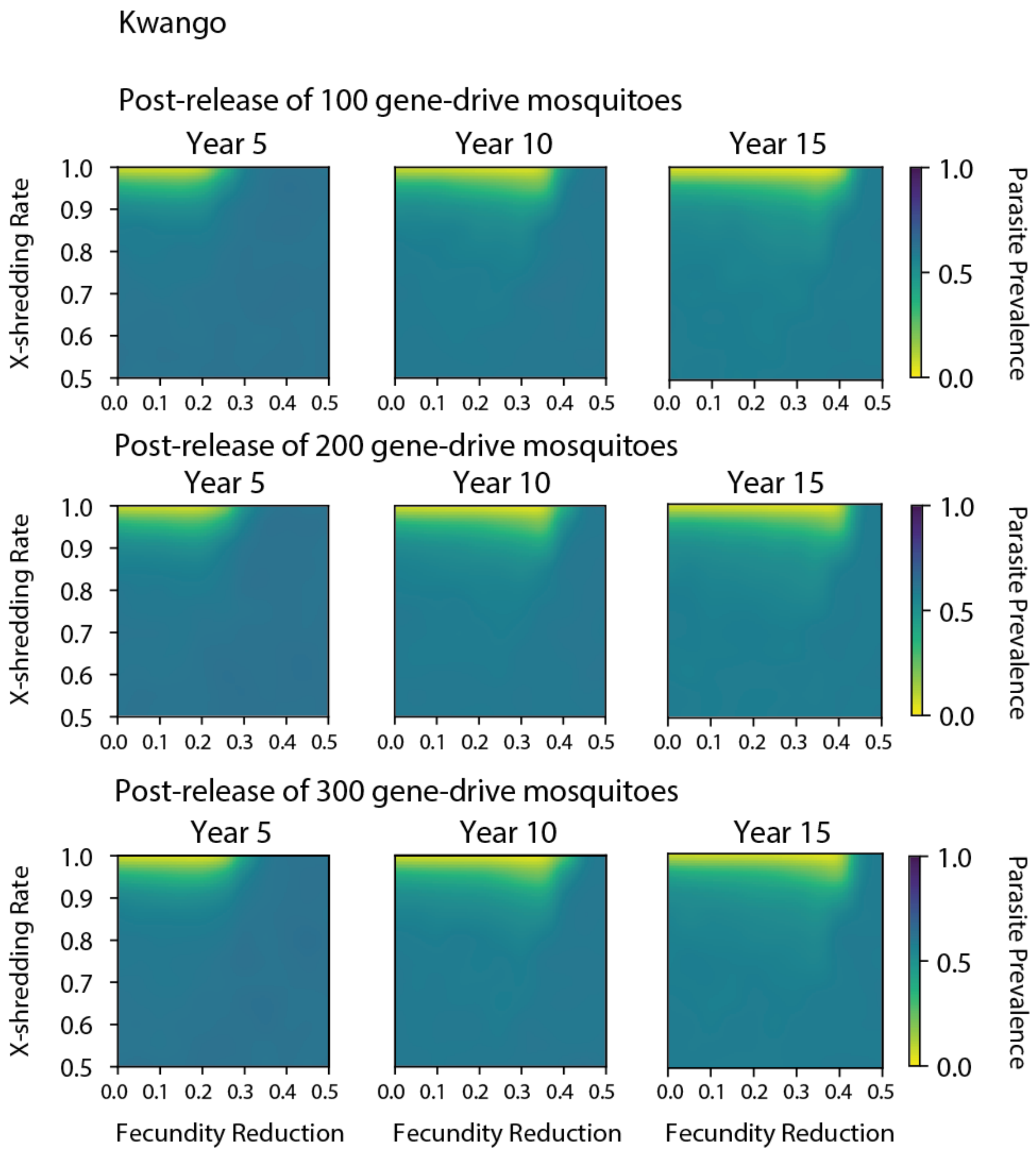
Kwango
Post-release of 100 gene-drive mosquitoes
Year 5
Year 10
Year 15
X-shredding Rate
Parasite Prevalence
Post-release of 200 gene-drive mosquitoes
Post-release of 300 gene-drive mosquitoes
Fecundity Reduction
1.0
0.9
0.8
0.7
0.6
0.5
0.0
0.1
0.2
0.3
0.4

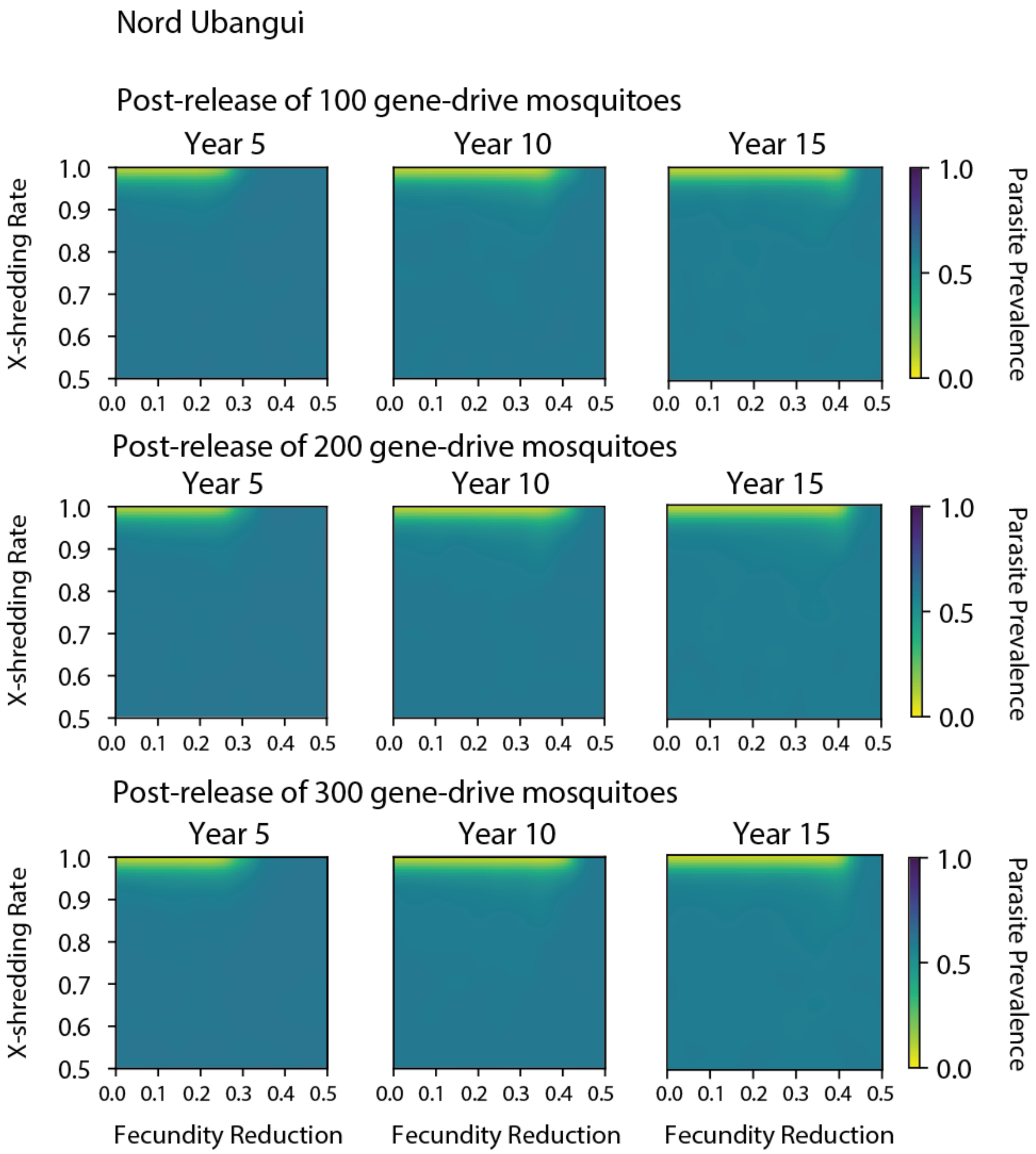
Nord Ubangui
Post-release of 100 gene-drive mosquitoes
Post-release of 200 gene-drive mosquitoes
Post-release of 300 gene-drive mosquitoes
Year 5
Year 10
Year 15
X-shredding Rate
Fecundity Reduction
Parasite Prevalence
1.0
0.9
0.8
0.7
0.6
0.5
0.0 0.1 0.2 0.3 0.4 0.5
1.0
0.5
0.0

22

**<u>Supplementary 3</u>, Non-spatial simulation framework: ratio between current and initial numbers of adult vectors, 15-year post- single release of 300 drive mosquitoes at year 0.**

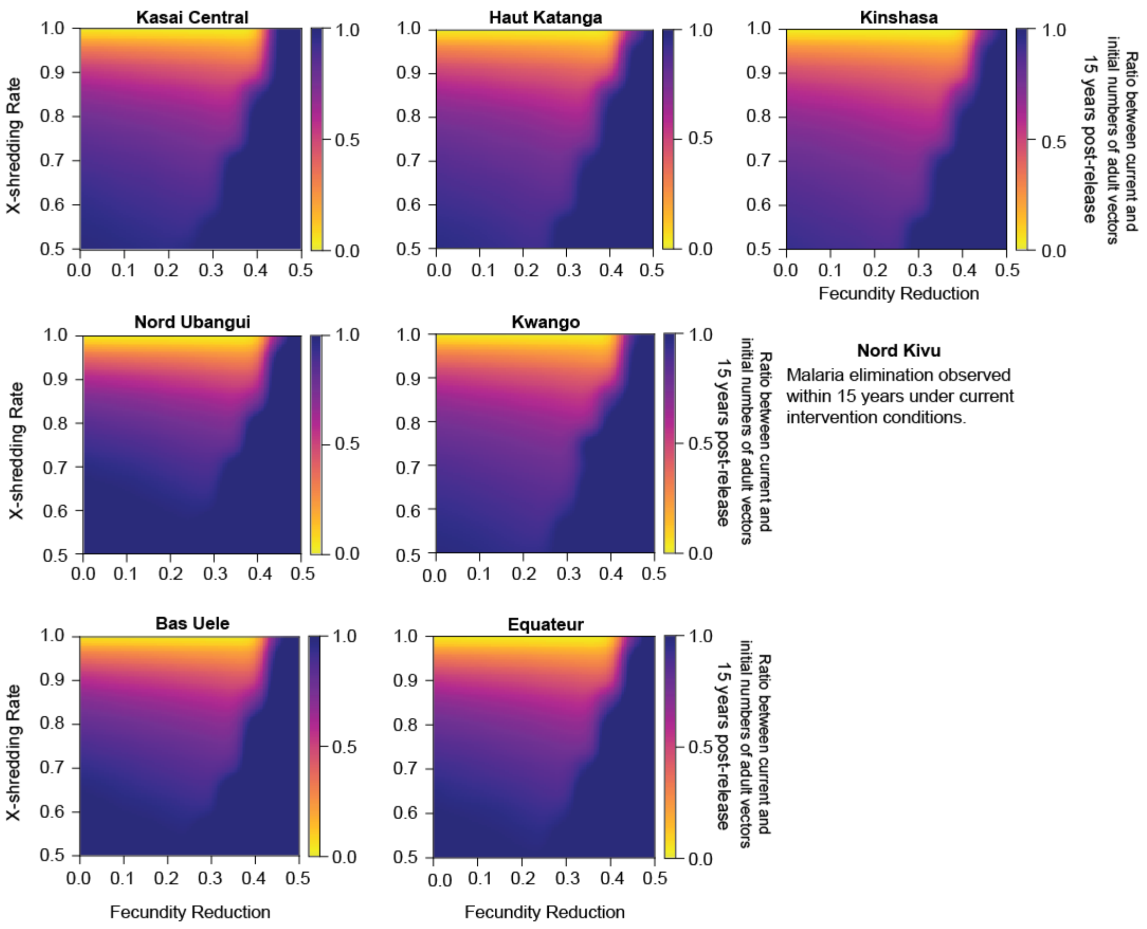

27 **<u>Supplementary 4</u>: Spatial simulation framework: simulation outputs.**

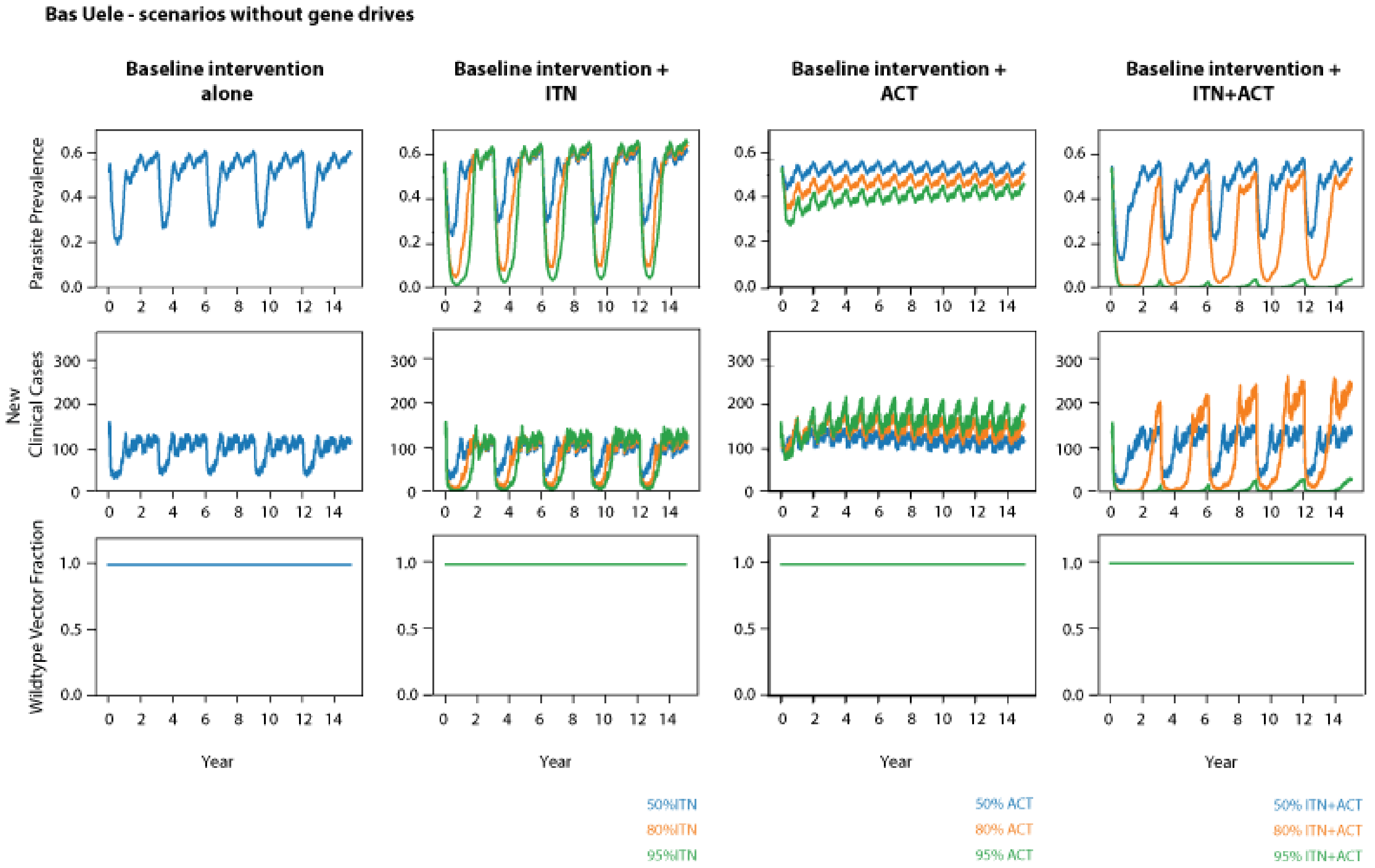


28

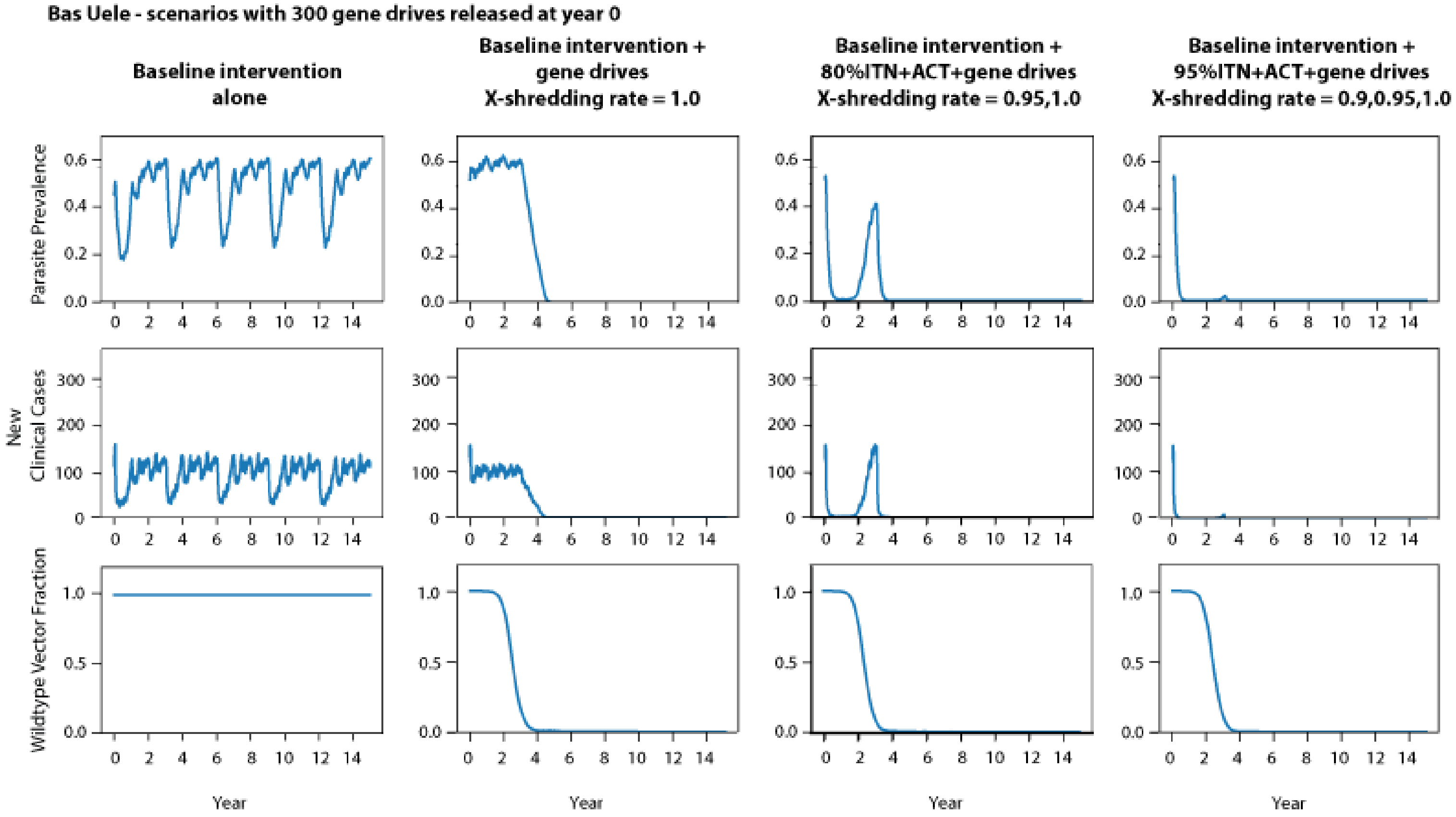
Bas Uele - scenarios with 300 gene drives released at year 0
Baseline intervention alone
Baseline intervention + gene drives X-shredding rate = 1.0
Baseline intervention + 80%ITN+ACT+gene drives X-shredding rate = 0.95,1.0
Baseline intervention + 95%ITN+ACT+gene drives X-shredding rate = 0.9,0.95,1.0
Parasite Prevalence
New Clinical Cases
Wildtype Vector Fraction
Year

29

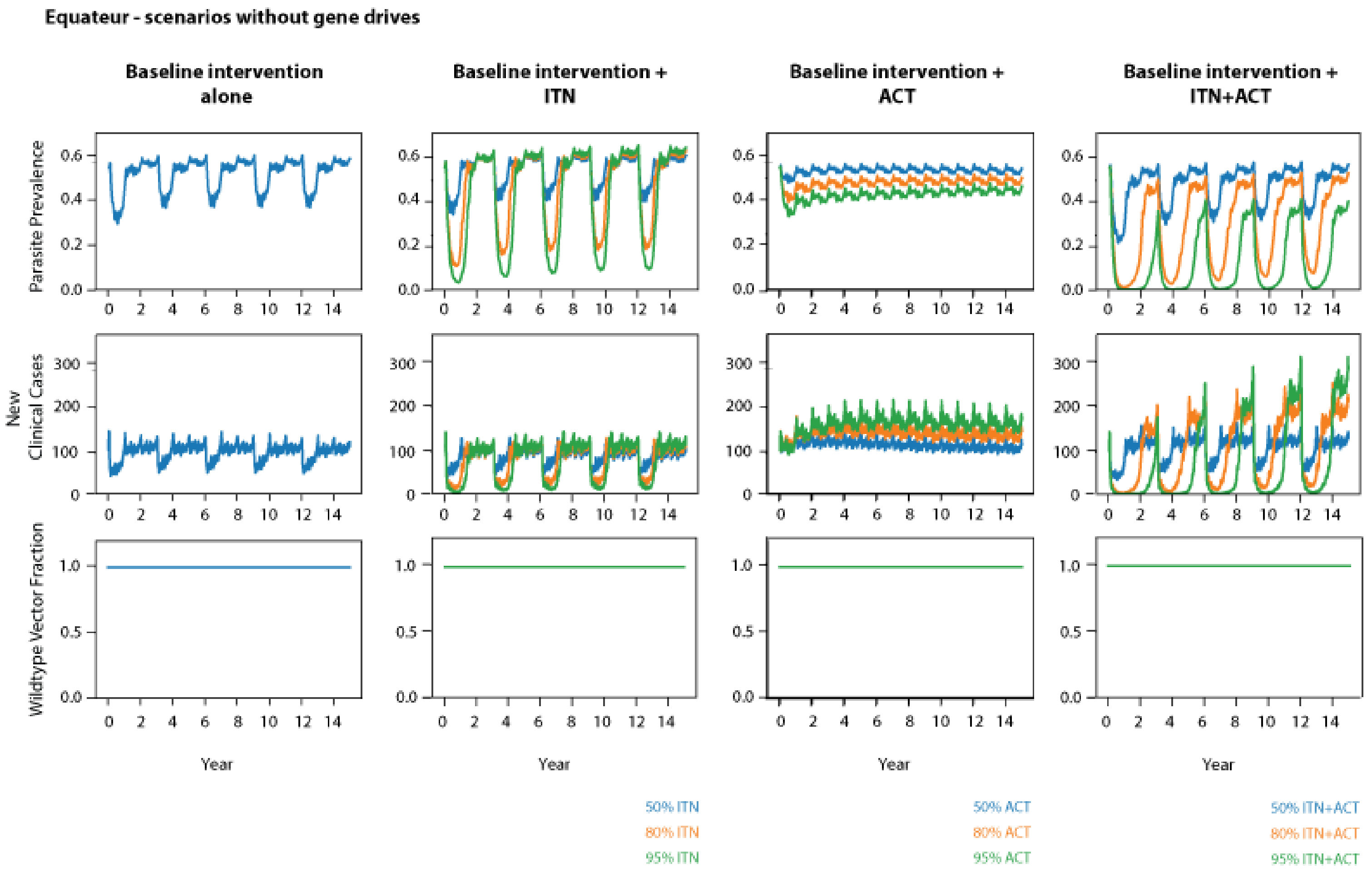
Equateur - scenarios without gene drives
Baseline intervention alone
Baseline intervention + ITN
Baseline intervention + ACT
Baseline intervention + ITN+ACT
Parasite Prevalence
New Clinical Cases
Wildtype Vector Fraction
Year
50% ITN
80% ITN
95% ITN
50% ACT
80% ACT
95% ACT
50% ITN+ACT
80% ITN+ACT
95% ITN+ACT

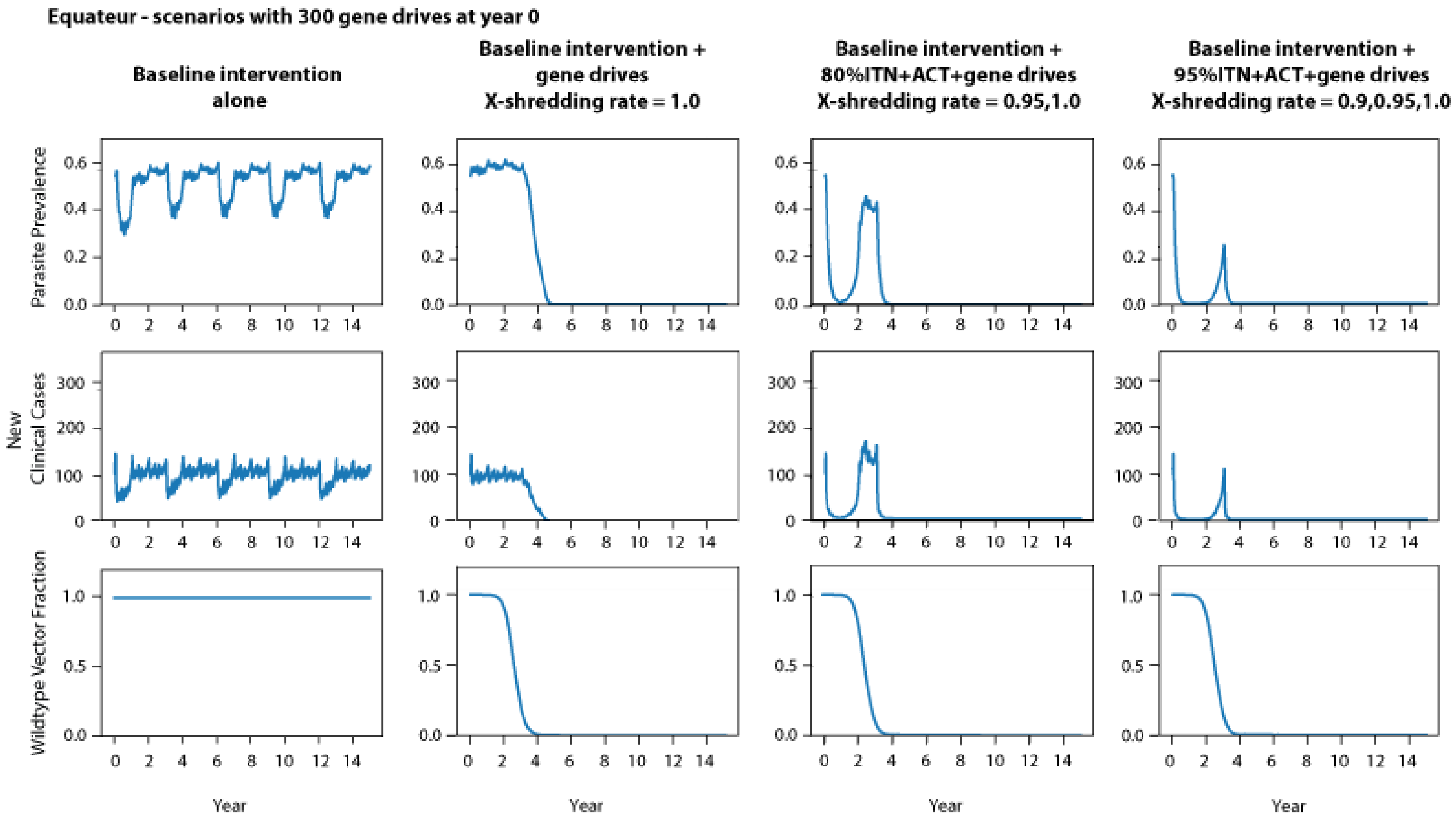
Equateur - scenarios with 300 gene drives at year 0
Baseline intervention alone
Baseline intervention + gene drives X-shredding rate = 1.0
Baseline intervention + 80%ITN+ACT+gene drives X-shredding rate = 0.95,1.0
Baseline intervention + 95%ITN+ACT+gene drives X-shredding rate = 0.9,0.95,1.0
Parasite Prevalence
New Clinical Cases
Wildtype Vector Fraction
Year

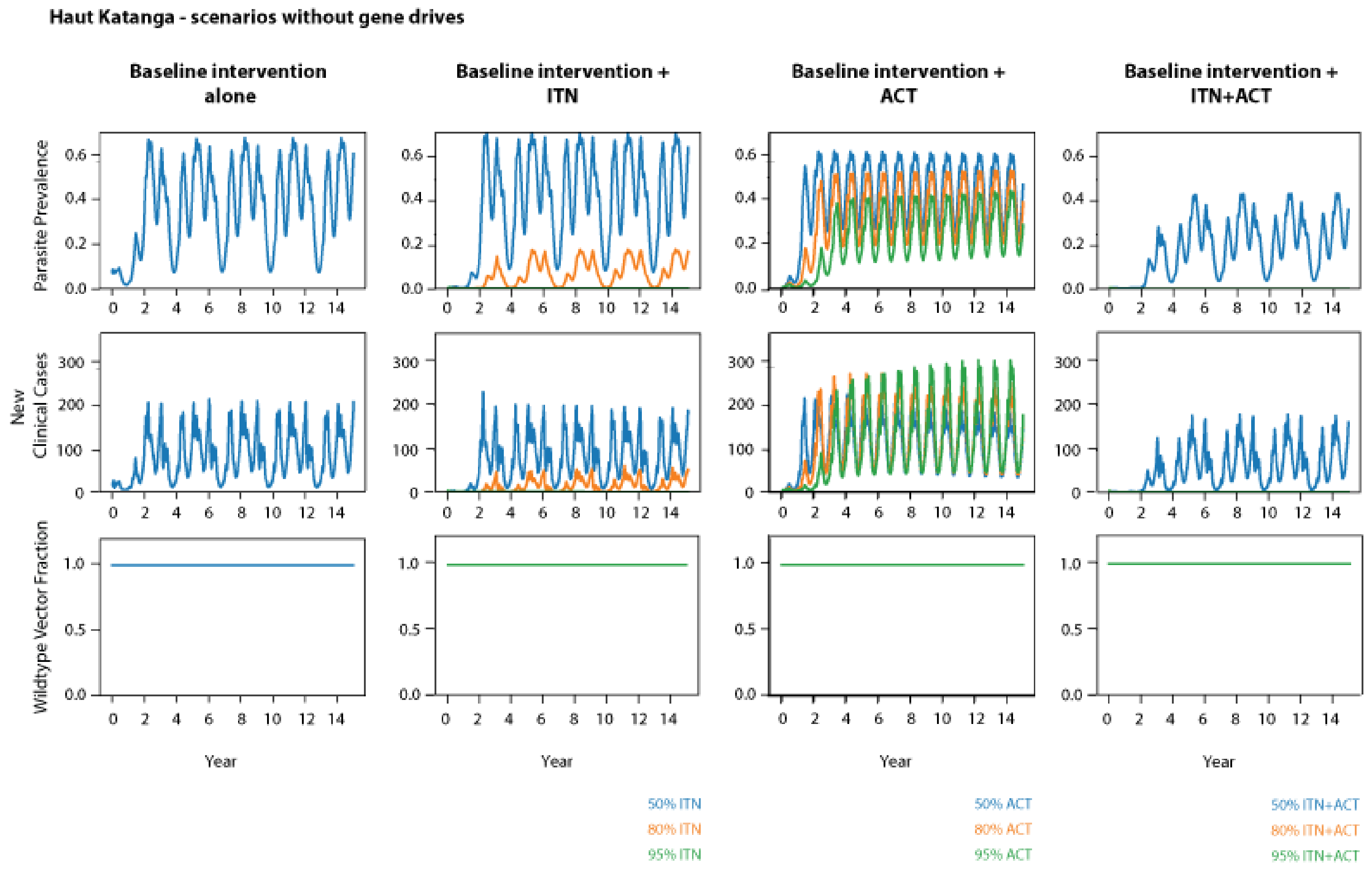
Haut Katanga - scenarios without gene drives
Baseline intervention alone
Baseline intervention + ITN
Baseline intervention + ACT
Baseline intervention + ITN+ACT
Parasite Prevalence
New Clinical Cases
Wildtype Vector Fraction
Year
50% ITN
80% ITN
95% ITN
50% ACT
80% ACT
95% ACT
50% ITN+ACT
80% ITN+ACT
95% ITN+ACT

32

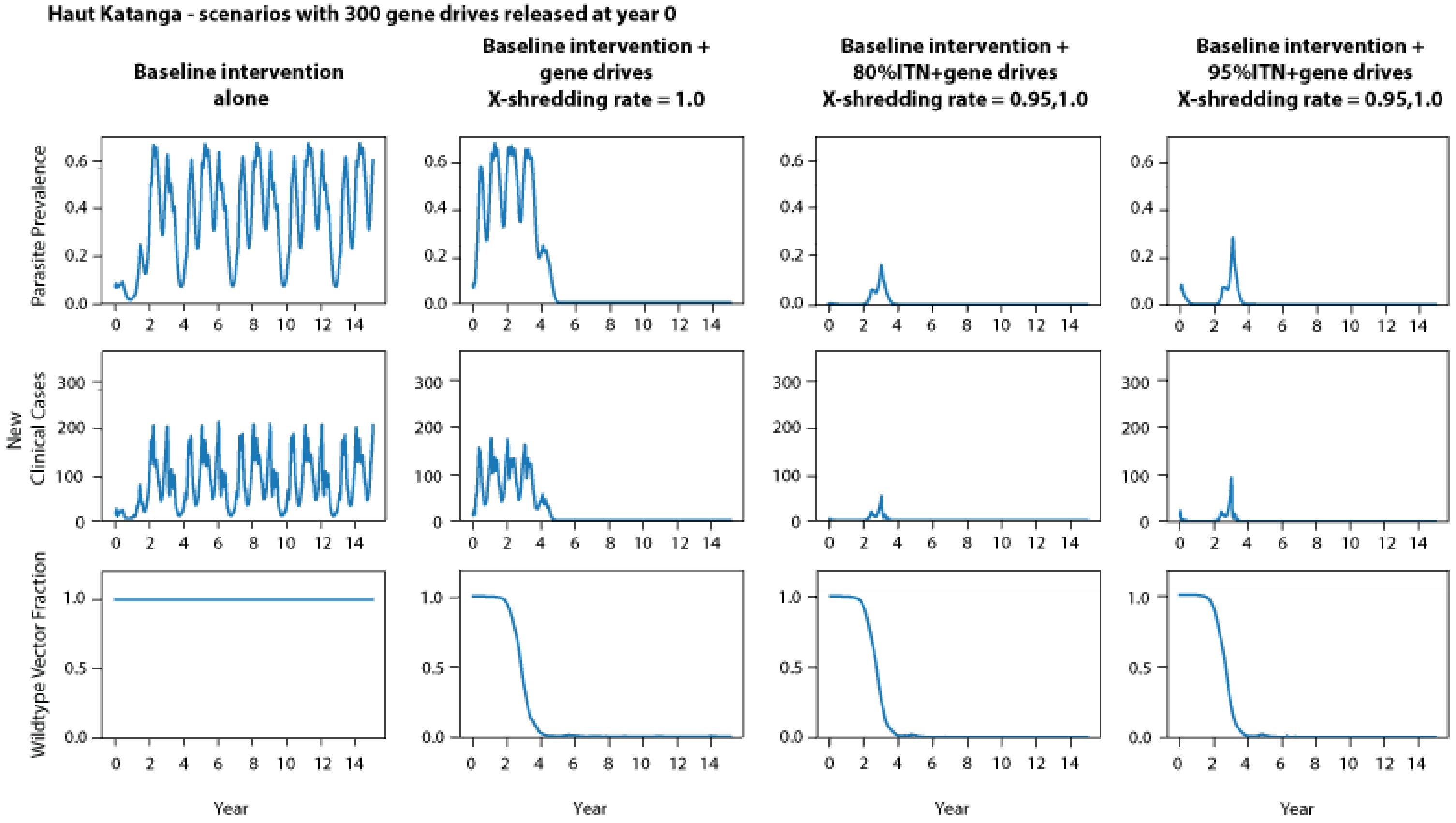
Haut Katanga - scenarios with 300 gene drives released at year 0
Baseline intervention alone
Baseline intervention + gene drives X-shredding rate = 1.0
Baseline intervention + 80%ITN+gene drives X-shredding rate = 0.95,1.0
Baseline intervention + 95%ITN+gene drives X-shredding rate = 0.95,1.0
Parasite Prevalence
New Clinical Cases
Wildtype Vector Fraction
Year

33

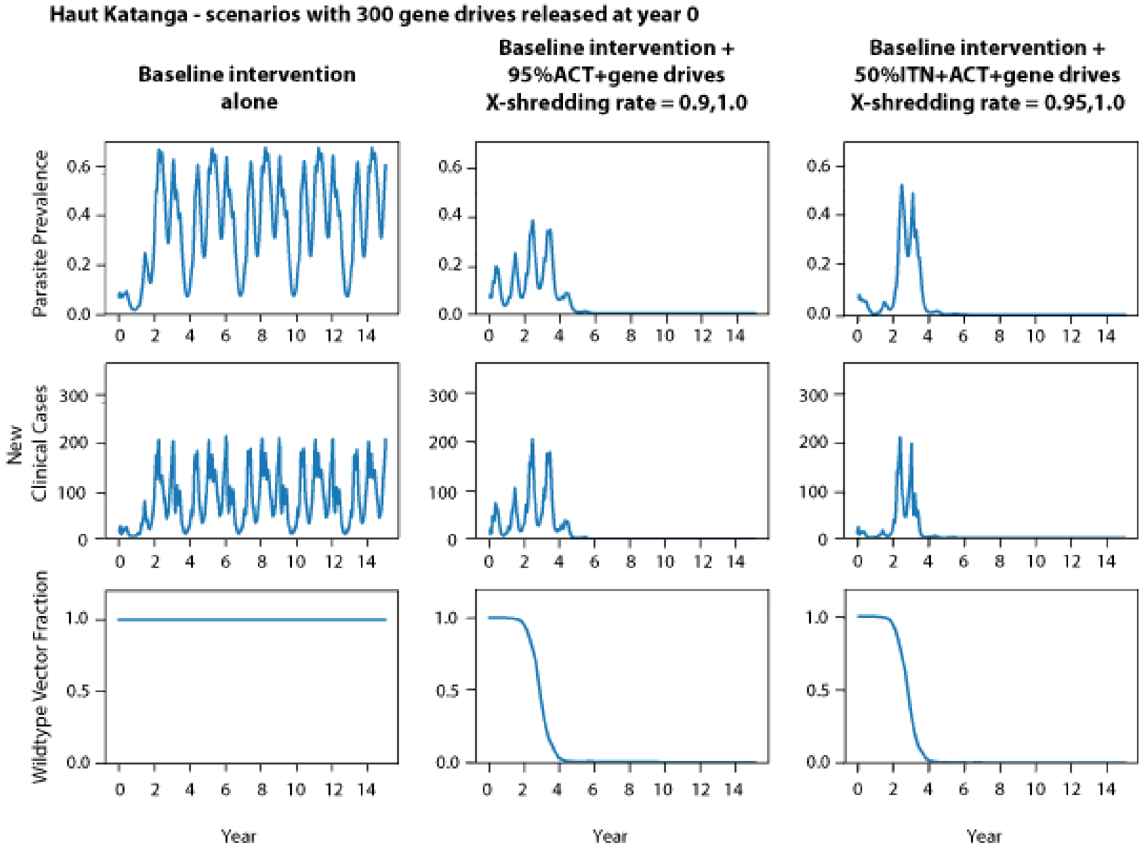
Haut Katanga - scenarios with 300 gene drives released at year 0
Baseline intervention alone
Baseline intervention + 95%ACT+gene drives X-shredding rate = 0.9,1.0
Baseline intervention + 50%ITN+ACT+gene drives X-shredding rate = 0.95,1.0
Parasite Prevalence
New Clinical Cases
Wildtype Vector Fraction
Year

34

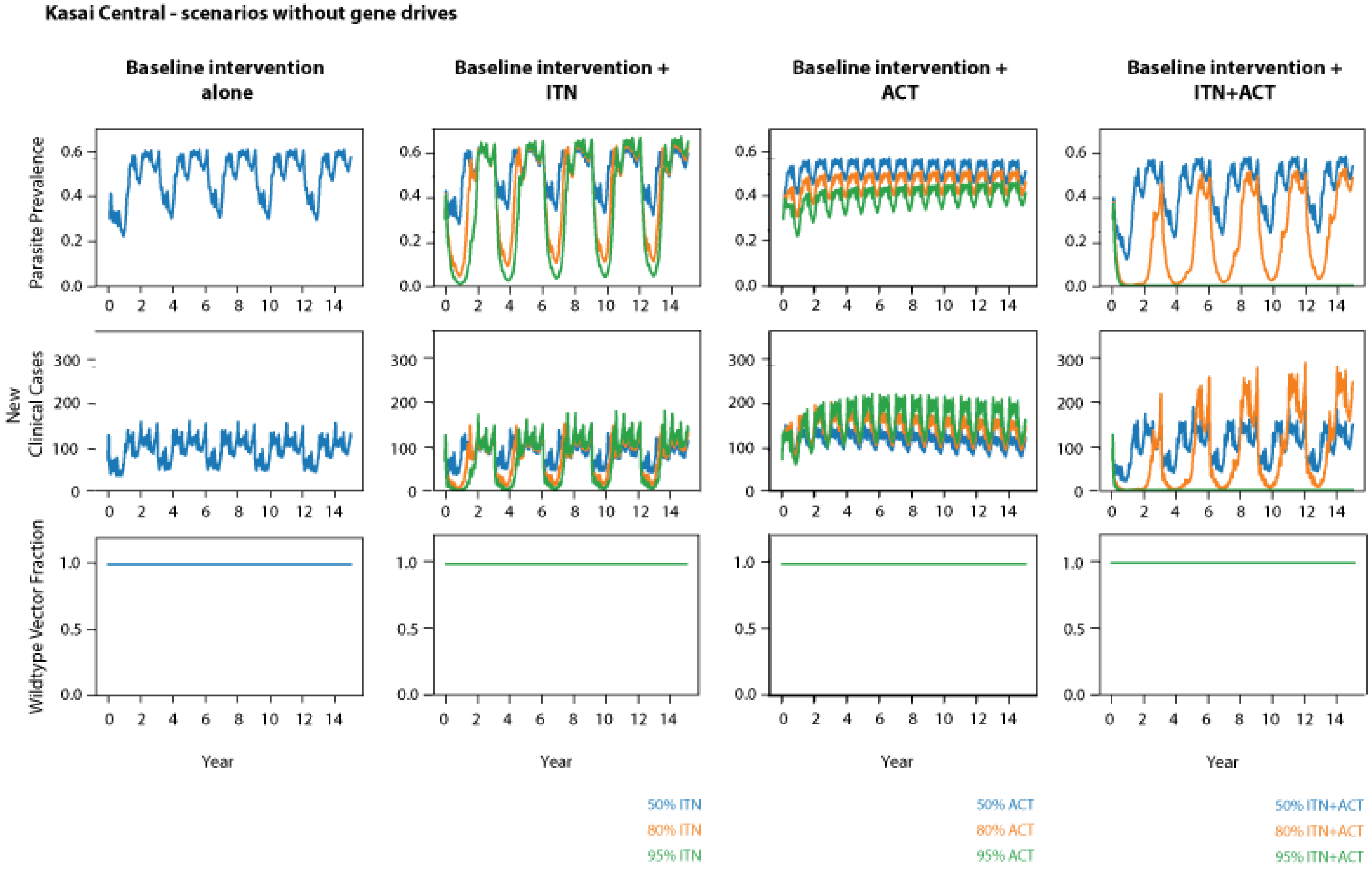
Kasai Central - scenarios without gene drives
Baseline intervention alone
Baseline intervention + ITN
Baseline intervention + ACT
Baseline intervention + ITN+ACT
Parasite Prevalence
New Clinical Cases
Wildtype Vector Fraction
Year
50% ITN
80% ITN
95% ITN
50% ACT
80% ACT
95% ACT
50% ITN+ACT
80% ITN+ACT
95% ITN+ACT

35

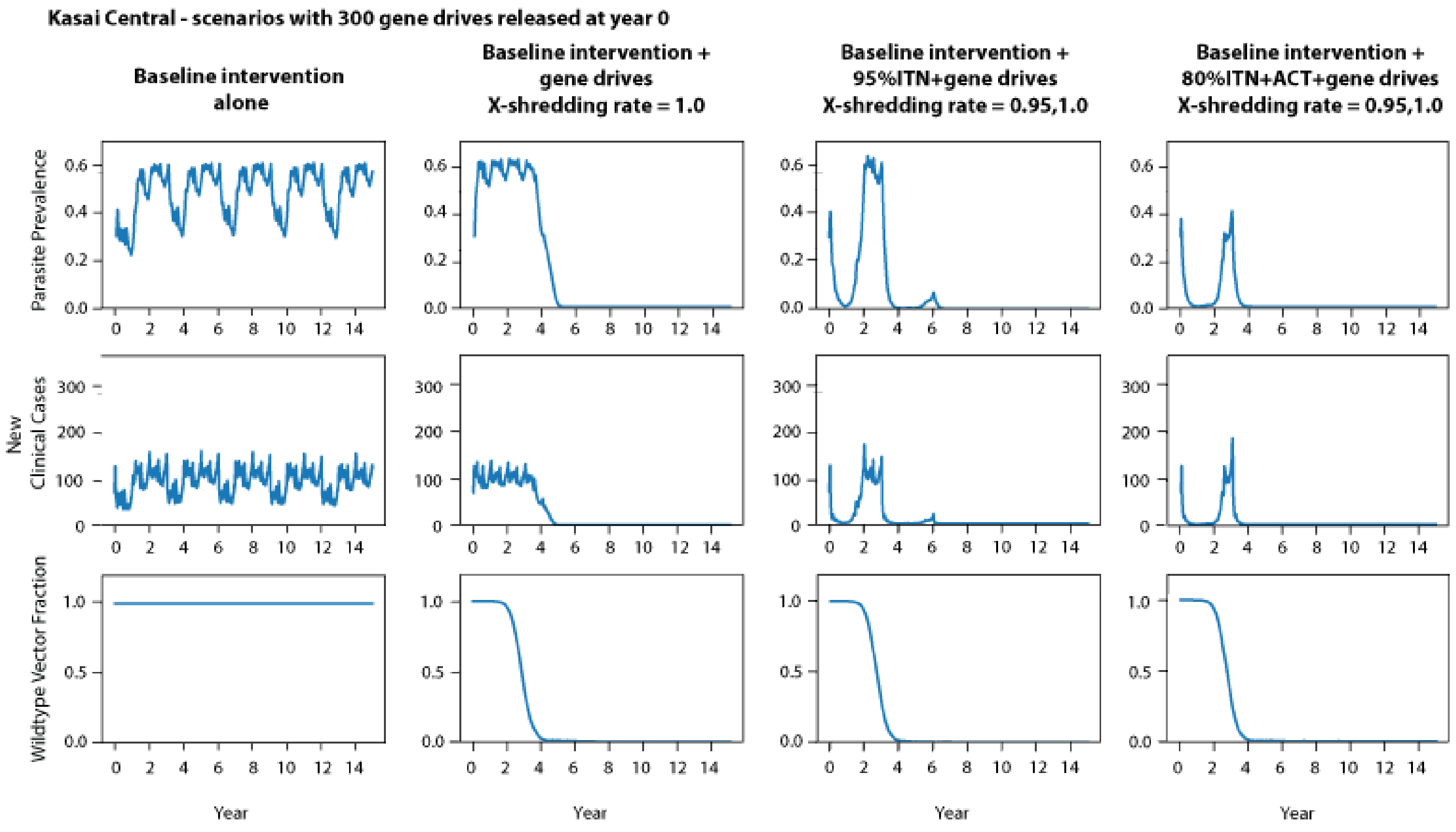
Kasai Central - scenarios with 300 gene drives released at year 0
Baseline intervention alone
Baseline intervention + gene drives X-shredding rate = 1.0
Baseline intervention + 95%ITN+gene drives X-shredding rate = 0.95,1.0
Baseline intervention + 80%ITN+ACT+gene drives X-shredding rate = 0.95,1.0
Parasite Prevalence
New Clinical Cases
Wildtype Vector Fraction
Year

36

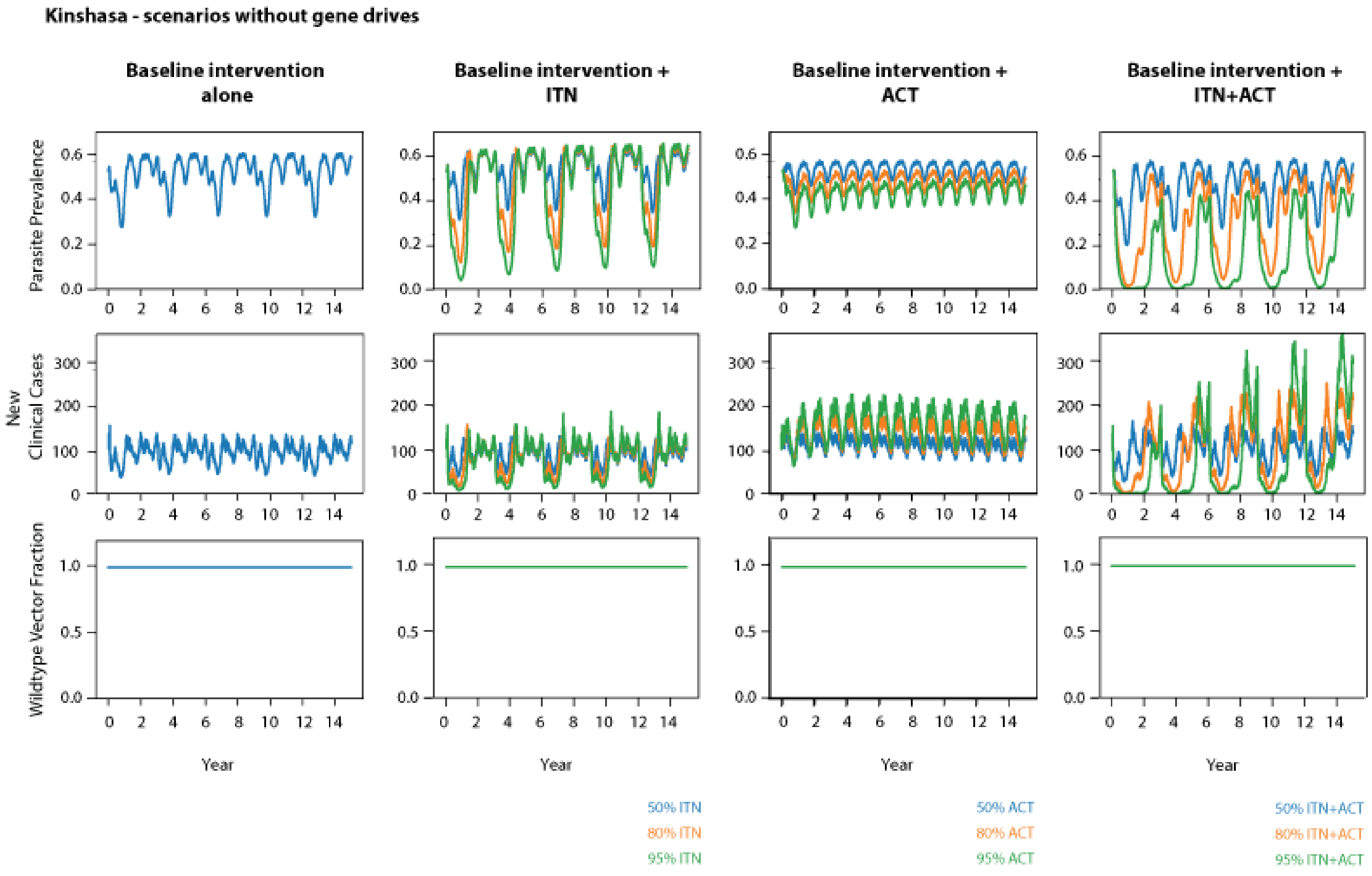
Kinshasa - scenarios without gene drives
Baseline intervention alone
Baseline intervention + ITN
Baseline intervention + ACT
Baseline intervention + ITN+ACT
Parasite Prevalence
New Clinical Cases
Wildtype Vector Fraction
Year
50% ITN
80% ITN
95% ITN
50% ACT
80% ACT
95% ACT
50% ITN+ACT
80% ITN+ACT
95% ITN+ACT

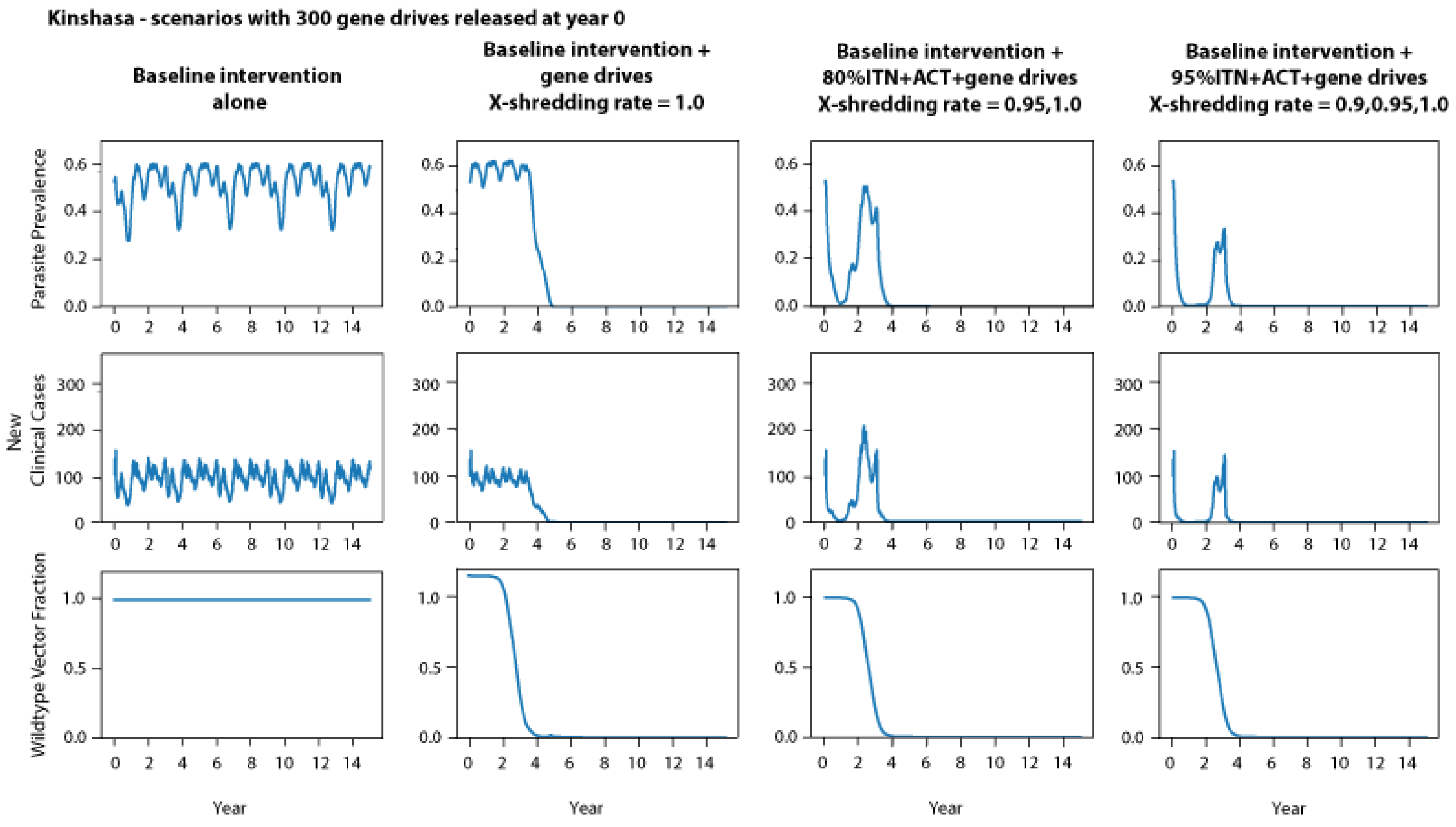
Kinshasa - scenarios with 300 gene drives released at year 0
Baseline intervention alone
Baseline intervention + gene drives X-shredding rate = 1.0
Baseline intervention + 80%ITN+ACT+gene drives X-shredding rate = 0.95,1.0
Baseline intervention + 95%ITN+ACT+gene drives X-shredding rate = 0.9,0.95,1.0
Parasite Prevalence
New Clinical Cases
Wildtype Vector Fraction
Year

38

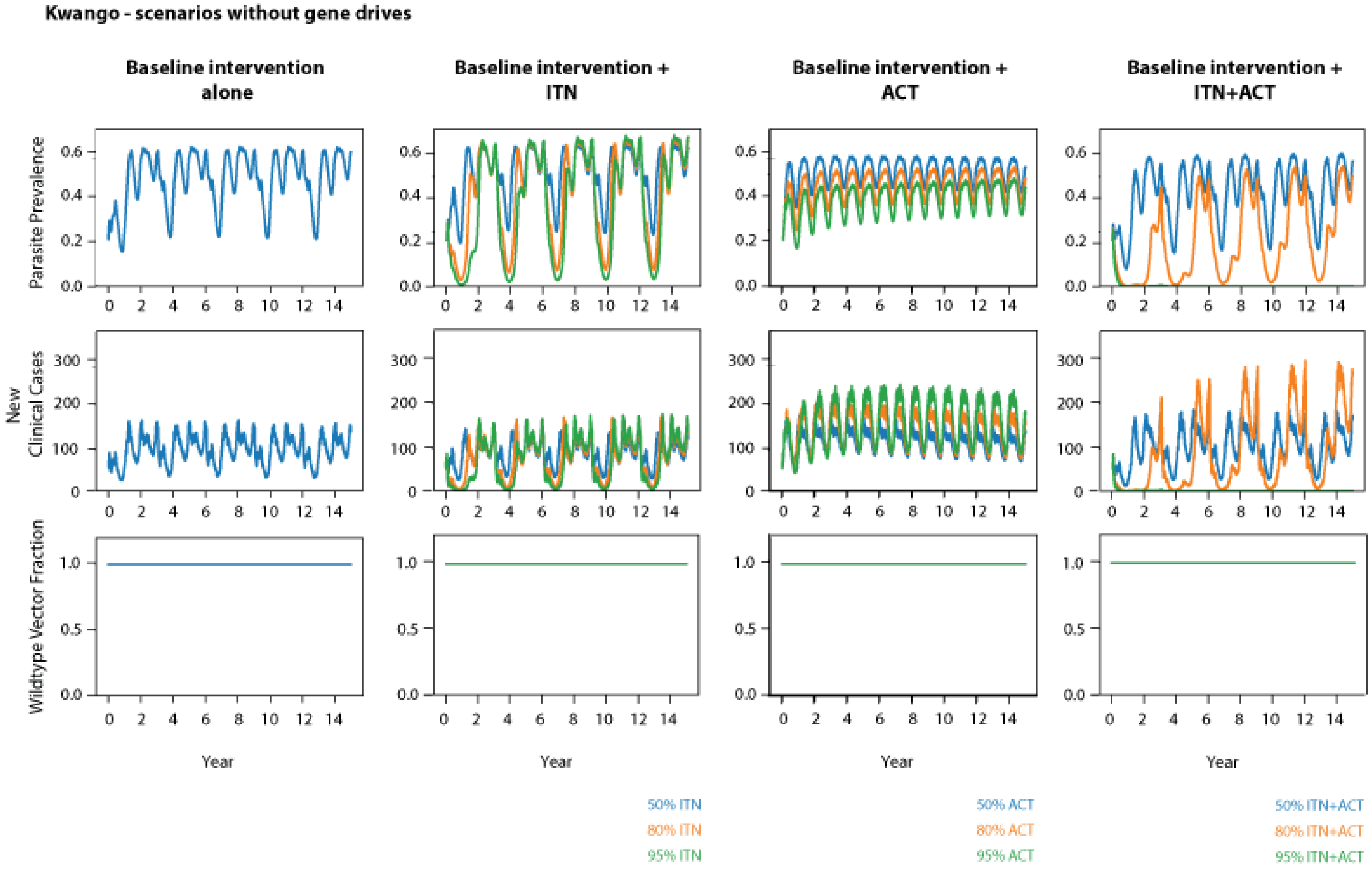
Kwango - scenarios without gene drives
Baseline intervention alone
Baseline intervention + ITN
Baseline intervention + ACT
Baseline intervention + ITN+ACT
Parasite Prevalence
New Clinical Cases
Wildtype Vector Fraction
Year
50% ITN
80% ITN
95% ITN
50% ACT
80% ACT
95% ACT
50% ITN+ACT
80% ITN+ACT
95% ITN+ACT

39

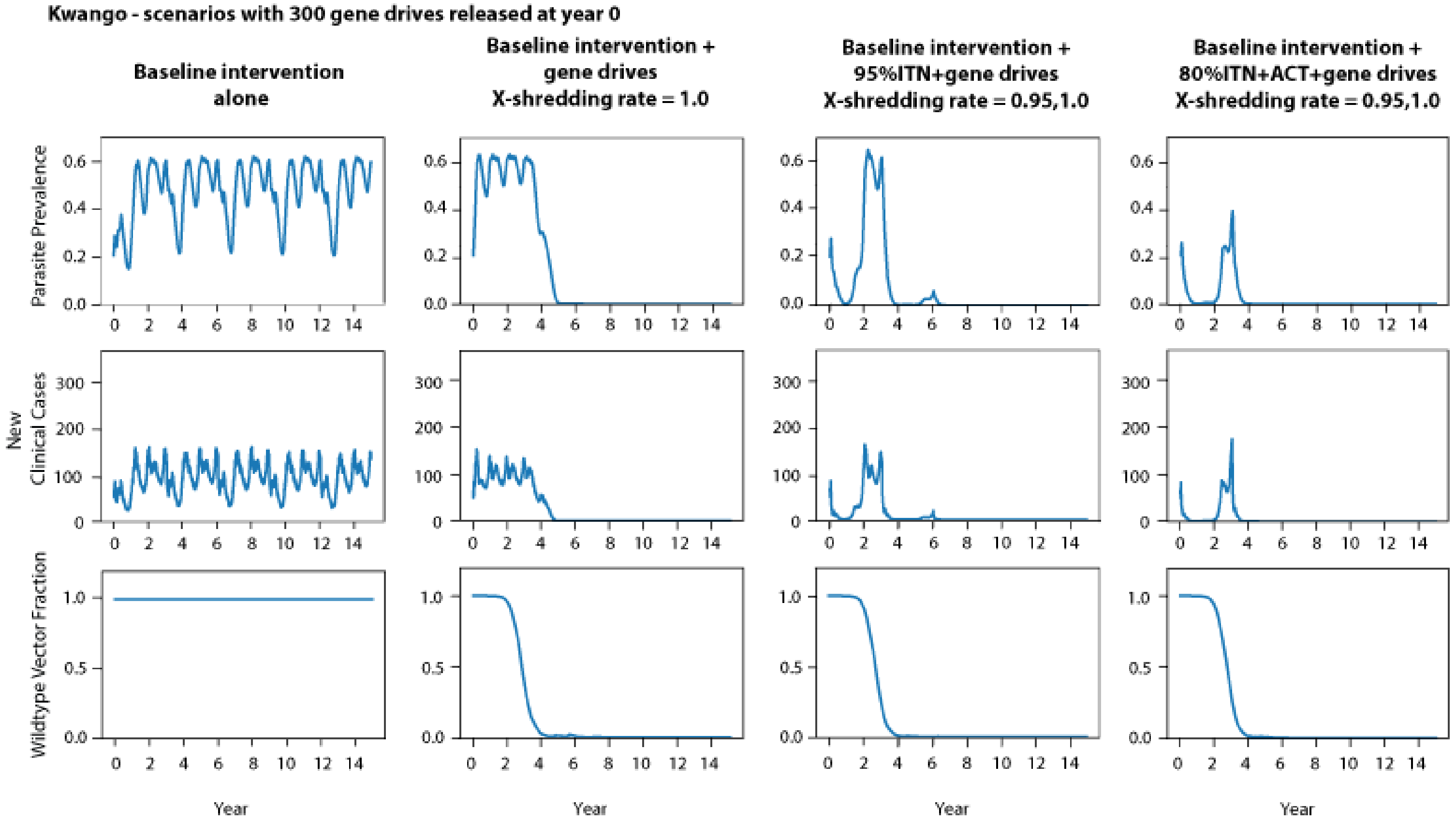

Kwango - scenarios with 300 gene drives released at year 0
Baseline intervention alone
Baseline intervention + gene drives X-shredding rate = 1.0
Baseline intervention + 95%ITN+gene drives X-shredding rate = 0.95,1.0
Baseline intervention + 80%ITN+ACT+gene drives X-shredding rate = 0.95,1.0
Parasite Prevalence
New Clinical Cases
Wildtype Vector Fraction
Year


40

Nord Ubangui - scenarios without gene drives

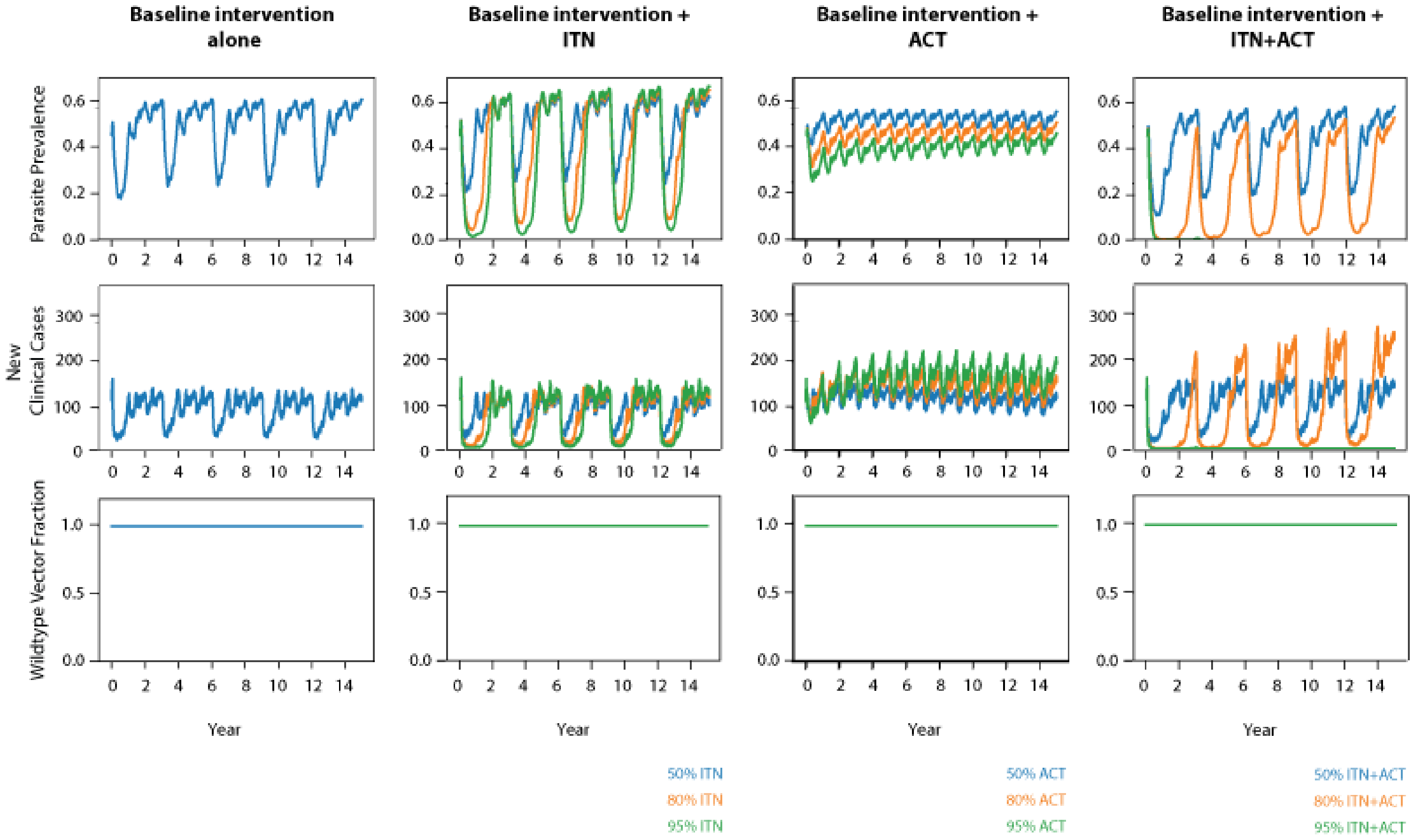

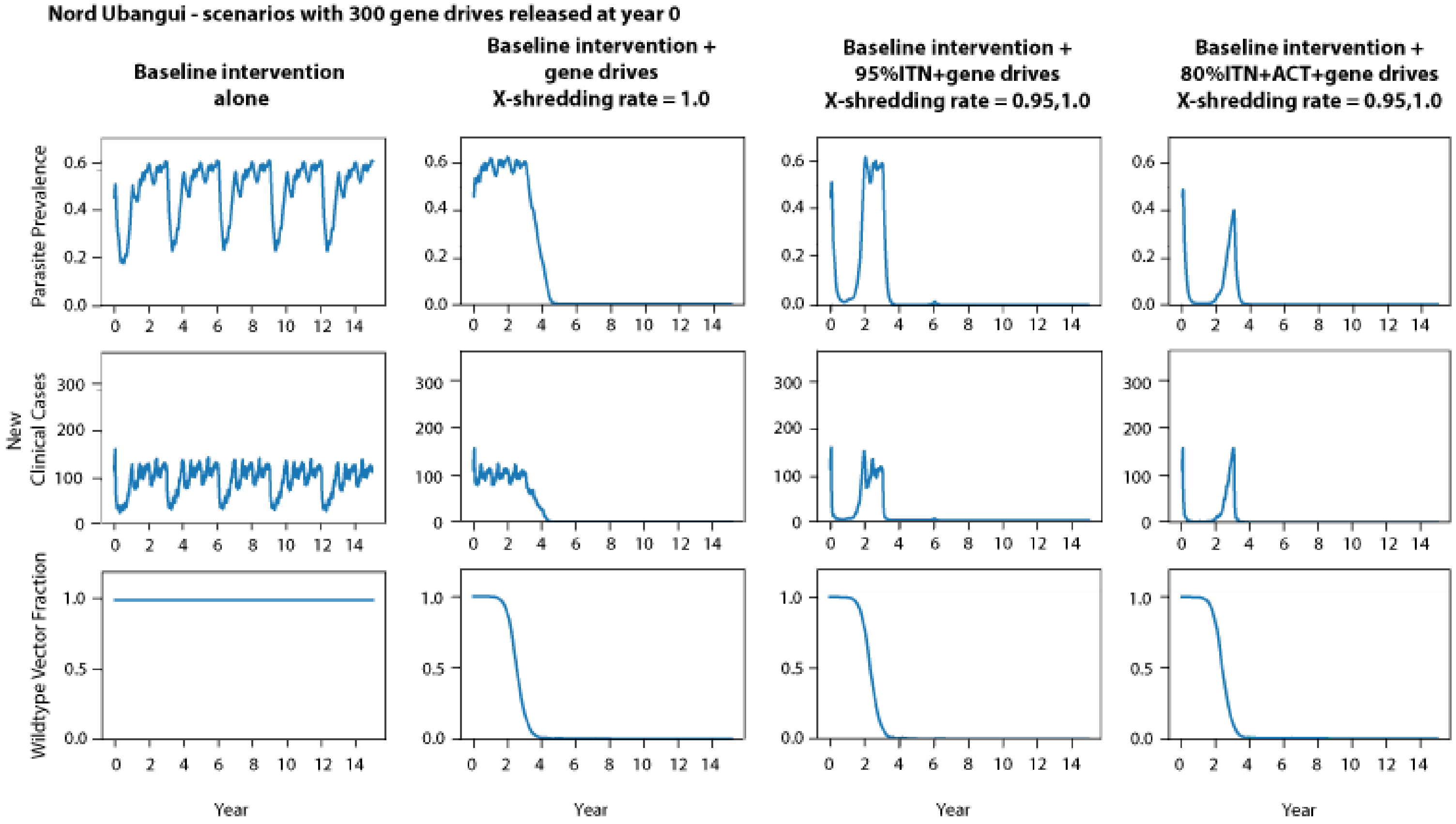
Nord Ubangui - scenarios with 300 gene drives released at year 0
Baseline intervention alone
Baseline intervention + gene drives X-shredding rate = 1.0
Baseline intervention + 95%ITN+gene drives X-shredding rate = 0.95,1.0
Baseline intervention + 80%ITN+ACT+gene drives X-shredding rate = 0.95,1.0
Parasite Prevalence
New Clinical Cases
Wildtype Vector Fraction
Year

42

### <u>Supplementary 5</u>: Reduced migration testing

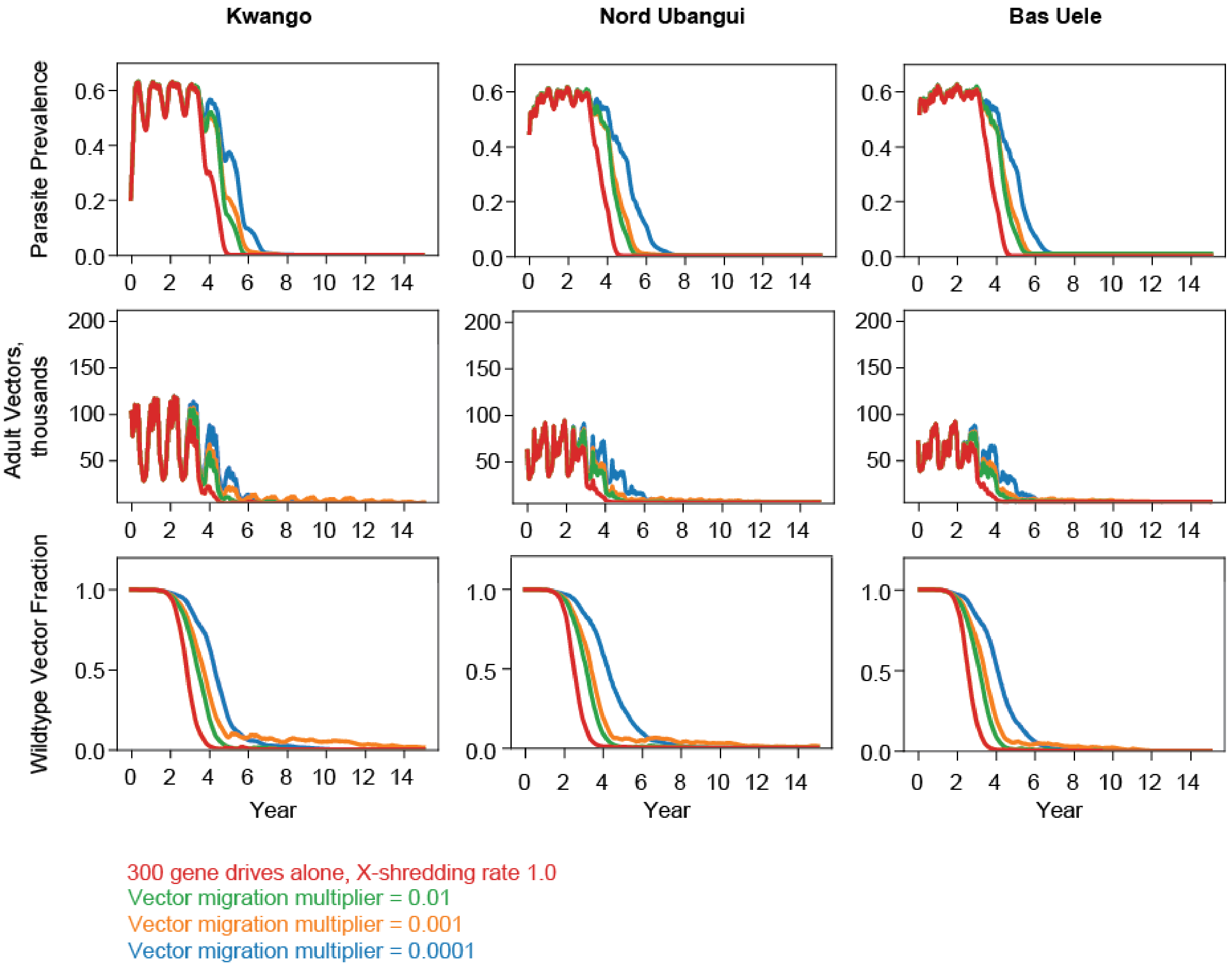

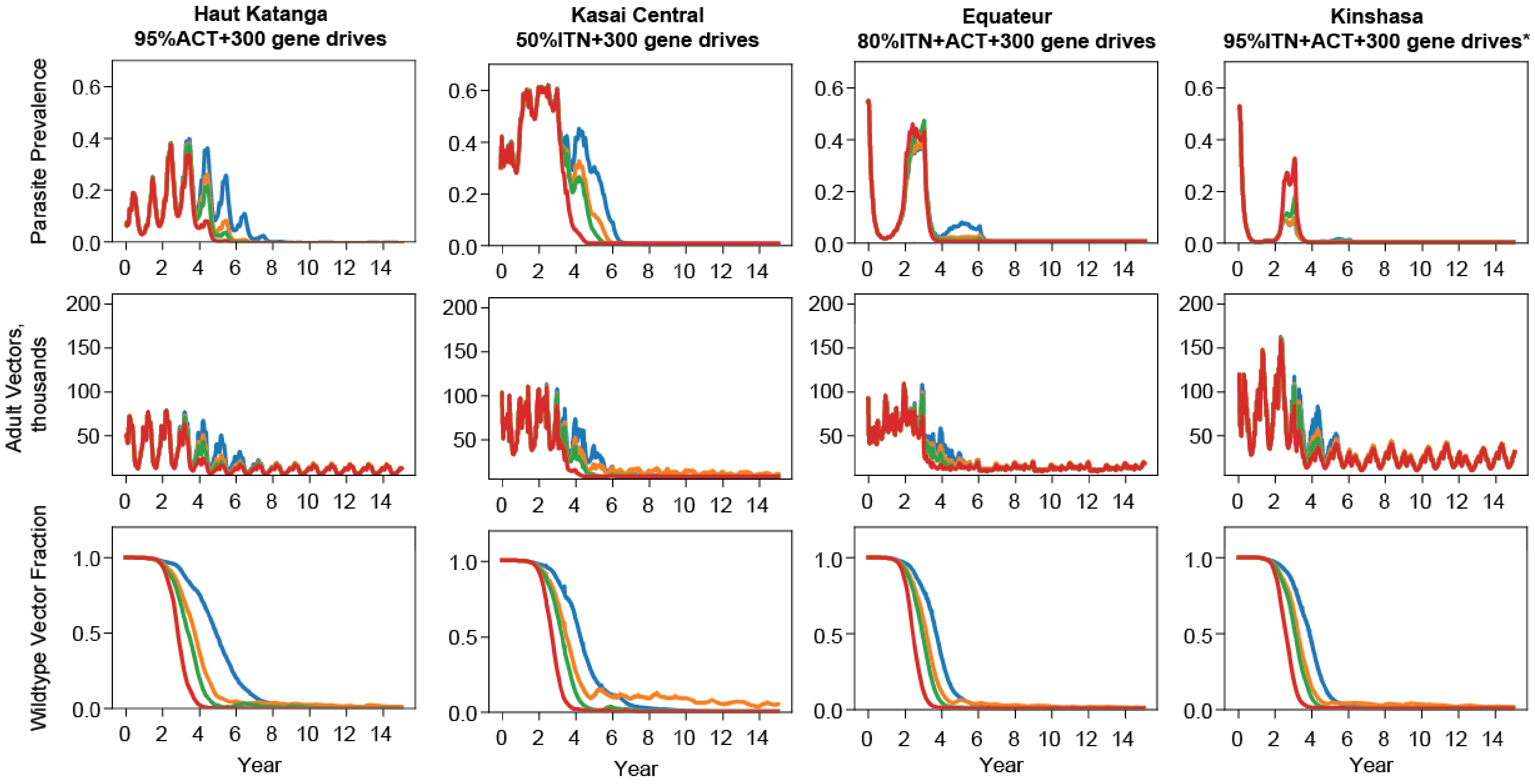


X-shredding rate 0.95 and 1.0, *X-shredding rate 0.9, 0.95, and 1.0
Vector migration multiplier = 0.01
Vector migration multiplier = 0.001
Vector migration multiplier = 0.0001

45 -

### **<u>Supplementary 6</u>: Cost-effectiveness analysis**

<u>Table S1</u> Average DALYs averted per year per one million population across all locations estimated from model's outputs in spatial simulation framework. Estimates of each scenario were compared with baseline scenario, which 50% ITNs and 19% ACT coverage were applied. For scenarios that included gene drives, only the estimates from scenarios that resulted in malaria elimination were included.

| | | | Average DALYs averted per year per one million population | | | |
|---|---|---|---|---|---|---|
| | | | Model's estimates | | | |
| | **Intervention** | **Coverage** | **Average over 15 years** | **The first interval: year 1-5** | **The second interval: year 6-10** | **The last interval: year 11-15** |
| Scenarios without gene drives | ITNs | 50% | -3,696 | -6,818 | -2,361 | -1,910 |
| | | 80% | 1,482 | 12,827 | -1,990 | -6,390 |
| | | 95% | 8,727 | 27,165 | 2,158 | -3,143 |
| | ACT | 50% | 1,680 | -7,733 | 2,506 | 10,266 |
| | | 80% | 12,962 | 10,390 | 10,212 | 18,283 |
| | | 95% | 21,437 | 25,261 | 15,883 | 23,169 |
| | ITNs & ACT | 50% | 8,004 | 14,973 | 4,432 | 4,606 |
| | | 80% | 33,477 | 60,693 | 21,288 | 18,451 |
| | | 95% | 72,706 | 81,875 | 69,014 | 67,230 |
| Scenarios with gene drives | 300 gene drive mosquitoes with X-shredding rates = 1.0 alone | NA | 57,298 | 2,888 | 82,542 | 86,464 |
| | ITNs plus gene drives with X-shredding rates = 0.95 and 1.0 | 80 | 57,561 | 12,201 | 81,580 | 78,904 |
| | ITNs plus gene drives with X-shredding rates = 0.95 and 1.0 | 95 | 68,222 | 43,505 | 75,420 | 85,741 |
| | ACT plus gene drives with X-shredding rates = 0.95 and 1.0 | 95 | 68,006 | 40,162 | 83,090 | 80,766 |
| | ITNs & ACT plus gene drives with X-shredding rates = 0.95 and 1.0 | 50 | 67,740 | 39,311 | 83,142 | 80,766 |
| | ITNs & ACT plus gene drives with X-shredding rates = 0.95 and 1.0 | 80 | 81,029 | 73,234 | 82,414 | 87,441 |
| | ITNs & ACT plus gene drives with X-shredding rates = 0.9, 0.95 and 1.0 | 95 | 85,307 | 86,609 | 81,819 | 87,492 |

<u>Notes for Table S1:</u>

1) NA: Not applicable
2) Green highlight: the scenario achieved malaria elimination
3) It is possible that the DALY averted results turned out to be negative figures in some scenarios since the combination of ITNs at 50% coverage and ACT at 19% coverage was applied in the baseline scenarios that were used as a comparator to reflect reality. For example, a 50%ITNs scenario means only ITNs at 50% coverage was applied as a single intervention in the scenario. Therefore, it is understandable that the lower efficacy of 50%ITNs alone could be observed once compared to the comparator in which the combination of 50%ITNs and 19%ACT was applied.
Negative DALYs averted are in red texts.
4) WHO estimated the DALYs averted using null (do nothing) scenario as a comparator.

Table S2 Average cost per DALY averted of interventions and combinations applied estimated from model's outputs.

| | | | | Scenarios without gene drives | | | | | | | | | | Scenarios with gene drives | | | | | | |
|---|---|---|---|---|---|---|---|---|---|---|---|---|---|---|---|---|---|---|---|---|
| | | | | Intervention(s) | | | | | | | | | | | | | | | | |
| | | | | ITNs | | | ACT | | | The combination of ITNs and ACT | | | | 300 gene-drive mosquitoes with X-shredding rates = 1.0 alone | ITNs plus gene drives with X-shredding rates = 0.95 and 1.0 | ITNs plus gene drives with X-shredding rates = 0.95 and 1.0 | ACT plus gene drives with X-shredding rates = 0.95 and 1.0 | ITNs & ACT plus gene drives with X-shredding rates = 0.95 and 1.0 | ITNs & ACT plus gene drives with X-shredding rates = 0.95 and 1.0 | ITNs & ACT plus gene drives with X-shredding rates = 0.9, 0.95 and 1.0 |
| | | | | Coverage (%) | | | | | | | | | | | | | | | | |
| | Parasite prevalence at the end of the 50-year run-in (%) | Coverage (%) | Interval | 50 | 80 | 95 | 50 | 80 | 95 | 50 | 80 | 95 | Bound | NA | 80 | 95 | 95 | 50 | 80 | 95 |
| | WHO's estimates for Afr E (4) | | | 49 | 42 | 41 | 21 | 14 | 12 | 43 | 35 | 28 | NA | NA | NA | NA | NA | NA | NA | NA |
| Estimates from model's outputs | 7.27 | Haut Katanga | Year 1-5 | -45 | 22 | 13 | -9 | 143 | 9 | 28 | 12 | 11 | Lower bound | -36 | 111 | 37 | 23 | 36 | NA | NA |
| | | | | | | | | | | | | | Upper bound | -355 | 641 | 205 | 184 | 200 | NA | NA |
| | | | Year 6-10 | -238 | -91 | 23 | 43 | 18 | 12 | 303 | 10 | 9 | Lower bound | 9 | 17 | 26 | 11 | 17 | NA | NA |
| | | | | | | | | | | | | | Upper bound | 96 | 142 | 89 | 95 | 96 | NA | NA |
| | | | Year 11-15 | -207 | -121 | 25 | 20 | 12 | 11 | 169 | 10 | 9 | Lower bound | 9 | 17 | 18 | 12 | 17 | NA | NA |
| | | | | | | | | | | | | | Upper bound | 99 | 98 | 91 | 97 | 99 | NA | NA |
| | 20.81 | Kwango | Year 1-5 | -60 | 51 | 26 | -24 | 23 | 9 | 47 | 11 | 8 | Lower bound | -340 | NA | 32 | NA | NA | 20 | NA |
| | | | | | | | | | | | | | Upper bound | -3,382 | NA | 175 | NA | NA | 101 | NA |

| | | | | | | | | | | | | | | | | | | | | |
|---|---|---|---|---|---|---|---|---|---|---|---|---|---|---|---|---|---|---|---|---|
| | | | Year 6-10 | -427 | -494 | -156 | 55 | 17 | 12 | 112 | 59 | 9 | Lower bound | 9 | NA | 17 | NA | NA | 18 | NA |
| | | | | | | | | | | | | | Upper bound | 85 | NA | 95 | NA | NA | 94 | NA |
| | | | Year 11-15 | -275 | -117 | -106 | 18 | 11 | 9 | 120 | 89 | 8 | Lower bound | 8 | NA | 16 | NA | NA | 18 | NA |
| | | | | | | | | | | | | | Upper bound | 82 | NA | 90 | NA | NA | 91 | NA |
| | 29.65 | Kasai Central | Year 1-5 | -69 | 51 | 26 | -29 | 11 | 8 | 46 | 11 | 8 | Lower bound | 11,661 | NA | 32 | NA | NA | 20 | NA |
| | | | | | | | | | | | | | Upper bound | 116,124 | NA | 176 | NA | NA | 103 | NA |
| | | | Year 6-10 | -175 | -245 | -161 | 64 | 20 | 14 | 157 | 68 | 9 | Lower bound | 9 | NA | 18 | NA | NA | 19 | NA |
| | | | | | | | | | | | | | Upper bound | 87 | NA | 97 | NA | NA | 97 | NA |
| | | | Year 11-15 | -220 | -102 | -81 | 20 | 11 | 9 | 142 | 121 | 9 | Lower bound | 8 | NA | 16 | NA | NA | 18 | NA |
| | | | | | | | | | | | | | Upper bound | 83 | NA | 91 | NA | NA | 92 | NA |
| | 45.03 | Nord Ubangui | Year 1-5 | -64 | 53 | 30 | -27 | 20 | 8 | 48 | 11 | 8 | Lower bound | 82 | NA | 31 | NA | NA | 20 | NA |
| | | | | | | | | | | | | | Upper bound | 816 | NA | 172 | NA | NA | 102 | NA |
| | | | Year 6-10 | -168 | -925 | -206 | 125 | 22 | 15 | 138 | 66 | 9 | Lower bound | 9 | NA | 17 | NA | NA | 19 | NA |
| | | | | | | | | | | | | | Upper bound | 88 | NA | 96 | NA | NA | 98 | NA |
| | | | Year 11-15 | -419 | -96 | -82 | 17 | 10 | 9 | 129 | 98 | 8 | Lower bound | 8 | NA | 16 | NA | NA | 17 | NA |
| | | | | | | | | | | | | | Upper bound | 82 | NA | 89 | NA | NA | 91 | NA |
| | 51.64 | Bas Uele | Year 0-5 | -81 | 61 | 29 | -36 | 19 | 8 | 48 | 12 | 10 | Lower bound | 65 | NA | NA | NA | NA | 20 | 16 |
| | | | | | | | | | | | | | Upper bound | 645 | NA | NA | NA | NA | 105 | 87 |
| | | | Year 6-10 | -181 | -606 | -322 | 117 | 22 | 15 | 140 | 170 | 9 | Lower bound | 9 | NA | NA | NA | NA | 19 | 18 |

| | | | | | | | | | | | | | | | | | | | | |
|---|---|---|---|---|---|---|---|---|---|---|---|---|---|---|---|---|---|---|---|---|
| | | | | | | | | | | | | | Upper bound | 87 | NA | NA | NA | NA | 97 | 96 |
| | | | Year 11-15 | -204 | -81 | -73 | 19 | 11 | 9 | 180 | 111 | 9 | Lower bound | 8 | NA | NA | NA | NA | 18 | 17 |
| | | | | | | | | | | | | | Upper bound | 82 | NA | NA | NA | NA | 91 | 90 |
| | **52.33** | **Kinshasa** | **Year 1-5** | -103 | 104 | 51 | -89 | 19 | 9 | 67 | 17 | 10 | Lower bound | 77 | NA | NA | NA | NA | 25 | 18 |
| | | | | | | | | | | | | | Upper bound | 765 | NA | NA | NA | NA | 129 | 97 |
| | | | **Year 6-10** | -170 | -366 | -1,602 | 110 | 20 | 13 | 181 | 69 | 23 | Lower bound | 9 | NA | NA | NA | NA | 19 | 18 |
| | | | | | | | | | | | | | Upper bound | 88 | NA | NA | NA | NA | 98 | 97 |
| | | | **Year 11-15** | -271 | -118 | -98 | 18 | 10 | 8 | 143 | 82 | 36 | Lower bound | 8 | NA | NA | NA | NA | 17 | 17 |
| | | | | | | | | | | | | | Upper bound | 81 | NA | NA | NA | NA | 91 | 90 |
| | **54.33** | **Equateur** | **Year 1-5** | -104 | 76 | 40 | -86 | 17 | 8 | 53 | 40 | 9 | Lower bound | 55 | NA | NA | NA | NA | 23 | 17 |
| | | | | | | | | | | | | | Upper bound | 547 | NA | NA | NA | NA | 120 | 91 |
| | | | **Year 6-10** | -180 | 5,457 | -706 | 115 | 21 | 13 | 143 | 75 | 19 | Lower bound | 9 | NA | NA | NA | NA | 19 | 18 |
| | | | | | | | | | | | | | Upper bound | 88 | NA | NA | NA | NA | 98 | 97 |
| | | | **Year 11-15** | -252 | -75 | -74 | 20 | 12 | 9 | 164 | 115 | 31 | Lower bound | 8 | NA | NA | NA | NA | 18 | 17 |
| | | | | | | | | | | | | | Upper bound | 83 | NA | NA | NA | NA | 92 | 91 |

Note for Table S2: 1) NA: Not applicable
2) The scenarios that could achieve malaria elimination when adding gene drives were highlighted in green.
3) $int: International Dollars
4) upper bound: upper bound price, lower bound: lower bound price

Table S3 Incremental cost-effectiveness ratio in year 1-5 after applying intervention(s)

| | | | | **ICER ($int per DALY averted)** | | | | | |
|---|---|---|---|---|---|---|---|---|---|
| | | | | **The first interval: year 1-5** | | **The second interval: year 6-10** | | **The last interval: year 11-15** | |
| | **Intervention** | **Coverage** | **Label** | **Lower bound** | **Upper bound** | **Lower bound** | **Upper bound** | **Lower bound** | **Upper bound** |
| Scenarios without gene drives | ITNs | 50% | A | dominated | dominated | negative | dominated | negative | dominated |
| | | 80% | B | dominated | dominated | negative | dominated | negative | dominated |
| | | 95% | C | 6.52 | 6.52 | negative | dominated | negative | dominated |
| | ACT | 50% | D | negative | negative | negative | negative | negative | negative |
| | | 80% | E | negative | negative | negative | negative | negative | negative |
| | | 95% | F | **First point** | **First point** | negative | **First point** | negative | **First point** |
| | ITNs & ACT | 50% | G | dominated | dominated | negative | dominated | negative | dominated |
| | | 80% | H | 0.43 | 0.43 | dominated | 2.82 | dominated | dominated |
| | | 95% | I | 0.23 | 0.23 | dominated | 0.25 | dominated | 0.30 |
| Scenarios with gene drives | 300 gene drive mosquitoes with X-shredding rates = 1.0 alone | NA | J | dominated | dominated | **First point** | 2.62 | **First point** | 2.76 |
| | ITNs plus gene drives with X-shredding rates = 0.95 and 1.0 | 80 | K | dominated | dominated | dominated | 2.90 | dominated | 3.42 |
| | ITNs plus gene drives with X-shredding rates = 0.95 and 1.0 | 95 | L | 1.67 | 10.56 | dominated | 3.23 | dominated | 3.07 |
| | ACT plus gene drives with X-shredding rates = 0.95 and 1.0 | 95 | M | 1.22 | 12.10 | 9.74 | 2.68 | dominated | 3.12 |
| | ITNs & ACT plus gene drives with X-shredding rates = 0.95 and 1.0 | 50 | N | 2.12 | 13.66 | 28.36 | 2.85 | dominated | 3.33 |
| | ITNs & ACT plus gene drives with X-shredding rates = 0.95 and 1.0 | 80 | O | 0.69 | 4.06 | dominated | 2.93 | 20.99 | 3.03 |
| | ITNs & ACT plus gene drives with X-shredding rates = 0.9, 0.95 and 1.0 | 95 | P | 0.51 | 3.14 | dominated | 2.92 | 17.93 | 2.99 |

**Keys for Table S3:**

- ICER: Incremental cost-effectiveness ratio
- Negative: the incremental cost and the incremental effect are negative
- Dominated: the incremental cost is positive, and the incremental effect is negative
- Vector control strategies that could reach malaria elimination were highlighted in green.
- **Incremental cost-effectiveness ratio (ICER)** is defined as the incremental change in cost, divided by the incremental change in its effectiveness. The first, second, and third points of each expansion path were highlighted in red, orange, and yellow accordingly.

### <u>Supplementary 7</u>: Costs of vector control approaches that involve the release of mosquitoes to modify vector population – A systematic scoping review.

<u>Background</u>

Vector control approaches that involve the release of mosquitoes to modify the vector population range from *Wolbachia* related technique to sterile insect production using radiation technique (1,2) . For *Wolbachia* related technique, the strategy proposed to infect mosquitoes with *Wolbachia* endosymbiotic that inhibits the viral replication and dissemination and eventually completely blocks vector-borne disease transmissions (3). The latter strategy, sterile insect technique (SIT) exposes male mosquitoes with low radiation in the laboratory production. This leads to sterilization of the male mosquitoes but maintain their copulation capacity (4). Once released to the environment, the sterile male mosquitoes mate wild female mosquitoes which would produce sterile eggs thereby eliminating the next generation progenies (5). Producing SIT insects at a large scale requires standardized mass-rearing procedures to produce good quality males that could compete with wild males to mate with females in the wild environment (6). Later development of these strategies includes the release of genetically engineered mosquitoes carrying dominant lethal allele that are capable of killing a subsequent generation (7). The produced OX513A mosquitoes are males that once mate with wild female mosquitoes will produce descendants that would not reach the adult stage due to lethal genes (8).

It is necessary for the decision to adapt these strategies specifically modifying mosquito vectors to be based on evidence of program effectiveness and cost-effectiveness of the interventions (9,10). However, evidence on costs and cost assessments of these strategies remain disparate. Data on costs could provide invaluable information for implementing vector control programs as malaria and other mosquito-borne infectious diseases continue to be a public health problem despite past and on-going control efforts (10). For sustenance of control efforts to achieve the disease elimination goal, it is important that the most cost-effective interventions are deployed (11). This supplementary is a systematic scoping review on costs and cost analysis of vector control approaches that involve the release of mosquitoes to modify the vector population focusing relevance on mosquito-borne diseases, especially malaria.

<u>Methods</u>

A systematic search of literatures in English language pertaining to costs of vector control methods to modify mosquito populations, published from 2010 to 2020, was performed (Diagram S1). Databases include National Center for Biotechnology Information (NCBI), Google Scholar, Crossref, and Scopus. Search terms include ‘Culicidae’ and ‘genetic engineering’ or ‘genetically modified’ and ‘costs’ or ‘cost analysis’ and ‘malaria’.

3,486 records of the initial search: 761 articles in PubMed Central, 13 articles in PubMed, 22 items in 13 books, 2480 articles in Google Scholar, 200 articles in Crossref, and 10 articles in Scopus, were screened for relevance, and those that included some forms of information on costs of new vector control intended to modify mosquito populations were assessed further in full texts for eligibility. 8 articles from reference lists thought to be relevant based on their titles alone were included in the full-text assessment. We excluded 3,428 articles that were duplicated, or full texts not provided. In full-text assessment, 66 articles were included based on selection criteria suggested costs or cost analysis of vector control methods intended to modify mosquito populations were described in monetary terms. 58 publications did not mention costs or mentioned but did not provide estimates in terms of monetary numbers were excluded. Details of literature selection procedures are in Diagram S1.

After eligible articles passed to the full-text assessment, we extracted details on publication year, country/region, target development, target disease, costs, the category of costs. Monetary cost data were first adjusted to US$ in the year of the initial study using historical exchange rates provided in the article if the adjustment was not supplied by the authors. If the currency year was not mentioned in the article, the publication date or date of article submission was used for the currency conversion. All monetary data were standardized to 2000 US$ using the US government consumer price index (CPI) data to adjust for inflation (12). All costs were adjusted to US$ value in year 2000 to allow comparison of the costs of interventions across data sources.

Diagram S1 Search strategy for costing studies

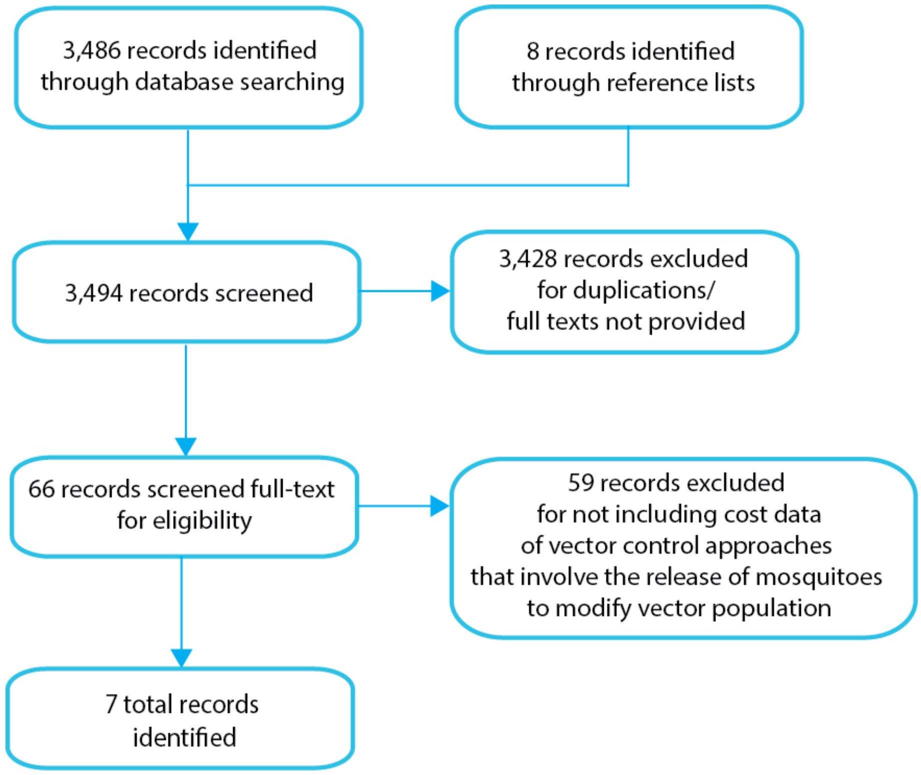


### Results

Four out of seven eligible records focused on or used data from research conducted in low- and middle-income countries and were published between 2011 and 2018. Most studies focused on Dengue using various techniques including *Wolbachia* infected mosquitoes, sterile insect technique (SIT), Release of Insects carrying Dominant Lethals (RIDL), and Genetically engineered (GE) mosquitoes i.e., Oxitec. Only one study specifically identified cost of in malaria control. One study (13) developed a cost model to calculate costs of genetic RIDL technology for dengue and was referenced by another study (14). In terms of costs, costs of the release of SIT insects range from 29.42 to 800 US$ (14) per million insects depending on cost categories. The costs of GM mosquitoes are varied and higher at the beginning of release with lower cost during later maintenance phase. *Wolbachia* infected mosquitoes gives low cost at 0.72 US$ per person. Table S4 summarized the study characteristics and details on costs described in each study.

Table S4 Summary of study characteristics and details on costs of vector control approaches that involve the release of mosquitoes to modify the vector population focusing relevance on mosquito-borne diseases.

| **References** | **Publication year** | **Country** | **Target development** | **Target disease** | **Target species** | **Cost originally indicated** | **Converted cost to year 2000** | **Category of costs** |
|---|---|---|---|---|---|---|---|---|
| Khamis D, El Mouden C, Kura K, et al. (14) | 2018 | Not specified | Sterile insect technique (SIT) mosquitoes | Malaria | Not specified | mean = 9.11 (min = 1.93, max = 25.36) US$ per 10,000 insect per day | 6.25 (1.32, 17.39) US$ per 10,000 insects per days | Operational costs |
| O'Neill SL, Ryan PA, Turley AP, Wilson G, Retzki K, Iturbe-Ormaetxe I et al. (15) | 2018 | Australia | *Wolbachia* infected mosquitoes | Dengue and other *Aedes* transmitted arboviruses | *Aedes aegypti* | *Wolbachia* infected mosquito: 1.07 US$ per person | 0.72 US$ per person* | Deployment cost |
| Meghani Z and BoeÈte C. (16) | 2018 | Brazil | Genetically engineered (GE) mosquitoes | Dengue | *Aedes aegypti* | 1.9 million US$ and 384,000 US$ for subsequent years per 50,000 urban population<br>Oxitec: 10 US$ per person (in year 2016) | 1.36 million US$ and 275,511.96 US$ for subsequent years per 50,000 urban population<br>Oxitec: 7.17 US$ per person** | Not specified but mentioned that the costs did not consider recurrent relicensing and subsidizing the cost |
| Alfaro-Murillo JA, Parpia AS, Fitzpatrick MC, Tamagnan JA, Medlock J, Ndeffo-Mbah ML, et al. (17) | 2016 | Middle-income countries (main country: Brazil) | GE mosquitoes (Oxitec) | Zika | *Aedes aegypti* (male) | 1.9 million US$ (in year 2015) in the first year and 384,000 US$ each year thereafter for an urban population of 50,000 | 1.38 million US$ (for 1st year) and 278,987.58 US$ per year afterward | Not stated |

| References | Publication year | Country | Target development | Target disease | Target species | Cost originally indicated | Converted cost to year 2000 | Category of costs |
|---|---|---|---|---|---|---|---|---|
| Undurraga EA, Halasa YA, Shepard DS. (18) | 2016 | Brazil, Mexico, Panama, Puerto Rico, and Thailand | Genetic modified (GM) mosquitoes | Dengue | *Aedes* | 25-75 US$ per person per year in suppression phase and 10-20 US$ per person per year in maintenance phase, based on preliminary estimates by Oxitec. | 17.94-53.81 US$ per person per year in suppression phase and 7.17**-14.35 US$ per person per year in maintenance phase | Overall cost estimate |
| Bellini R, Medici A, Puggioli A, Balestrino F, Carrieri AM. (19) | 2013 | India | SIT | Arboviral diseases in humans | *Aedes albopictus* (Skuse) | 40 US$ per million male mosquitoes per day | 29.42 US$ per million male mosquitoes per day | Production cost |
| Alphey N, Alphey L, and Bonsall MB. (13) | 2011 | Not specified | Genetic Release of Insects carrying Dominant Lethals (RIDL) technology (Release of Insects carrying a Dominant Lethal) | Dengue | *Aedes aegypti* | 1 US$ per 1000 insects (in year 2008) | 0.80 US$ per 1,000 insects | Construction and operational costs |
| | | | | | | 2.30 US$ per case averted (in year 2008) | 1.84 US$ per case averted | |
| | | | | | | Mean cost per person protected per year during the assessment period is 0.05–0.07 US$ or 0.52–0.68 US$ (release ratio 1 or 10, respectively in year 2008) | Mean cost per person protected per year during the assessment period is 0.04-0.07 US$ or 0.42-0.54 US$ (release ratio 1 or 10 respectively) | |

*lower bound cost applied in the main study.
**upper bound cost applied in the main study.

### Discussion

During the past 10 years, only handful costing evidence of vector control approaches that involve the release of mosquitoes to modify the vector population focusing relevance on mosquito-borne diseases, especially malaria, has been made available. When costs were mentioned, the studies were often not undertaken alongside an evaluation of the clinical and epidemiological effect of the methods of interest. This systematic scoping review is an early attempt to combine evidence on costs of the approaches.
